# Genomic and proteomic evidence for hormonal and metabolic foundations of polycystic ovary syndrome

**DOI:** 10.1101/2024.04.18.24306020

**Authors:** Loes M.E. Moolhuijsen, Jia Zhu, Benjamin H. Mullin, Natàlia Pujol-Gualdo, Ky’Era V. Actkins, Jasmine A. Mack, Hridya Rao, Bhavi Trivedi, Katherine A. Kentistou, Yajie Zhao, David Westergard, Jaakko S. Tyrmi, Gudmar Thorleifsson, Yanfei Zhang, Laura Wittemans, Amber DeVries, Kelly Brewer, Ryan Sisk, Rebecca Danning, Michael H. Preuss, Michelle R. Jones, Katherine S. Ruth, Marianne Andersen, Ricardo Azziz, Karina Banasik, Michael Boehnke, Linda Broer, Søren Brunak, Yee-Ming Chan, Daniel I. Chasman, Mark Daly, David A. Ehrmann, Bart C. Fauser, Lars G. Fritsche, M. Geoffrey Hayes, Chunyan He, Hongyan Huang, Irina Kowalska, Peter Kraft, Richard S. Legro, Nan Lin, Ruth J. Loos, Yvonne V. Louwers, Reedik Magi, Mark I. McCarthy, Laure Morin-Papunen, Jean V. Morrison, Cynthia Morton, Girish N. Nadkarni, Benjamin M. Neale, Henriette Svarre Nielsen, Mette Nyegaard, Sisse R. Ostrowski, Ole B.V. Pedersen, Erik Sørensen, Christina Mikkelsen, Christian Erikstrup, Kathrine A. Kaspersen, Mie T. Bruun, Bitten Aagaard, Henrik Ullum, Barbara Obermayer-Pietsch, Aarno Palotie, Mary P. Reeve, Andres Salumets, Richa Saxena, Timothy D. Spector, Bronwyn G. A. Stuckey, Unnur Thorsteinsdottir, André G. Uitterlinden, Margrit Urbanek, Sebastian Zollner, Genes and Health Research Team, DBDS Genomic Consortium, 23andMe Research Team, David A. Van Heel, Joel N. Hirschhorn, Kari Stefansson, John R.B. Perry, Unnur Styrkarsdottir, Scott G. Wilson, Terhi Piltonen, Triin Laisk, Marjo-Riitta Jarvelin, Kharis Burns, Anne E. Justice, Hannele Laivuori, Ken K. Ong, Mark O. Goodarzi, Lea K. Davis, Andrea Dunaif, Cecilia M. Lindgren, Joop S.E. Laven, Stephen Franks, Jenny A. Visser, Corrine K. Welt, Tugce Karaderi, Felix R. Day

## Abstract

Polycystic ovary syndrome (PCOS) and its underlying features remain poorly understood. In this genetic and proteomic study, we expand the number of genetic loci from 19 to 29, and identify 31 associated plasma proteins. Many risk-increasing loci were associated with later age at menopause, underscoring the reproductive longevity related to a larger functional ovarian reserve. Hormonal regulation in the aetiology of this condition, through metabolic and reproductive features, was emphasised. The proteomic analysis highlighted perturbations of metabolically-related biology that are typical in women with PCOS. A PCOS polygenic risk score was associated with adverse cardio-metabolic outcomes, with differing contributions of testosterone and BMI in women and men. Finally, while oligo- and anovulatory infertility are characteristic features of PCOS, we observed no impact of PCOS susceptibility on childlessness. We suggest that PCOS susceptibility confers balanced pleiotropic influences on fertility in women, and life-long adverse metabolic consequences in both sexes.

## Introduction

Polycystic ovary syndrome (PCOS) is the most common reproductive endocrinopathy in women^1^, with varied impacts across the lifespan. The diagnostic criteria include two of the following three features: hyperandrogenism, oligo-anovulation, and polycystic ovarian morphology (PCOM)^2^. PCOS is the most common cause of anovulatory infertility and is frequently associated with insulin resistance, which confers an increased risk of adverse metabolic outcomes such as type 2 diabetes (T2D)^3,4^. Previous large-scale genetic studies demonstrated that PCOS is a complex polygenic disorder that encompasses interactions between brain, metabolic and gonadal function^5^. High BMI and raised fasting insulin levels were identified as causal risk factors for PCOS, supporting the link to metabolic disease. Association at the *FSHB* genetic locus has highlighted the pituitary as a driver for PCOS^5,6^. Later age at menopause risk was also identified as causal, suggesting links to DNA damage and repair pathways in PCOS aetiology^7^. However, the small number of susceptibility loci identified thus far has limited exploration of the hereditary component^8^. There are no adequately powered prospective studies of women with PCOS beyond their reproductive years. Therefore, their long-term health outcomes remain unknown. In addition, our understanding of the genetic risk factors for PCOS on other health outcomes in women and men is not complete.

To address these limitations, we conducted a GWAS meta-analysis, which doubled the number of women with PCOS compared to previous GWAS^6^ (**Supplementary Table 1**), including data from 21,570 cases and 523,971 controls from 13 studies. We assessed the identified signals for association with a range of phenotypes encompassing three mechanisms thought to be related to PCOS: metabolic pathways, hormonal regulation including through the hypothalamic-pituitary-gonadal axis, and the oocyte/follicle complement. We also conducted a complementary proteomic-based analysis to further identify the biology related to this condition.

In addition, previous studies used genetic instruments to explore the causal links from a range of phenotypes to PCOS, but few have considered the downstream impacts of PCOS^9^. Many of these are likely to be a feature of “common soil” effects, a situation in which several conditions stem from the same source; in this case the adverse metabolic or hormonal background. However, there may be conditions in which PCOS has a specific, additional, adverse effect. In particular, while previous work has highlighted that variants associated with male-pattern balding increase the risk of PCOS^5^, whether PCOS risk variants impact disease risk in men has only been addressed to a limited extent^10^. Here the impact of PCOS on cardio-metabolic disease, disorders of reproductive organs (both cancer and non-cancer), and mental health are examined in both men and women.

## Results

### Genome-wide discovery for signals for PCOS

We identified 29 independent loci associated with PCOS (*P*<5×10^-8^) in the all-ancestries meta-analysis, of which 13 had not previously been reported^5,6,11–13^ (**Figure 1, Supplementary Table 2**). Novel GWAS signals for PCOS include a variant at *FTO* (rs8047587), confirming earlier findings from a candidate gene study^14^. The link to *FTO* strengthens the reported effect of increasing BMI on risk of PCOS^6,15^. Other novel PCOS signals have relevance to reproductive hormone pathways, such as *AMH* (rs732310), *INHBB* (rs6712151), and *SHBG* (rs1641518). Alongside the known European signal at *FSHB* (rs11031005) we report a signal at *FSHR* (rs13004711), replicating the association seen in Han Chinese for the first time in a majority European-ancestry GWAS^11^. Three of the 29 loci (*INHBB*, *NEIL2*, *DENND1A*) had evidence of secondary signals (within 500kb of the lead signal) (**Supplementary Table 3**).

**Figure 1.**
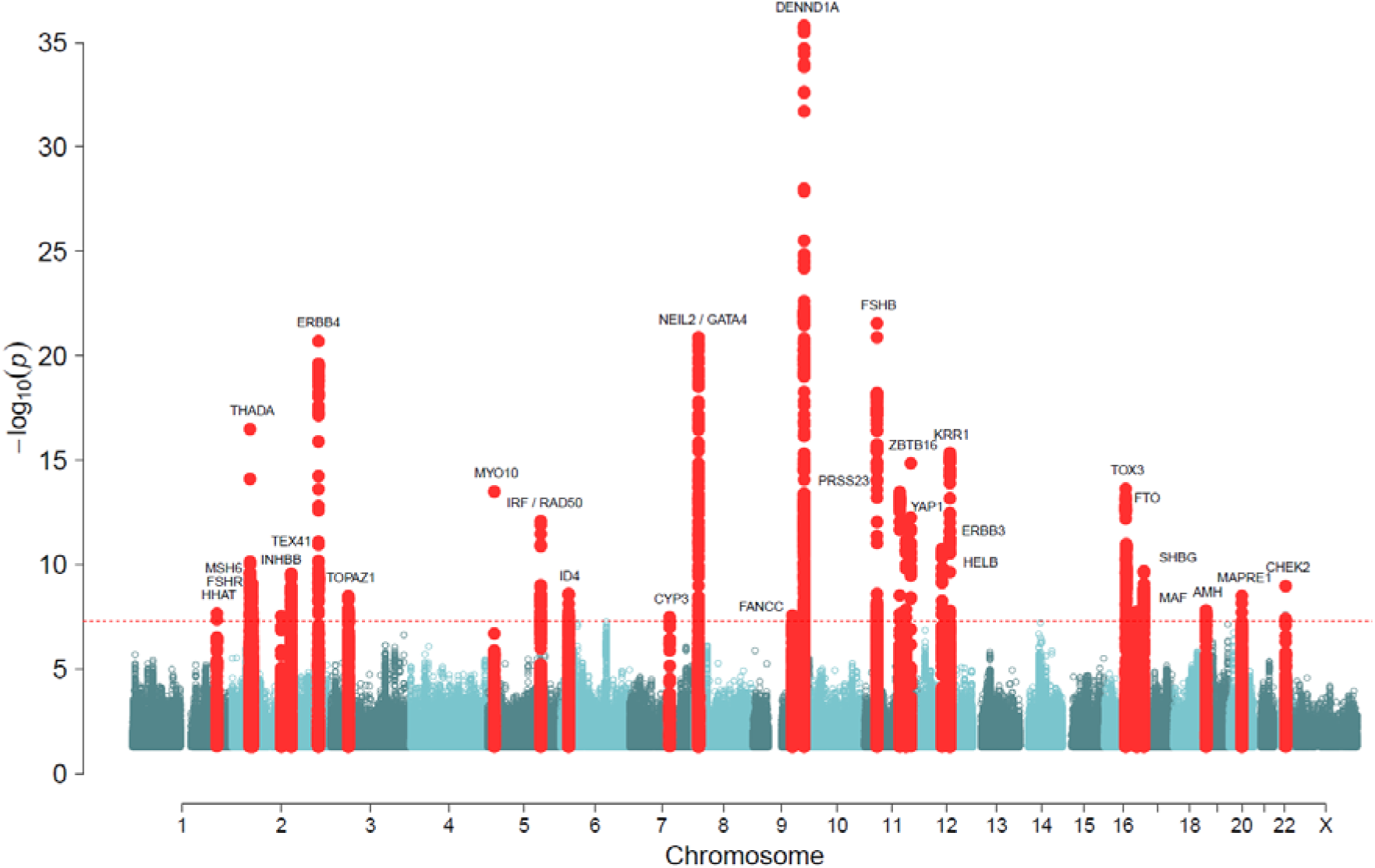
Manhattan plot showing the 29 genomic loci associated with PCOS. Variants within 300kb on either side of a genome-wide significant signal are highlighted in red. The dotted line indicates the genome-wide significance level. Gene names indicate the consensus PCOS gene at each locus.

We also performed a BMI-adjusted model in a subset of the cohorts (**Supplementary Table 4; Supplementary** Figure 1). In this adjusted analysis only the association at the *FTO* locus was substantially attenuated (*P*=0.019 after adjustment for BMI) (**Supplementary** Figure 2). To explore whether our GWAS findings were affected by differences in the diagnostic criteria used to assign a PCOS diagnosis, we stratified studies based on the method of case identification. There was no difference in the effects of the 29 PCOS signals by case definition (**Supplementary** Figure 3). Furthermore, effect sizes for each variant were comparable across individual studies (**Supplementary** Figure 4).

We assessed the transferability of 10 PCOS signals previously reported in East Asian women^11^ (**Supplementary Table 5**). Of these signals all but three (variants near *C9orf3*, *INSR* and *SUMO1P1*) had statistically significant associations in our cohort (*P*<0.005). The *SUMO1P1* minor allele was more common and had a relatively smaller effect size (OR=1.03) with borderline evidence for association (*P*=0.0058) in our data. The variant at *C9orf3* had a frequency in our data of approximately half that in the Chinese data, whereas the variant in *INSR,* coding for the insulin receptor, was more common in our data. Although rare deleterious variants in *INSR* cause a severe PCOS phenocopy^16^, previous studies have not demonstrated a genome-wide PCOS association at the *INSR* locus in women of European ancestry^17^. The relative frequency of the *INSR* variant suggests that there might be a gene-environment interaction that explains the different findings between these two studies or that the Han Chinese variant tags a haplotype that is not present in Europeans.

### Fine-mapping and genes of interest

Fine-mapping of the identified PCOS loci was performed using a LD reference based on European ancestry women in the MyCode/DiscovEHR and, separately, using the BioVU study (**Supplementary Tables 6 and 7**). At seven of the loci, both analyses resolved to the same sentinel variant. At *LLPH*, rs117568227 was the only variant in the 95% credible set (PP>0.99) in both studies, and at *YAP1,* rs7925543 was the only variant (PP=0.64 in MyCode/DiscovEHR and 0.79 in BioVU). Another 10 PCOS loci resolved to 20 or fewer variants across the two analyses.

We used two complementary approaches to link potential PCOS risk genes, which we call “consensus genes”. First, we performed a literature review of all genes within 500kb of the 29 signals to identify genes with a reported link to one of four processes - i) reproductive function, ii) steroid metabolism and sex-hormone levels, iii) pathways related to metabolic syndrome, and iv) DNA damage repair, selected due to reported links between PCOS and age at menopause for which DNA damage repair is the dominant process^7^ (**Figure 2**). Second, we used the GWAS-to-Gene bioinformatic approach that leverages data on eQTLs, pQTLs, predicted deleterious variants, and variant based scoring methods to rank genes based on their causal likelihood (**Supplementary Table 8**)^18^. Findings from these two approaches were then harmonised for each PCOS signal (**Supplementary Table 9**).

**Figure 2.**
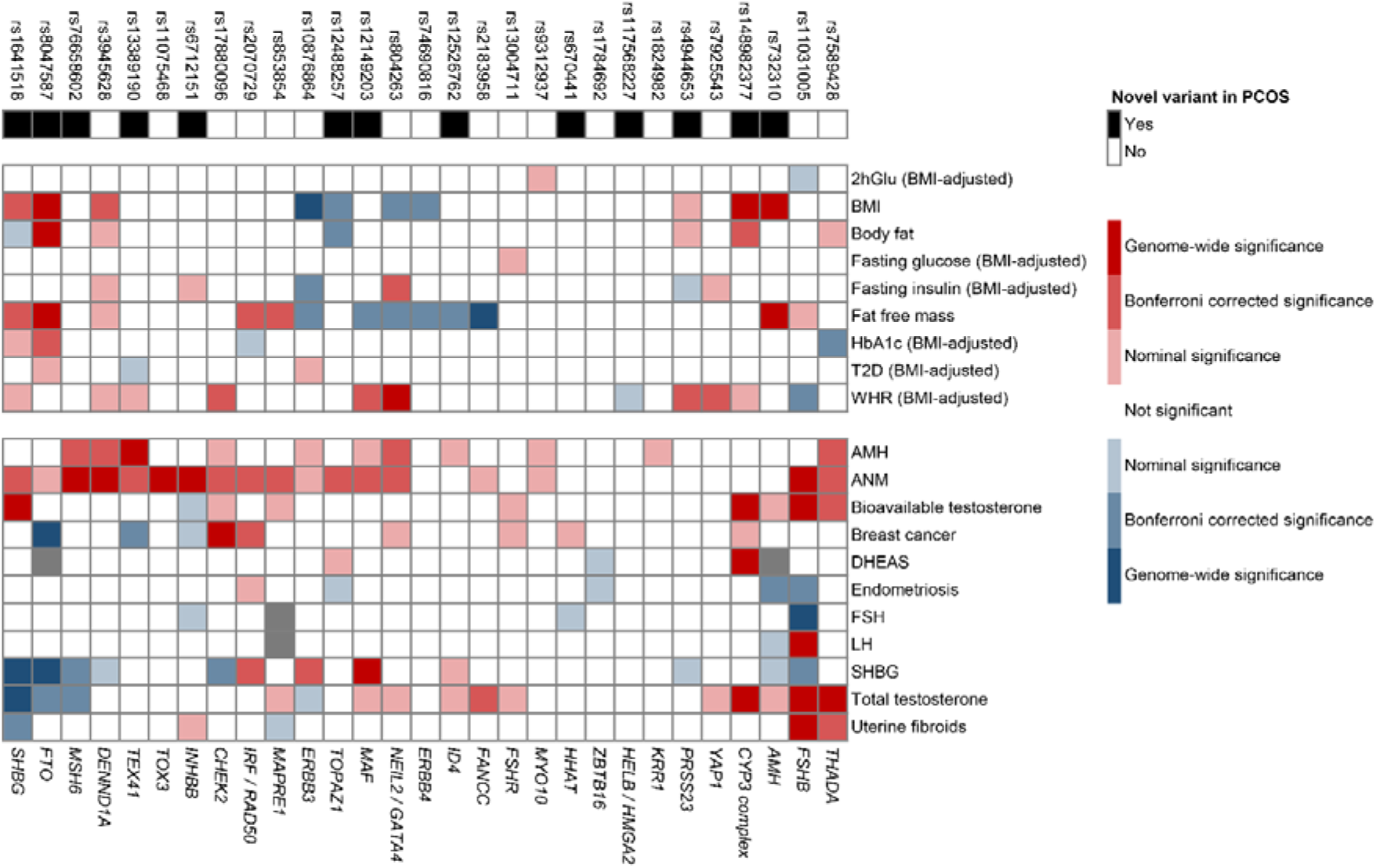
Heatmap of GWAS associations for the 29 PCOS loci with other relevant traits. Direction and the strength of association between the 29 PCOS risk-increasing alleles with 20 other relevant traits and diseases with available GWAS summary statistics. Colour coding indicates strength and direction (z-scores) of associations: positive (red) or negative (blue). In the upper row of the heatmap, novel PCOS loci identified in this study (black boxes) and previously reported loci (white boxes) are shown with corresponding lead variants. Grey boxes indicate missing variant-trait association data. Genes are presented in the lower x-axis as ‘consensus gene’. BMI=body mass index, T2D=type 2 diabetes mellitus, WHR=waist:hip ratio, AMH=anti-Müllerian hormone, ANM=age at natural menopause, FSH=follicle stimulating hormone, LH=luteinizing hormone, SHBG=sex hormone binding globulin.

In sixteen cases both approaches prioritised the same gene. In others, there was a strong rationale for prioritizing the literature-based gene instead of the bioinformatics-identified gene, such as *SHBG* over *ATP1B2* at rs1641518, particularly because the variant was also genome-wide significantly associated with circulating SHBG levels (**Figure 2**). The rs732310 variant was assigned to the *AMH* gene, based on the known functions of AMH in inhibiting recruitment of ovarian follicles from the primordial follicle pool, inhibiting FSH-sensitivity of growing follicles and regulating GnRH-dependent LH pulsatility^19,20^. Although rs732310 shows no association with AMH levels (**Figure 2**), the GWAS data used for the AMH analysis was derived from normo-ovulatory women^21^, and may not reflect variations in AMH levels in women with PCOS, in whom the expression pattern of AMH differs from normo-ovulatory women^22^.

### Relationship of the identified loci with other phenotypes

Several of the 29 PCOS variants had previously been associated in GWASs for age at menopause (14 variants), age at menarche (six), female testosterone levels (seven), BMI (eight), and male-pattern baldness (two) (see **Supplementary Table 10** for a complete list). We annotated our signals for the 29 PCOS loci with publicly available GWAS results, focusing in particular on the relationships between these variants and a range of metabolic, reproductive and hormonal phenotypes, given the suspected relationships to PCOS. Notably, all 29 PCOS signals showed at least nominal significance with one or more metabolic, reproductive and/or hormonal trait(s), providing evidence to support the multifactorial aetiology and comorbidities (**Figure 2, Supplementary Tables 11and 12**).

In validation of the substantial overlap between signals for PCOS and age at menopause (ANM), eight signals showed evidence of colocalization (PP≥0.75). PCOS signals at *FSHB*, *DENND1A*, *TOX3*, *RAD50*, and *MAF/MAFTRR* were associated with ANM (**Supplementary Table 13**); conversely the reported ANM signals at *FSHB, DENND1A, CASC22*, *BMP4*, *PPARG*, and *MAF/MAFTRR* were associated with PCOS (**Supplementary Table 14**). At all eight colocalised signals, the PCOS risk-increasing allele conferred later ANM. Of these loci, the *FSHB* signal has been well described to impact other reproductive phenotypes including age at menarche^23^, and dizygous twinning^24^. Other shared loci are also related to breast cancer pathways - *TOX3*, *CASC22 and RAD50*. Interestingly, the shared *PPARG* locus suggests an effect of metabolic pathways independent of those related to BMI.

A number of the PCOS-associated loci also showed strong effects on hormone levels, in particular on SHBG levels (including rs1641518 near *SHBG*). Approximately 30% (8/29) of the PCOS risk increasing alleles had nominal association with lower SHBG levels. There were 2 loci in which PCOS risk alleles were associated with lower SHBG and higher total testosterone, suggesting a relationship driven by higher androgens, whereas 3 other loci, including SHBG and FTO had lower SHBG and lower total testosterone, suggesting an SHBG mediated effect (**Figure 2**). Three of the signals had evidence of colocalization with SHBG levels including the *CYP3* complex, *FTO* and *FSHB* (**Supplementary Table 15**); notably this did not include the signal at *SHBG*^25^. The signal at *FTO* is likely due to the established links between an increase in BMI and decrease in SHBG levels^26^. The *CYP3* complex metabolizes estradiol and testosterone^27^, with mouse knockouts showing substantially increased free testosterone levels^28^ and thus the *CYP3* risk allele may result in decreased SHBG. The *FSHB* locus is associated with an increased LH, which stimulates androstenedione, and therefore testosterone^29^, which would lower SHBG.

There was additional evidence for the role of PCOS-associated loci impacting regulation of gonadotropins and the functional ovarian reserve. The PCOS risk-increasing allele at the *FSHB* locus was associated with lower levels of FSH and *INHBB* was nominally associated with lower FSH. Although the signal at *FSHR* was not associated with FSH levels in 2,913 Europeans, a variant found in Han Chinese women, which shows some evidence of LD (R^2^=0.3 in Europeans) with the variant reported here, was associated with lower FSH levels ^30,31^ (**Figure 2, Supplementary Table 11**)*. FSHR* has also been recently linked to rates of twinning and FSH levels using gene-based approaches^24^. At twelve loci, PCOS risk alleles were also nominally associated with higher AMH levels (*P*<0.05), with 6 additional loci nominally significant. Nine of these twelve loci overlap with the loci related to age at natural menopause and are all associated with higher AMH levels and later age at menopause. AMH is measured clinically to indicate functional ovarian reserve, and its concentrations are strongly related to the age at which menopause occurs. Loci that are associated with both elevated AMH levels and later age at menopause imply their involvement in the establishment and preservation of the functional ovarian reserve as a fundamental element of PCOS.

### Protein-based analysis

A total of 31 plasma proteins levels were phenotypically associated with ‘ovarian dysfunction’ in women defined as International Classification of Disease (ICD) 10 category E28 which includes PCOS (all P<3.4×10^-5^, **Figure 3, Supplementary Table 16**). These included recognized metabolic disease-associated proteins such as PCSK9, LDLR, FURIN, FABP1 and FABP4. Other associated proteins metabolise hydroxysteroids, retinol and lipids, including ALDH1A1 and ADH4, which may regulate the metabolic response to a high fat diet^32,33^. Other proteins are potential contributors to diabetes and metabolic disease, GGT1^34^, or their complications, SORD^35^. Finally, there were enzymes important for biosynthesis of progesterone or testosterone, GSTA1 and GSTA3^36^, and fertilisation and implantation, PAEP^37^. We used STRING to perform a combined pathway-based analysis using proteins drawn from either the protein-based approach or the GWAS associated loci (**Supplementary** Figure 5**, Supplementary Tables 17 and 18**)^38^. In general, the two discovery methods highlighted different biology, with only one pathway, *benzaldehyde dehydrogenase activity*, driven by both proteomic and GWAS findings (which included *FANCC,* a known DNA damage repair gene). The results did include evidence of enrichment or pathways representing androgen binding, and ovarian follicle development neuregulin receptor activity, the PCSK9-LDLR complex consistent with the lipid abnormalities of PCOS^39,40^.

**Figure 3.**
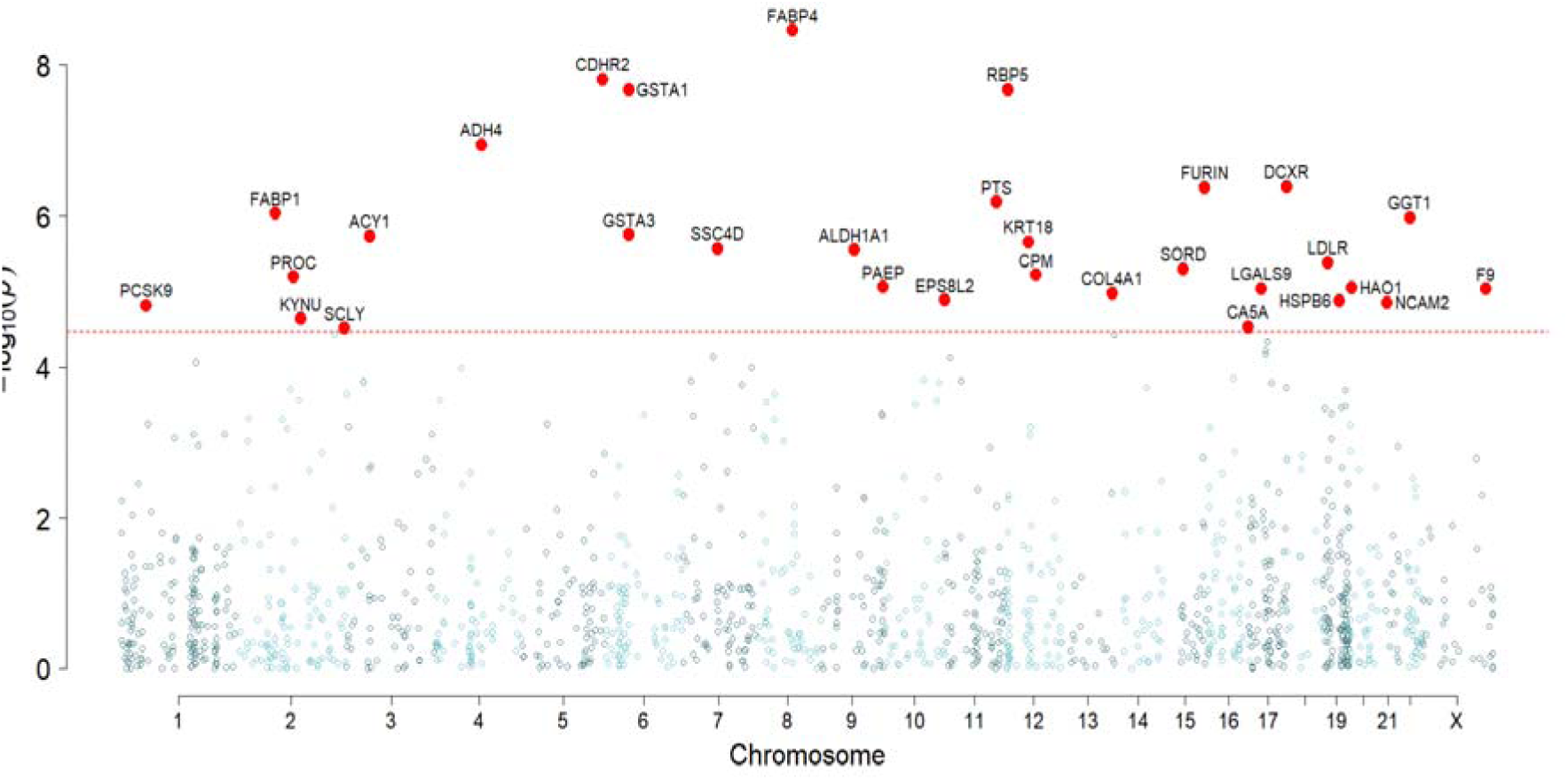
Manhattan plot showing the 31 plasma proteins associated with ovarian dysfunction. Ovarian dysfunction was defined by ICD10 code E28, the supra-category that includes PCOS.

In order to build causal pathways, we identified plasma proteins whose levels were associated with each of our PCOS GWAS signal variants, resulting in a total of 299 proteins (*P*<3.4×10^-5^, **Supplementary Table 19**). Of note, the PCOS signals at *ERBB3, ERBB4* and *ZBTB16* were associated with plasma levels of their encoded proteins, providing further support for these consensus genes. The PCOS signal at *SHBG* was associated with levels of TNSF12 and TNSF13, encoded by genes in the same region, and related to apoptosis and regulation of steroidogenesis ^41^. One PCOS signal at *RAD50/IRF1* accounted for 199 protein associations. This signal lies in a gene rich region on chromosome 2 that contains a large number of immune-related genes - *IRF1*, *IL4*, *IL5* and *IL13* and likely has a widespread impact on the plasma proteome. Another PCOS signal, overlapping the established obesity signal at *NEIL2*/*GATA4*, was associated with 29 plasma proteins. Other associated proteins included leptin, which is higher in obesity; PPY, a regulator of food intake; and several fatty acid binding proteins, which regulate fatty acid uptake in adipose cells.

Having identified these 299 proteins, we then assessed if any showed evidence of association with a diagnosis of ICD E28 (ovarian dysfunction), based on a threshold of *P*<1.7×10^-4^ (0.05/299). This two-stage approach resulted in nine variant-protein pairs. Three proteins were associated with the PCOS signal at *MAF* (CDHR2, IGFBP2, CPM), one with *ERBB3* (NCAM2) and five with *FTO* (ADM, FABP1, FABP4, LEP and SSC4D) (**Supplementary Table 20**). The *FTO*-locus related proteins are likely to be wholly explained the adiposity effect at this locus; this either as a result of the casual BMI association, or via a common soil effect. This is further highlighted by the fact that, of these proteins, FABP4 and LEP have recently been shown to be linked to aging specifically in adipose tissue^42^, which would suggest that changes in the levels are due to a primarily BMI effect. To understand the direction between individual plasma proteins and PCOS, we compared the variance in each trait explained by its respective PCOS signal (variance in BMI was also compared for those proteins associated with *FTO*) (**Figure 4**). The approach is based on the assumption that if a variant was associated with greater explained variance in protein levels compared to PCOS, it was unlikely that protein was affected as a result of PCOS. All five *FTO* associated proteins, NCAM2 associated with *ERBB3*, and IGFBP2 associated with *MAF*, appeared to be upstream (i.e. more likely to be determinants, or common soil) of PCOS. The two other proteins associated with *MAF,* CDHR2 and CPM, may have their levels altered as a consequence of PCOS.

**Figure 4.**
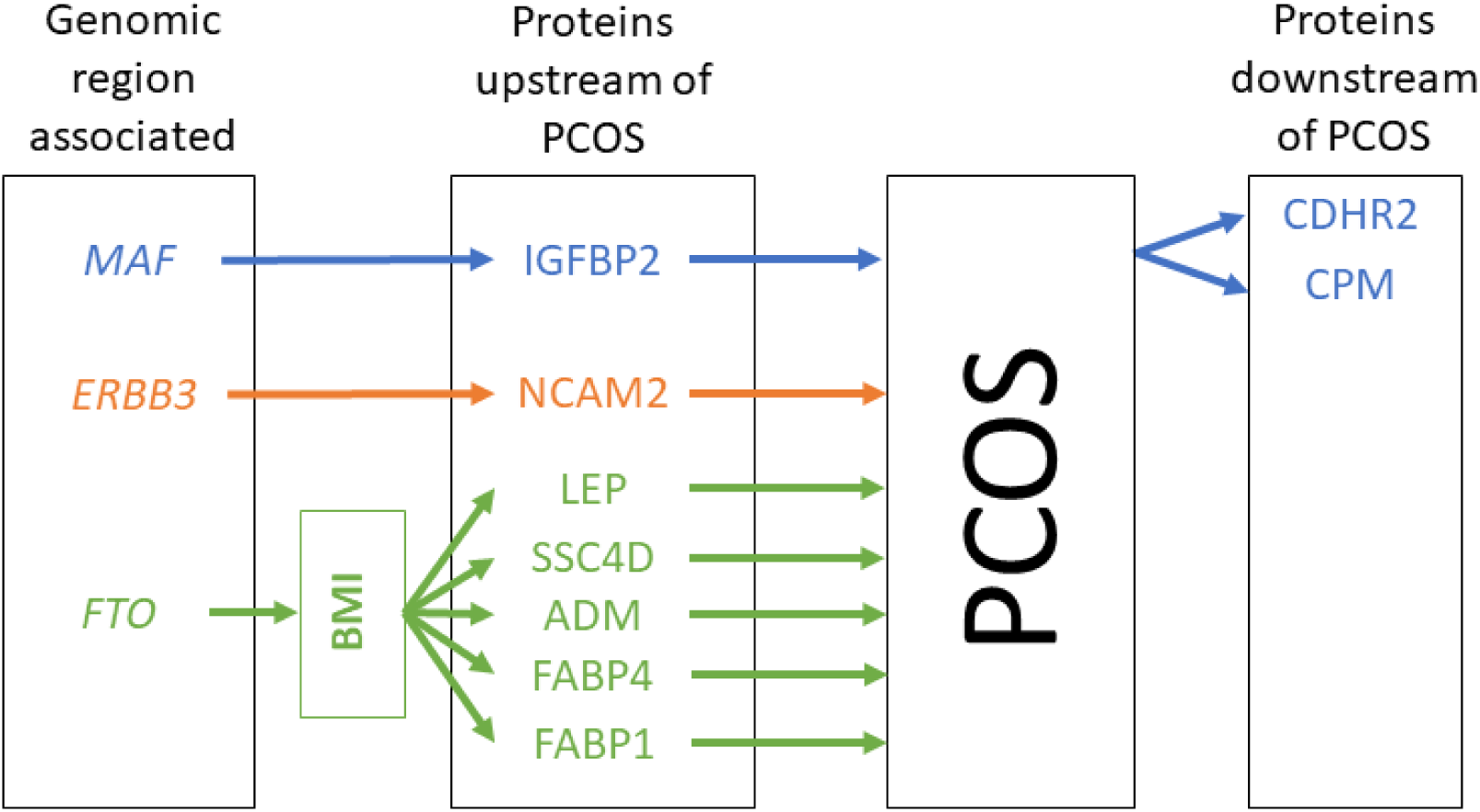
Causal map of the associations between PCOS and E28 (ovarian dysfunction) associated proteins.

### Inferring causal impacts of PCOS on other co-morbidities

The impact of PCOS on fertility, metabolic disease and mental health is well known, but few studies have used a genetic approach to uncover additional comorbidities^43,44^. Previous genetic causal modelling, using Mendelian Randomisation (MR) approaches, have shown that aspects of metabolic syndrome traits are risk factors for PCOS^6^. However, those MR studies did not determine whether PCOS had an effect on broader health status. Therefore, we calculated a polygenic risk score (PRS) for PCOS comprising ∼1.1 million genetic variants to explore the likely causal effect of PCOS on a number of other outcomes.

To identify phenotypes that may share genetic influences with PCOS, we performed polygenic risk score (PRS) based analyses in the UK Biobank study (which was independent of the PCOS GWAS discovery dataset). We used the PRS-CS software to calculate a polygenic risk score for PCOS, which is a Bayesian regression framework that weights the effect size of each variant by the strength of its association (p-value) in the GWAS meta-analysis^45^. To facilitate interpretation, the PRS was standardised; effect sizes (beta or OR) in Table 1 are reported per 1 SD increase in the PCOS PRS. To validate the score, we confirmed that the PCOS PRS was strongly associated with PCOS in women in the UK Biobank (*P*=9×10^-27^) and that the odds of PCOS increased across increasing quintiles of PRS (**Supplementary** Figure 6). As expected, there was also a strong association between a higher PCOS PRS and increased BMI in both women and men (Table 1).

**Table 1.**
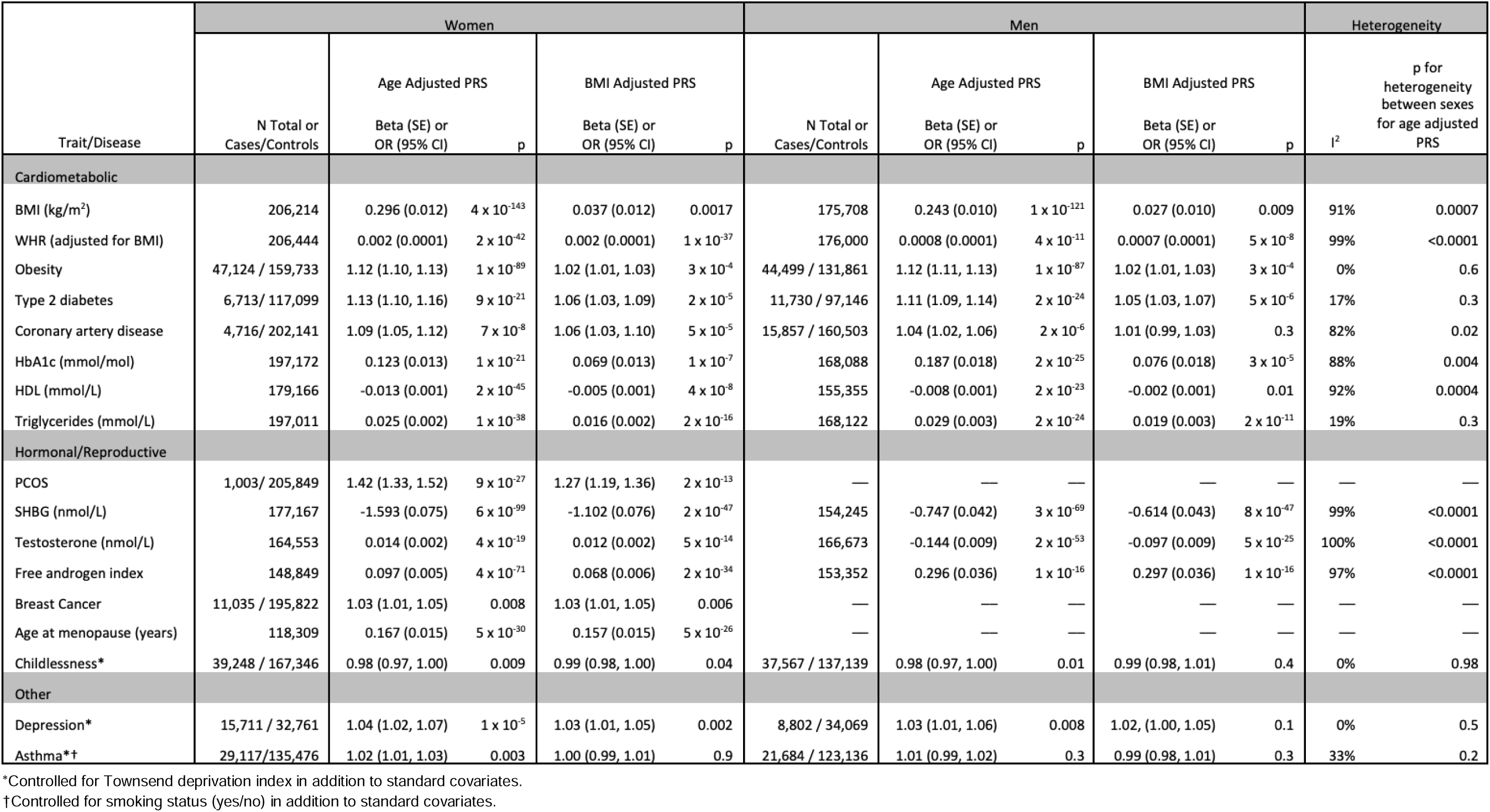
PCOS PRS associations with traits and diseases of interest. We calculated the PCOS PRS in women and men in the UK Biobank. Then, we tested for association between PCOS PRS and phenotypes of interest. We applied Bonferroni correction for multiple testing for association with 16 phenotypes, and hence, associations with p-values < 0.003 are considered statistically significant. Statistically significant heterogeneity between the sexes were considered when *I*^2^>80% and Cochran’s Q *P*-value for heterogeneity<0.004 (0.05/13 tests; BMI: Body mass index, WHR: Waist-to-hip ratio, HbA1c: Hemoglobin A1c, HDL: High-density lipoprotein, SHBG: Sex hormone binding globulin).

Since increased BMI is a risk factor for both PCOS and many of the expected metabolic comorbidities, we assessed the BMI-independent causal relationship between PCOS and metabolic outcomes in two additional analyses: 1) we included measured BMI as a covariate and 2) we generated and tested a second BMI-adjusted PRS for PCOS. The association of the BMI-adjusted PRS with baseline BMI in the UK Biobank was substantially attenuated in women and men (**Table 1** and **Supplementary Table 21**). The PRS analyses were replicated using an additional polygenic risk score tool LDpred2, which provided consistent results (**Supplementary Table 21**).

Both in women and in men, a higher PCOS PRS was associated with increased risk of coronary artery disease (CAD), T2D and obesity (Table 1). It was also associated with a number of cardiometabolic risk factors: higher waist-to-hip ratio adjusted for BMI (WHR_adjBMI_), higher HbA1c, higher triglycerides and lower HDL cholesterol. Regarding hormonal/reproductive risk factors, higher PRS was associated with lower SHBG levels and higher free androgen index (FAI) in both sexes (**Table 1**). The sex-stratified BMI-adjusted model analyses provided support for all these observations.

We then tested for the presence of sex differences in the effects of the PRS on i) cardiometabolic outcomes where different effects of sex were observed for BMI, WHR_adjBMI_, CAD, HbA1c and HDL cholesterol; and ii) hormonal/reproductive outcomes where differential sex-related effects were observed for SHBG, total testosterone and FAI (**Table 1**). There has been increasing interest in the links between reproductive diseases and mental health^46,47^; we found only a nominal association between the PCOS PRS and depression in women after BMI adjustment (**Table 1**).

We used Mendelian Randomisation to provide additional evidence to determine whether highlighted associations were causal (**Supplementary Tables 22 and 23**). While MR might provide more robust causal inference there are a number of caveats: the available outcome data is often in men and women combined, and with only 29 variants this analysis is likely very under-powered. Most of the associations with non-reproductive phenotypes were not significant; including when BMI was the outcome, likely demonstrating that the method is underpowered. Interestingly, the MR analysis showed associations with two non-cardiometabolic outcomes - depression and asthma, with no attenuation when controlling for BMI.

### Pleiotropic effects of PCOS on reproductive outcome

Consistent with the overlap between signals for PCOS and ANM, PCOS susceptibility was associated with later ANM in both PRS (**Table 1**) and MR analyses (Supplementary Table 23). Conversely, we also observed an apparent effect of susceptibility to later ANM on higher PCOS risk (Supplementary Table 23), indicating a bidirectional relationship. Given the importance of DNA repair as a mechanism that regulates ANM, we tested to determine whether the same pathways also contributed to PCOS. We performed MR analyses for PCOS with reported variants for ANM stratified by their likely functions on DNA repair and genes unrelated to DNA repair^7^. Both strata of ANM variants indicated an effect of later ANM on higher risk of PCOS (**Supplementary Table 23**). However, the MR estimated effect of ANM was larger using non-DNA repair ANM variants than when using DNA repair variants (*P*-value for the difference between estimates = 1.5×10^-6^, **Supplementary** Figure 7), which suggests a role for sex hormone-related pathways common to PCOS and ANM. The role of hormone levels as common causal factor is highlighted by the shared signal at *FSHB* associated with lower FSH levels, higher PCOS risk and later age at menopause.

The DNA repair-related ANM variants that are associated likely act via various DNA-repair pathways, which have differing effects on PCOS. For example, ANM variants at *BRCA1* and *BRCA2,* genes involved in homologous recombination repair of double strand DNA breaks, are robustly associated with an earlier ANM but not with PCOS (P=0.94 and P=0.11 respectively). On the other hand, *CHEK2,* known to maintain DNA integrity through checkpoint control^48^, is associated with a later ANM, greater risk of PCOS and higher serum AMH levels in PCOS.

The PCOS PRS showed unexpected nominal associations with lower risk of childlessness, although these were not confirmed in the MR analysis. We therefore examined the impact of PCOS susceptibility on eight infertility phenotypes in a hospital-based cohort, the Copenhagen Hospital Biobank (CHB)^49^ (multiple testing threshold *P*<0.006; = P<0.05/8; **Supplementary Table 24**). A higher PCOS PRS was associated with increased risk of infertility in women (OR=1.03, *P*=0.02 after age adjustment; OR=1.04, *P*=0.00011 after age and BMI adjustment). The stronger association after adjustment for BMI suggests that in addition to affecting fertility via BMI, PCOS also affects fertility through other mechanisms, such as hyperandrogenism, that are independent of BMI.

Interestingly, the PCOS PRS was associated with an increased number of oocytes aspirated during IVF treatment (Beta=0.025, *P*=1×10^-4^ after age adjustment; beta=0.027, *P*=2×10^-5^ after age and BMI adjustment). In separate data from 892 women, two of the 29 loci, at *ZBTB16* and *SHBG*, were associated with larger ovarian volume; which is both a marker of ovarian reserve linked to fertility, and a symptomatic presentation of PCOS. These findings suggest a greater available oocyte pool in PCOS.

There was also support for the hypothesis that some genetic PCOS susceptibility might exhibit a balancing pleiotropy effect on reproductive success. We assessed the links from PCOS to age at first birth^50^, age at last birth^50^, childlessness^51^ and number of children^51^ using publicly available GWAS datasets (**Supplementary Table 23**). There were no apparent associations with childlessness or number of children. The latter result was confirmed in the CHB data, in which there was no association between the PCOS PRS and the completed family size (*P*=0.07) or rates of pregnancy (*P*=0.25; **Supplementary Table 24**). However, there was a nominally significant association with later age at last birth when data were controlled for age and BMI (*P*=0.01). We further assessed the association between PCOS and age at last live birth in the UK Biobank in 1,003 women with PCOS and 205,849 controls. PCOS was associated with later age at live birth (Beta=0.46 years, *P*=0.04 after age adjustment, Beta=0.81 years, *P*=0.0003 after age and BMI adjustment). These age at last birth results could be explained by a longer reproductive window or by a shifting of the window, and the lack of association with childlessness would indicate that some compensatory mechanisms exist.

## Discussion

This study expands the number of PCOS genome-wide significant loci from 19 to 29. The new PCOS locus at *FTO*, well-established for obesity, highlights the link between PCOS, metabolic syndrome and obesity. Other signals at *SHBG*, *FSHR* (associated for the first time in a European GWAS) and the *CYP3* complex highlight hormonal regulation in the aetiology of PCOS. Alongside these results, we also present proteins that are associated with ovarian dysfunction. The protein associations confirm some top candidate genes at GWAS loci (*ERBB3, ERBB4* and *ZBTB16*).

The identified genetic loci underscore the sex hormonal origins of PCOS. Annotation of these signals to consensus genes has identified signals at *FSHB*^52^ and *FSHR*^11^, highlighting the role of pituitary gonadotrophs (LH and FSH) in ovarian follicle stimulation. Other consensus genes, *SHBG*, *INHBB*, *AMH* and *TEX41* - this last robustly associated with AMH levels – also point to hormones related to ovarian folliculogenesis, and feedback on the hypothalamic-pituitary-adrenal axis. Inhibin B is secreted by granulosa cells of the small to large antral follicles and inhibits FSH release, ensuring the growth of one dominant follicle^53,54^. In support, the *INHBB* variant was nominally, inversely associated with FSH levels. Circulating AMH levels reflect the number of small antral follicles and AMH reduces FSH sensitivity of growing follicles^20^. Furthermore, AMH inhibits aromatase activity at the level of a growing follicle, and increases LH-dependent gonadotropin-releasing hormone (GnRH) pulsatility at the hypothalamus^55^. Finally, SHBG is a binding protein for androgens, thereby regulating free and bioavailable androgen levels^56^. In men there is a corresponding increase in free androgen index which explains the connection between PCOS variants and male pattern balding^6^. In summary, PCOS risk is affected by a number of classical and well-established sex hormonal pathways.

A significant proportion of the identified PCOS loci overlap with those associated with age at menopause. In all cases, PCOS risk alleles confer later age at menopause. There are two possible mechanisms which could explain, perhaps in tandem, the links between these two phenotypes. First, several overlapping variants are related to genes linked to DNA repair mechanisms such as *MSH6*, *CHEK2* and *RAD50*. The partitioned Mendelian Randomisation analysis suggested that both DNA damage repair and non-DNA damage repair (thought to be predominantly hormonal pathways) were causal for PCOS, but that the latter had a stronger influence. While age at menopause is thought to be impacted by a range of pathways linked to DNA repair, those signals shared with PCOS might be relate to more specific mechanisms, such as *CHEK2*, where the effect is to have DNA-damaged oocytes persist for longer ^7^. Thus, there may be follicles with DNA-damaged oocytes that remain in the ovary because the DNA checkpoint removal mechanism failed. In PCOS, this may reduce oocyte atresia, leading to continuous AMH expression and thereby stronger inhibition of primordial follicle recruitment (associated with later ANM) and reduced FSH-sensitivity (contributing to the polycystic ovary morphology seen in PCOS)^19,20^. In addition, PCOS was not associated with *BRCA1* or *BRCA2* variants, which appear to influence earlier age at menopause based on less functional DNA-repair mechanisms and potential loss of damaged oocytes^7^.

Secondly, changes in hormonal levels may increase follicle numbers as demonstrated for increased androgen levels^57^ and activin, the product of two INHB subunits^58^. It is also possible that hormone levels reduce depletion of the primordial follicle pool causing a later end to the reproductive window, as has been demonstrated for the variants causing lower FSH levels^59^. Moreover, the variant in *FSHB* was also linked to less follicle selection across the reproductive lifespan potentially leading to a greater follicle pool consistent with PCOS. In addition, increased serum AMH levels, seen in PCOS patients, reduce the rate of primordial follicle recruitment and may thereby slow follicle pool depletion, leading to later menopause^19,20^. In addition, observational studies suggest that women with PCOS or PCOS symptoms have children as often as asymptomatic women and have reproductive success when followed over a sufficiently long term^60–63^. In both our PCOS PRS and epidemiological analysis of women with PCOS in the UK Biobank, there was a suggestion of an impact of a later age of last birth in both the PRS genetic analyses and epidemiological analysis and this effect was also seen in the CHB data. This is consistent with a previous finding that a longer or shifted window of reproduction was required to achieve the same cumulative family size in women with PCOS^63^. The main cause of infertility in PCOS is irregular ovulation, and therefore, the relative infertility at younger ages may be balanced by improved ovulations at later ages^64,65^.

In summary, the new loci contain a number of risk genes expected to increase the follicle complement in PCOS^66,67^. This finding supports the Rotterdam diagnostic criteria for PCOS, which highlight polycystic ovarian morphology (number of growing small antral follicles), hyperandrogenism (hormone regulation) and related irregular ovulation and menses as primary etiologic features of PCOS^2^.

The score-based analyses stressed the link to metabolic diseases, with a number of strong associations between PCOS and clinical endpoints, mirroring observed associations^68^, and previous studies^10,43^. Unsurprisingly, a higher PCOS PRS was associated with higher BMI and increased risk of obesity in both women and men; thus, undoubtedly, much of the effect on cardio-metabolic diseases seen in the BMI unadjusted models is via the “common soil” impact of BMI. However, many of the associations remained significant in women (though substantially attenuated), and not in men after controlling for BMI. These female-specific effects, that were not BMI-related, suggest a shared causal factor between PCOS and metabolic disease. A plausible mechanism explaining this is the higher androgen levels in PCOS, which have been shown to be a risk factor for CAD^25,69,70^. Although testosterone and other androgens decrease to the same level as in controls after menopause, the continued lower SHBG in women with PCOS after menopause and the lasting impact of androgens during reproductive age on physiology may result in long term increased CAD risk^71,72^.

In addition to this, the proteins associated with reproductive dysfunction stress the links between reproductive phenotypes and the metabolic syndrome. Associations were seen with classical adiposity and metabolic proteins including leptin and furin. Other associated proteins are vital to cholesterol metabolism such as PCSK9 and the LDL receptor, which are important for both cardiovascular risk and steroidogenesis. There were also proteins that contribute to the metabolic response to a high fat diet. It is important to consider that most of the women in whom the protein-based analysis was done were assessed after the end of their reproductive window. The data again implicate the lower SHBG and higher free androgen levels in PCOS after menopause^71^, and potentially the sustained effects of hyperandrogenism even after the reproductive years. Thus, these results and the score-based analyses together suggest that there is an ongoing, adverse pattern of cardiometabolic health in women with a genetic risk for PCOS.

## Conclusions

Here we identify genetic regions and proteins associated with PCOS. In general, the genomic loci appear to primarily implicate hormonal pathways as the causal factors for PCOS; while the proteins stress the common causal factors between PCOS and metabolic disease, particularly pathways related to increased BMI. Our findings highlight important links between PCOS and type 2 diabetes and coronary artery disease, via mechanisms that are related to and also independent of adiposity. We also expand our understanding of the factors affecting the ovarian follicle complement on the condition, including both hormonal influences and specific DNA repair mechanisms, and their role in PCOS. We also demonstrate some evidence of balanced pleiotropy conferred by PCOS genetic susceptibility that maintains the high prevalence of PCOS in the population.

## Methods

### Data Collection and Quality Control

Summary results of genome-wide association analysis (GWAS) using a case-control setting were provided by the studies contributing to the meta-analysis. At the study level, the analyses were adjusted for age, principal components and body mass index (BMI, only for BMI-adjusted analyses). Central quality control (QC) was performed by two independent analysts using the EasyQC pipeline^73^. Variant exclusion filters used included: (1) Minor allele frequency (MAF) <1%, (2) imputation quality (*R*^2^) <0.3 or info <0.4 for MACH and IMPUTE2, respectively^73^.

### Meta-analysis

A fixed-effect, inverse-weighted-variance meta-analysis approach was used with the collected summary statistics from the individual studies. Either GWAMA^74^ or METAL^75^ were employed as meta-analysis tools. We performed meta-analyses for all ancestries combined and only European ancestry combined. These meta-analyses were carried out using two models; age-adjusted, and age and BMI-adjusted, given the association between obesity and PCOS^5^. Variants present in at least three strata were reported and used in further analyses.

These meta-analysis results were then combined with the previously published genome-wide meta-analysis summary statistics^6^ to increase the statistical power and discover further associations with PCOS status. We called this analysis *the 2-strata meta-analysis*. As previous research had found no substantial heterogeneity in variant discovery as a function of different diagnostic criteria^6^, studies with any method of case ascertainment were combined. Variants present in all strata were reported and used in the follow-up analyses. Identified variants were annotated and investigated further with regards to their biological function using FUMA^76^. Forest plots for comparing the effect sizes across the strata in the meta-analysis were made using the ggplot2 package in R.

Furthermore, we compared the effect sizes across different phenotype definitions used; PCOS definitions based on i) electronic health records (EHRs), ii) clinical diagnosis and iii) self-reports were included in this comparison. Additional meta-analysis was performed to statistically test for heterogeneity across these three PCOS definitions. In addition to visually inspecting forest plots for the meta-analysis, Cochrane’s Q *P*-value and *I*^2^ were used for assessing heterogeneity.

The summary statistics from the age-adjusted meta-analyses were further combined with the previously published summary statistics from 23andMe, Inc. in order to increase the statistical power^6^. We called this analysis *the 3-strata meta-analysis*. The resulting sample size was 545,541 (21,570 cases and 523,971 controls).

We assessed the lead GWAS variants (*p*<5x10^-8^) by examining their relationship with 20 related metabolic, hormonal and reproductive phenotypes with available GWAS results data. Except for LH and FSH all other traits were publicly available (**Supplementary Table 10**). The heatmap was drawn using the “pheatmap” library in R 3.6.1.

### Fine-mapping

To identify a credible set of variants containing the most likely causal variant underlying our association signals, we conducted fine-mapping using the shotgun stochastic search method as performed in FINEMAP^77^. We used summary statistics from our two-stage summary GWAS meta-analysis results without the data from 23andMe, Inc., and considered all variants within 1 MB +/- from our tag variants. We used two different contributing studies as LD references to perform fine-mapping. First, we used unrelated (up to 2nd degree), European ancestry women from the MyCode Community Health Initiative Study (DiscovEHR) (N=47,061) with genetic data imputed to the 1000 Genomes Phase III global reference panel. European ancestry was inferred using genetic data as described elsewhere^78^. Second, we used an unrelated dataset of European ancestry females (N=36,890) in the EHR-linked biobank at Vanderbilt University Medical Center (BioVU). Genetic data were imputed to the Haplotype Reference Consortium and European ancestry was defined by principal components^79^. We assumed a single causal variant for all loci, and for four loci with evidence of a secondary signal, we also performed fine-mapping assuming two causal variants.

### Functional Mapping and Annotation of GWAS

Functional mapping and annotation of GWAS was performed with FUMA, and further annotation of the association results with PhenoScanner (date accessed 25 March 2022)^80,81^. FUMA analyses were performed using the summary statistics for: i) the top 29 PCOS-associated variants in the 3-strata meta-analysis; and ii) the genome-wide 2-strata meta-analysis. Unless specified otherwise, the default settings were used in the FUMA analyses for both SNP2GENE and GENE2FUNC^76^.

### Proteomic Analysis

Proteomic analysis using logistic regression for the association of normalised protein levels with disease was run in the ∼22,000 women with data from the Olink panel of plasma proteins, aged 56.5±8.1 yrs^82^. Here the outcome was the first occurrence data, and to maximise sample size we used a diagnosis of any of ICD10 category E28, the supra-group that includes PCOS. Proteins were considered significantly associated if they passed a Bonferroni corrected p-value threshold of 3.4×10^-5^. The generation of the protein data is described elsewhere^83^.

Separately, we performed a protein Phewas for each of the variants that we identified in the GWAS meta-analysis using the total sample of ∼44,000 (men and women) with Olink data to identify proteins linked to our PCOS signals. Again, we used a Bonferroni corrected p-value threshold of 3.4×10^-5^. Once this panel of proteins had been identified we then considered if these were also associated with the diagnosis of ICD10 E28. Finally, once we had established our variant-protein pairs, we attempted to establish the position in the causal pathways by considering the relative *R*^2^, with those variants that had a higher *R*^2^ with PCOS suggesting that the protein was downstream of PCOS, and vice-versa.

### GWAS-to-Gene pipeline

The GWAS-to-Genes pipeline leverages a range of annotation methods to highlight likely causal genes at each of the identified signals, as described elsewhere^18^. Briefly, tissue enrichment for GWAS associations was performed using LD score regression to identify key tissues for annotations with tissue specific datasets. Then a gene score is generated from the following panel of annotations: a) The closest gene to the signal with these scored 1.5 points. b) eQTL colocalisation from both SMR-HEIDI and coloc were scored 1.5 points, or 1 if only from one of these. An additional point was given to genes with eQTLs at secondary signals. c) pQTL colocalisation scored the same as for eQTLs. d) Coding variants, with variants of deleterious or damaging predicted consequence in LD with GWAS PCOS signals were scored 1.0 point, or only 0.5 points if the coding variants were predicted to be benign or tolerated. e) Genes targeted by enhancers which overlapped with or were correlated with GWAS PCOS signals were scored 1.0 point. f) PoPs prioritised genes at each locus were scored 1.5 points^84^.

### Gene-set Enrichment Analysis

To perform gene-set enrichment analysis that leveraged information across both the proteomics and the implicated genes we used GProfiler selecting either the consensus gene or the associated protein from the proteomics analysis. Clustering of the pathways was done using an index of dissimilarity based on the shared genes across the enriched intersections of each pathway^18^.

### Polygenic Risk Score Analyses in the UK Biobank

We used the PRS-CS software to calculate a polygenic risk score (PRS) for PCOS, which is a Bayesian regression framework that applies continuous shrinkage parameters to estimate posterior effect sizes^45^. This work was performed in the UK Biobank, a population-based cohort of ∼500,000 individuals in the United Kingdom^85^, which was independent of the discovery GWAS sample. The tuning or global shrinkage parameter phi=1×10^-4^ that optimised the association of the PRS for PCOS in the UK Biobank as previously reported was used^10^. Using this method, our PCOS polygenic risk score included 1,119,009 genetic variants. In the same UK Biobank sample, we replicated these analyses using another PRS tool, LDpred-2, which employs Bayesian shrinkage model^86^.

To identify women with PCOS in the UK Biobank study, data from self-report, primary-care clinical events, and/or ICD 9 and ICD 10, as previously reported^10^ was used. We binned women with and without a diagnosis of PCOS by their quintile of PRS and used logistic regression to determine the odds of PCOS for each quintile using the lowest quintile as a reference. Women with a PCOS PRS in the highest quintile had an increased odds of PCOS (OR 2.41, 95% CI 1.96-2.98; *P*=2×10^-16^). Thus, our PCOS PRS is able to represent the genetic risk for PCOS in women in the UK Biobank.

Ascertainment of cardiometabolic and androgenic phenotypes have been previously reported^10^. All other phenotypes, including measures of fertility and longevity, asthma, and mental health disorders were based on a composite of self-reported measures, diagnosis codes from hospitalisation records, and age at diagnosis (**Supplementary Information**). We used linear and logistic regression to analyse the associations between continuous and dichotomous phenotypes and the PCOS PRS, respectively. We adjusted all analyses for age, age squared, genotyping array, the UK Biobank assessment centre, and the first ten genetic principal components; for asthma and psychological outcomes, we additionally controlled for the Townsend Deprivation Index, and for asthma, we further controlled for smoking status. Adjustment for BMI was performed in two ways, first a measured BMI was included in the model as a covariate; second we constructed a score based on the GWAS meta-analysis where the genetic associations were adjusted for BMI.

### Polygenic Risk Score Analysis in Copenhagen Hospital Biobank based on the Danish Registries

Polygenic Risk Scores (PRS) for PCOS were calculated using LDPred2^87^. These genome-wide scores were calculated using the meta-analysis excluding data from 23andMe Inc. Autosomal genotype data from 138,669 individuals in the Copenhagen Hospital Biobank (CHB) were filtered to only include variants present in a set of 1,054,330 reference variants recommended by LDPred2 developers. Missing genotype information was imputed to be the reference allele for the affected locus. GWAS summary statistics were pre-processed with MungeSumStats.

The completed family size was determined by counting the number of live births from the Medical Birth Registry (MBR)^88^. This study was initiated in 1973, and data is considered complete. Only women born in the years 1957-1973 were included in this analysis, thus the youngest would be 45 years old, and 61 years old when data collection ended (31st December 2018). Data were treated as count data, and we tested to determine whether there was equi-, under-, or over-dispersion using the AER R package (v1.2.10). We found significant underdispersion (dispersion=0.60, p<2.2×10^-16^). Consequently, data were analysed using a Conway-Maxwell Poisson distribution.

From the MBR, we also identified the age at first birth and last birth (expressed in days). Data were analysed using a linear regression, and model fit was inspected from residuals. There were no signs of deviation from a Gaussian error.

The Danish IVF registry was initiated in 1994, and contains all treatments and procedures related to medically assisted reproduction. Reporting is mandatory for both private and public clinics. Furthermore, there is information on any treatment related to the procedure, and treatment duration. Female infertility was defined using the 628 (ICD8) and N97 (ICD10; excluding N97.4) in the National Patient Registry (public hospitals only^89^) and “female cause” in the IVF Register. Male infertility was defined using the 606 (ICD8) and N46 (ICD10) in the National Patient Registry and “male cause” (excluding male infertility due to sterilization) in the IVF Register. For number of oocytes, we extracted all aspirations performed in the period from 18th January 1994 - 31st December 2018. The mandatory reported data were changed in 2005, and thus, we analysed the two time periods (1994-2005 and 2006-2018) separately and meta-analysed. We additionally extracted information on treatment (Klomifen, HMG-FSH, GNRH-A, Oestrogen, Progesterone, HCG) and the number of treatment days prior to the aspiration. Both time periods were over dispersed and were analysed using a negative binomial distribution. To take into account multiple aspirations for a single woman, we included an individual random term.

Lastly, we investigated the number of cycles before a woman got pregnant or ceased treatment. Each woman was only included until the first pregnancy. All transfer or insemination attempts were summarised, and analysed using a Conway-Maxwell Poisson distribution, as data were under dispersed. Furthermore, a term for zero-inflated was also tested as only 81% of the population achieved a pregnancy. A model that included a zero-inflation term was found to fit significantly better (*p*<2.2×10^-16^, likelihood-ratio test). This was also substantiated by lower AIC and BIC scores.

All models were fit using glmmTMB, and no rate models, except the number of cycles until pregnant, had indications of zero-inflation.

GWAS Catalog Accessions for calculated bioavailable testosterone, total testosterone and SHBG: GCST90012102, GCST90012106 and GCST90012112.

### Mendelian Randomisation Analysis

Mendelian Randomisation analysis was performed using two sample inverse weighted methods (IVW)^90^. In addition, the intercept from the MR-EGGER^91^ was calculated to provide a test of directional pleiotropy and the I^2^ metric to assess general heterogeneity of the variants. Data for the outcomes was taken from the most recent genome-wide study for each of the outcomes (thus in most cases this data were not sex-specific). To correct for any impact of the role of BMI on analyses multivariate IVW^92^ was implemented. The betas for these variants to BMI was taken from the most recent GIANT consortium study which combined GWAS meta-analysis data with that from UK Biobank^93^. For the analysis of association between menopause variants and PCOS split by evidence for a DNA damage effect, the variants were classified based on the proximity to a known DNA damage repair gene as per Ruth *et al.*^7^.

## Data Availability

Data produced will either be available online or upon reasonable request to the authors after peer-reviewed publication of the manuscript.

## Acknowledgements

This work was conducted using UK Biobank, application numbers 9905, and 31823. We would like to thank the research participants and employees of 23andMe Inc. for making this work possible. Tugce Karaderi is supported by the Novo Nordisk Foundation Data Science Investigator grant (NNF20OC0062294). Further Acknowledgements can be found in the Supplement.

## References

1. Knochenhauer ES, Key TJ, Kahsar-Miller M, Waggoner W, Boots LR, Azziz R. Prevalence of the polycystic ovary syndrome in unselected black and white women of the southeastern United States: a prospective study. J Clin Endocrinol Metab 1998;83:3078–82.

2. Rotterdam ESHRE/ASRM-Sponsored PCOS consensus workshop group. Revised 2003 consensus on diagnostic criteria and long-term health risks related to polycystic ovary syndrome (PCOS). Hum Reprod 2004;19:41–7.

3. Ehrmann DA, Barnes RB, Rosenfield RL, Cavaghan MK, Imperial J. Prevalence of impaired glucose tolerance and diabetes in women with polycystic ovary syndrome. Diabetes Care 1999;22:141–6.

4. Legro RS, Kunselman AR, Dodson WC, Dunaif A. Prevalence and predictors of risk for type 2 diabetes mellitus and impaired glucose tolerance in polycystic ovary syndrome: a prospective, controlled study in 254 affected women. J Clin Endocrinol Metab 1999;84:165–9.

5. Day FR, Hinds DA, Tung JY, Stolk L, Styrkarsdottir U, Saxena R, Bjonnes A, Broer L, Dunger DB, Halldorsson BV, Lawlor DA, Laval G, Mathieson I, McCardle WL, Louwers Y, Meun C, Ring S, Scott RA, Sulem P, Uitterlinden AG, Wareham NJ, Thorsteinsdottir U, Welt C, Stefansson K, Laven JS, Ong KK, Perry JR. Causal mechanisms and balancing selection inferred from genetic associations with polycystic ovary syndrome. Nat Commun 2015;6:8464.

6. Day F, Karaderi T, Jones MR, Meun C, He C, Drong A, Kraft P, Lin N, Huang H, Broer L, Magi R, Saxena R, Laisk T, Urbanek M, Hayes MG, Thorleifsson G, Fernandez-Tajes J, Mahajan A, Mullin BH, Stuckey BGA, Spector TD, Wilson SG, Goodarzi MO, Davis L, Obermayer-Pietsch B, Uitterlinden AG, Anttila V, Neale BM, Jarvelin MR, Fauser B, Kowalska I, Visser JA, Andersen M, Ong K, Stener-Victorin E, Ehrmann D, Legro RS, Salumets A, McCarthy MI, Morin-Papunen L, Thorsteinsdottir U, Stefansson K, and Me Research T, Styrkarsdottir U, Perry JRB, Dunaif A, Laven J, Franks S, Lindgren CM, Welt CK. Large-scale genome-wide meta-analysis of polycystic ovary syndrome suggests shared genetic architecture for different diagnosis criteria. PLoS Genet 2018;14:e1007813.

7. Ruth KS, Day FR, Hussain J, Martinez-Marchal A, Aiken CE, Azad A, Thompson DJ, Knoblochova L, Abe H, Tarry-Adkins JL, Gonzalez JM, Fontanillas P, Claringbould A, Bakker OB, Sulem P, Walters RG, Terao C, Turon S, Horikoshi M, Lin K, Onland-Moret NC, Sankar A, Hertz EPT, Timshel PN, Shukla V, Borup R, Olsen KW, Aguilera P, Ferrer-Roda M, Huang Y, Stankovic S, Timmers P, Ahearn TU, Alizadeh BZ, Naderi E, Andrulis IL, Arnold AM, Aronson KJ, Augustinsson A, Bandinelli S, Barbieri CM, Beaumont RN, Becher H, Beckmann MW, Benonisdottir S, Bergmann S, Bochud M, Boerwinkle E, Bojesen SE, Bolla MK, Boomsma DI, Bowker N, Brody JA, Broer L, Buring JE, Campbell A, Campbell H, Castelao JE, Catamo E, Chanock SJ, Chenevix-Trench G, Ciullo M, Corre T, Couch FJ, Cox A, Crisponi L, Cross SS, Cucca F, Czene K, Smith GD, de Geus E, de Mutsert R, De Vivo I, Demerath EW, Dennis J, Dunning AM, Dwek M, Eriksson M, Esko T, Fasching PA, Faul JD, Ferrucci L, Franceschini N, Frayling TM, Gago-Dominguez M, Mezzavilla M, Garcia-Closas M, Gieger C, Giles GG, Grallert H, Gudbjartsson DF, Gudnason V, Guenel P, Haiman CA, Hakansson N, Hall P, Hayward C, He C, He W, Heiss G, Hoffding MK, Hopper JL, Hottenga JJ, Hu F, Hunter D, Ikram MA, Jackson RD, Joaquim MDR, John EM, Joshi PK, Karasik D, Kardia SLR, Kartsonaki C, Karlsson R, Kitahara CM, Kolcic I, Kooperberg C, Kraft P, Kurian AW, Kutalik Z, La Bianca M, LaChance G, Langenberg C, Launer LJ, Laven JSE, Lawlor DA, Le Marchand L, Li J, Lindblom A, Lindstrom S, Lindstrom T, Linet M, Liu Y, Liu S, Luan J, Magi R, Magnusson PKE, Mangino M, Mannermaa A, Marco B, Marten J, Martin NG, Mbarek H, McKnight B, Medland SE, Meisinger C, Meitinger T, Menni C, Metspalu A, Milani L, Milne RL, Montgomery GW, Mook-Kanamori DO, Mulas A, Mulligan AM, Murray A, Nalls MA, Newman A, Noordam R, Nutile T, Nyholt DR, Olshan AF, Olsson H, Painter JN, Patel AV, Pedersen NL, Perjakova N, Peters A, Peters U, Pharoah PDP, Polasek O, Porcu E, Psaty BM, Rahman I, Rennert G, Rennert HS, Ridker PM, Ring SM, Robino A, Rose LM, Rosendaal FR, Rossouw J, Rudan I, Rueedi R, Ruggiero D, Sala CF, Saloustros E, Sandler DP, Sanna S, Sawyer EJ, Sarnowski C, Schlessinger D, Schmidt MK, Schoemaker MJ, Schraut KE, Scott C, Shekari S, Shrikhande A, Smith AV, Smith BH, Smith JA, Sorice R, Southey MC, Spector TD, Spinelli JJ, Stampfer M, Stockl D, van Meurs JBJ, Strauch K, Styrkarsdottir U, Swerdlow AJ, Tanaka T, Teras LR, Teumer A, Thornorsteinsdottir U, Timpson NJ, Toniolo D, Traglia M, Troester MA, Truong T, Tyrrell J, Uitterlinden AG, Ulivi S, Vachon CM, Vitart V, Volker U, Vollenweider P, Volzke H, Wang Q, Wareham NJ, Weinberg CR, Weir DR, Wilcox AN, van Dijk KW, Willemsen G, Wilson JF, Wolffenbuttel BHR, Wolk A, Wood AR, Zhao W, Zygmunt M, Biobank-based Integrative Omics Study C, e QC, Biobank Japan P, China Kadoorie Biobank Collaborative G, kConFab I, LifeLines Cohort S, InterAct c, andMe Research T, Chen Z, Li L, Franke L, Burgess S, Deelen P, Pers TH, Grondahl ML, Andersen CY, Pujol A, Lopez-Contreras AJ, Daniel JA, Stefansson K, Chang-Claude J, van der Schouw YT, Lunetta KL, Chasman DI, Easton DF, Visser JA, Ozanne SE, Namekawa SH, Solc P, Murabito JM, Ong KK, Hoffmann ER, Murray A, Roig I, Perry JRB. Genetic insights into biological mechanisms governing human ovarian ageing. Nature 2021;596:393–7.

8. Vink JM, Sadrzadeh S, Lambalk CB, Boomsma DI. Heritability of polycystic ovary syndrome in a Dutch twin-family study. J Clin Endocrinol Metab 2006;91:2100–4.

9. Zhu T, Goodarzi MO. Causes and Consequences of Polycystic Ovary Syndrome: Insights From Mendelian Randomization. J Clin Endocrinol Metab 2022;107:e899–e911.

10. Zhu J, Pujol-Gualdo N, Wittemans LBL, Lindgren CM, Laisk T, Hirschhorn JN, Chan YM. Evidence From Men for Ovary-independent Effects of Genetic Risk Factors for Polycystic Ovary Syndrome. J Clin Endocrinol Metab 2022;107:e1577–e87.

11. Shi Y, Zhao H, Shi Y, Cao Y, Yang D, Li Z, Zhang B, Liang X, Li T, Chen J, Shen J, Zhao J, You L, Gao X, Zhu D, Zhao X, Yan Y, Qin Y, Li W, Yan J, Wang Q, Zhao J, Geng L, Ma J, Zhao Y, He G, Zhang A, Zou S, Yang A, Liu J, Li W, Li B, Wan C, Qin Y, Shi J, Yang J, Jiang H, Xu JE, Qi X, Sun Y, Zhang Y, Hao C, Ju X, Zhao D, Ren CE, Li X, Zhang W, Zhang Y, Zhang J, Wu D, Zhang C, He L, Chen ZJ. Genome-wide association study identifies eight new risk loci for polycystic ovary syndrome. Nat Genet 2012;44:1020–5.

12. Hayes MG, Urbanek M, Ehrmann DA, Armstrong LL, Lee JY, Sisk R, Karaderi T, Barber TM, McCarthy MI, Franks S, Lindgren CM, Welt CK, Diamanti-Kandarakis E, Panidis D, Goodarzi MO, Azziz R, Zhang Y, James RG, Olivier M, Kissebah AH, Reproductive Medicine N, Stener-Victorin E, Legro RS, Dunaif A. Genome-wide association of polycystic ovary syndrome implicates alterations in gonadotropin secretion in European ancestry populations. Nat Commun 2015;6:7502.

13. Tyrmi JS, Arffman RK, Pujol-Gualdo N, Kurra V, Morin-Papunen L, Sliz E, FinnGen Consortium, Estonian Biobank Research Team, Piltonen TT, Laisk T, Kettunen J, Laivuori H. Leveraging Northern European population history: novel low-frequency variants for polycystic ovary syndrome. Hum Reprod 2022;37:352–65.

14. Barber TM, Bennett AJ, Groves CJ, Sovio U, Ruokonen A, Martikainen H, Pouta A, Hartikainen AL, Elliott P, Lindgren CM, Freathy RM, Koch K, Ouwehand WH, Karpe F, Conway GS, Wass JA, Jarvelin MR, Franks S, McCarthy MI. Association of variants in the fat mass and obesity associated (FTO) gene with polycystic ovary syndrome. Diabetologia 2008;51:1153–8.

15. Brower MA, Hai Y, Jones MR, Guo X, Chen YI, Rotter JI, Krauss RM, Legro RS, Azziz R, Goodarzi MO. Bidirectional Mendelian randomization to explore the causal relationships between body mass index and polycystic ovary syndrome. Hum Reprod 2019;34:127–36.

16. Semple RK, Savage DB, Cochran EK, Gorden P, O’Rahilly S. Genetic syndromes of severe insulin resistance. Endo Rev 2011;32:498–514.

17. Urbanek M, Legro RS, Driscoll DA, Azziz R, Ehrmann DA, Norman RJ, Strauss JF, 3rd, Spielman RS, Dunaif A. Thirty-seven candidate genes for polycystic ovary syndrome: strongest evidence for linkage is with follistatin. Proc Natl Acad Sci USA 1999;96:8573–8.

18. Kentistou KA, Kaisinger LR, Stankovic S, Vaudel M, de Oliveira EM, Messina A, Walters RG, Liu X, Busch AS, Helgason H, Thompson DJ, Santon F, Petricek KM, Zouaghi Y, Huang-Doran I, Gudbjartsson DF, Bratland E, Lin K, Gardner EJ, Zhao Y, Jia R, Terao C, Riggan M, Bolla MK, Yazdanpanah M, Yazdanpanah N, Bradfield JP, Broer L, Campbell A, Chasman DI, Cousminer DL, Franceschini N, Franke LH, Girotto G, He C, Jarvelin MR, Joshi PK, Kamatani Y, Karlsson R, Luan J, Lunetta KL, Magi R, Mangino M, Medland SE, Meisinger C, Noordam R, Nutile T, Concas MP, Polasek O, Porcu E, Ring SM, Sala C, Smith AV, Tanaka T, van der Most PJ, Vitart V, Wang CA, Willemsen G, Zygmunt M, Ahearn TU, Andrulis IL, Anton-Culver H, Antoniou AC, Auer PL, Barnes CL, Beckmann MW, Berrington A, Bogdanova NV, Bojesen SE, Brenner H, Buring JE, Canzian F, Chang-Claude J, Couch FJ, Cox A, Crisponi L, Czene K, Daly MB, Demerath EW, Dennis J, Devilee P, De Vivo I, Dork T, Dunning AM, Dwek M, Eriksson JG, Fasching PA, Fernandez-Rhodes L, Ferreli L, Fletcher O, Gago-Dominguez M, Garcia-Closas M, Garcia-Saenz JA, Gonzalez-Neira A, Grallert H, Guenel P, Haiman CA, Hall P, Hamann U, Hakonarson H, Hart RJ, Hickey M, Hooning MJ, Hoppe R, Hopper JL, Hottenga JJ, Hu FB, Hubner H, Hunter DJ, Investigators A, Jernstrom H, John EM, Karasik D, Khusnutdinova EK, Kristensen VN, Lacey JV, Lambrechts D, Launer LJ, Lind PA, Lindblom A, Magnusson PK, Mannermaa A, McCarthy MI, Meitinger T, Menni C, Michailidou K, Millwood IY, Milne RL, Montgomery GW, Nevanlinna H, Nolte IM, Nyholt DR, Obi N, O’Brien KM, Offit K, Oldehinkel AJ, Ostrowski SR, Palotie A, Pedersen OB, Peters A, Pianigiani G, Plaseska-Karanfilska D, Pouta A, Pozarickij A, Radice P, Rennert G, Rosendaal FR, Ruggiero D, Saloustros E, Sandler DP, Schipf S, Schmidt CO, Schmidt MK, Small K, Spedicati B, Stampfer M, Stone J, Tamimi RM, Teras LR, Tikkanen E, Turman C, Vachon CM, Wang Q, Winqvist R, Wolk A, Zemel BS, Zheng W, van Dijk KW, Alizadeh BZ, Bandinelli S, Boerwinkle E, Boomsma DI, Ciullo M, Chenevix-Trench G, Cucca F, Esko T, Gieger C, Grant SF, Gudnason V, Hayward C, Kolcic I, Kraft P, Lawlor DA, Martin NG, Nohr EA, Pedersen NL, Pennell CE, Ridker PM, Robino A, Snieder H, Sovio U, Spector TD, Stockl D, Sudlow C, Timpson NJ, Toniolo D, Uitterlinden A, Ulivi S, Volzke H, Wareham NJ, Widen E, Wilson JF, Lifelines Cohort S, Danish Blood Donor s, Ovarian Cancer Association C, Breast Cancer Association C, Biobank Japan P, China Kadoorie Biobank Collaborative G, Pharoah PD, Li L, Easton DF, Njolstad P, Sulem P, Murabito JM, Murray A, Manousaki D, Juul A, Erikstrup C, Stefansson K, Horikoshi M, Chen Z, Farooqi IS, Pitteloud N, Johansson S, Day FR, Perry JR, Ong KK. Understanding the genetic complexity of puberty timing across the allele frequency spectrum. medRxiv 2023.

19. Silva MSB, Giacobini P. New insights into anti-Mullerian hormone role in the hypothalamic-pituitary-gonadal axis and neuroendocrine development. Cell Mol Life Sci 2021;78:1–16.

20. Visser JA, Themmen AP. Anti-Mullerian hormone and folliculogenesis. Molec Cell Endocrinol 2005;234:81–6.

21. Verdiesen RMG, van der Schouw YT, van Gils CH, Verschuren WMM, Broekmans FJM, Borges MC, Goncalves Soares AL, Lawlor DA, Eliassen AH, Kraft P, Sandler DP, Harlow SD, Smith JA, Santoro N, Schoemaker MJ, Swerdlow AJ, Murray A, Ruth KS, Onland-Moret NC. Genome-wide association study meta-analysis identifies three novel loci for circulating anti-Mullerian hormone levels in women. Hum Reprod 2022;37:1069–82.

22. Kristensen SG, Kumar A, Kalra B, Pors SE, Botkjaer JA, Mamsen LS, Colmorn LB, Fedder J, Ernst E, Owens LA, Hardy K, Franks S, Andersen CY. Quantitative Differences in TGF-beta Family Members Measured in Small Antral Follicle Fluids From Women With or Without PCO. J Clin Endocrinol Metab 2019;104:6371–84.

23. Day FR, Thompson DJ, Helgason H, Chasman DI, Finucane H, Sulem P, Ruth KS, Whalen S, Sarkar AK, Albrecht E, Altmaier E, Amini M, Barbieri CM, Boutin T, Campbell A, Demerath E, Giri A, He C, Hottenga JJ, Karlsson R, Kolcic I, Loh PR, Lunetta KL, Mangino M, Marco B, McMahon G, Medland SE, Nolte IM, Noordam R, Nutile T, Paternoster L, Perjakova N, Porcu E, Rose LM, Schraut KE, Segre AV, Smith AV, Stolk L, Teumer A, Andrulis IL, Bandinelli S, Beckmann MW, Benitez J, Bergmann S, Bochud M, Boerwinkle E, Bojesen SE, Bolla MK, Brand JS, Brauch H, Brenner H, Broer L, Bruning T, Buring JE, Campbell H, Catamo E, Chanock S, Chenevix-Trench G, Corre T, Couch FJ, Cousminer DL, Cox A, Crisponi L, Czene K, Davey Smith G, de Geus E, de Mutsert R, De Vivo I, Dennis J, Devilee P, Dos-Santos-Silva I, Dunning AM, Eriksson JG, Fasching PA, Fernandez-Rhodes L, Ferrucci L, Flesch-Janys D, Franke L, Gabrielson M, Gandin I, Giles GG, Grallert H, Gudbjartsson DF, Guenel P, Hall P, Hallberg E, Hamann U, Harris TB, Hartman CA, Heiss G, Hooning MJ, Hopper JL, Hu F, Hunter DJ, Ikram MA, Im HK, Jarvelin MR, Joshi PK, Karasik D, Kellis M, Kutalik Z, LaChance G, Lambrechts D, Langenberg C, Launer LJ, Laven JSE, Lenarduzzi S, Li J, Lind PA, Lindstrom S, Liu Y, Luan J, Magi R, Mannermaa A, Mbarek H, McCarthy MI, Meisinger C, Meitinger T, Menni C, Metspalu A, Michailidou K, Milani L, Milne RL, Montgomery GW, Mulligan AM, Nalls MA, Navarro P, Nevanlinna H, Nyholt DR, Oldehinkel AJ, O’Mara TA, Padmanabhan S, Palotie A, Pedersen N, Peters A, Peto J, Pharoah PDP, Pouta A, Radice P, Rahman I, Ring SM, Robino A, Rosendaal FR, Rudan I, Rueedi R, Ruggiero D, Sala CF, Schmidt MK, Scott RA, Shah M, Sorice R, Southey MC, Sovio U, Stampfer M, Steri M, Strauch K, Tanaka T, Tikkanen E, Timpson NJ, Traglia M, Truong T, Tyrer JP, Uitterlinden AG, Edwards DRV, Vitart V, Volker U, Vollenweider P, Wang Q, Widen E, van Dijk KW, Willemsen G, Winqvist R, Wolffenbuttel BHR, Zhao JH, Zoledziewska M, Zygmunt M, Alizadeh BZ, Boomsma DI, Ciullo M, Cucca F, Esko T, Franceschini N, Gieger C, Gudnason V, Hayward C, Kraft P, Lawlor DA, Magnusson PKE, Martin NG, Mook-Kanamori DO, Nohr EA, Polasek O, Porteous D, Price AL, Ridker PM, Snieder H, Spector TD, Stockl D, Toniolo D, Ulivi S, Visser JA, Volzke H, Wareham NJ, Wilson JF, LifeLines Cohort S, InterAct C, kConFab AI, Endometrial Cancer Association C, Ovarian Cancer Association C, consortium P, Spurdle AB, Thorsteindottir U, Pollard KS, Easton DF, Tung JY, Chang-Claude J, Hinds D, Murray A, Murabito JM, Stefansson K, Ong KK, Perry JRB. Genomic analyses identify hundreds of variants associated with age at menarche and support a role for puberty timing in cancer risk. Nat Genet 2017;49:834–41.

24. Mbarek H, Steinberg S, Nyholt DR, Gordon SD, Miller MB, McRae AF, Hottenga JJ, Day FR, Willemsen G, de Geus EJ, Davies GE, Martin HC, Penninx BW, Jansen R, McAloney K, Vink JM, Kaprio J, Plomin R, Spector TD, Magnusson PK, Reversade B, Harris RA, Aagaard K, Kristjansson RP, Olafsson I, Eyjolfsson GI, Sigurdardottir O, Iacono WG, Lambalk CB, Montgomery GW, McGue M, Ong KK, Perry JRB, Martin NG, Stefansson H, Stefansson K, Boomsma DI. Identification of Common Genetic Variants Influencing Spontaneous Dizygotic Twinning and Female Fertility. Am J Hum Genet 2016;98:898–908.

25. Ruth KS, Day FR, Tyrrell J, Thompson DJ, Wood AR, Mahajan A, Beaumont RN, Wittemans L, Martin S, Busch AS, Erzurumluoglu AM, Hollis B, O’Mara TA, Endometrial Cancer Association C, McCarthy MI, Langenberg C, Easton DF, Wareham NJ, Burgess S, Murray A, Ong KK, Frayling TM, Perry JRB. Using human genetics to understand the disease impacts of testosterone in men and women. Nat Med 2020;26:252–8.

26. Cooper LA, Page ST, Amory JK, Anawalt BD, Matsumoto AM. The association of obesity with sex hormone-binding globulin is stronger than the association with ageing--implications for the interpretation of total testosterone measurements. Clin Endocrinol 2015;83:828–33.

27. Aoyama T, Yamano S, Waxman DJ, Lapenson DP, Meyer UA, Fischer V, Tyndale R, Inaba T, Kalow W, Gelboin HV, et al. Cytochrome P-450 hPCN3, a novel cytochrome P-450 IIIA gene product that is differentially expressed in adult human liver. cDNA and deduced amino acid sequence and distinct specificities of cDNA-expressed hPCN1 and hPCN3 for the metabolism of steroid hormones and cyclosporine. J Biol Chem 1989;264:10388–95.

28. Hashimoto M, Kobayashi K, Yamazaki M, Kazuki Y, Takehara S, Oshimura M, Chiba K. Cyp3a deficiency enhances androgen receptor activity and cholesterol synthesis in the mouse prostate. J Steroid Biochem Mol Biol 2016;163:121–8.

29. Magoffin DA, Weitsman SR. Differentiation of ovarian theca-interstitial cells in vitro: regulation of 17 alpha-hydroxylase messenger ribonucleic acid expression by luteinizing hormone and insulin-like growth factor-I. Endocrinol 1993;132:1945–51.

30. Ruth KS, Campbell PJ, Chew S, Lim EM, Hadlow N, Stuckey BG, Brown SJ, Feenstra B, Joseph J, Surdulescu GL, Zheng HF, Richards JB, Murray A, Spector TD, Wilson SG, Perry JR. Genome-wide association study with 1000 genomes imputation identifies signals for nine sex hormone-related phenotypes. Eur J Hum Genet 2015;24:284–90.

31. Saxena R, Georgopoulos NA, Braaten TJ, Bjonnes AC, Koika V, Panidis D, Welt CK. Han Chinese polycystic ovary syndrome risk variants in women of European ancestry: relationship to FSH levels and glucose tolerance. Hum Reprod 2015;30:1454–9.

32. Koppaka V, Thompson DC, Chen Y, Ellermann M, Nicolaou KC, Juvonen RO, Petersen D, Deitrich RA, Hurley TD, Vasiliou V. Aldehyde dehydrogenase inhibitors: a comprehensive review of the pharmacology, mechanism of action, substrate specificity, and clinical application. Pharmacol Rev 2012;64:520–39.

33. Pares X, Farres J, Kedishvili N, Duester G. Medium- and short-chain dehydrogenase/reductase gene and protein families : Medium-chain and short-chain dehydrogenases/reductases in retinoid metabolism. Cell Mol Life Sci 2008;65:3936–49.

34. Kunutsor SK, Abbasi A, Adler AI. Gamma-glutamyl transferase and risk of type II diabetes: an updated systematic review and dose-response meta-analysis. Ann Epidemiol 2014;24:809–16.

35. Cortese A, Zhu Y, Rebelo AP, Negri S, Courel S, Abreu L, Bacon CJ, Bai Y, Bis-Brewer DM, Bugiardini E, Buglo E, Danzi MC, Feely SME, Athanasiou-Fragkouli A, Haridy NA, Inherited Neuropathy C, Isasi R, Khan A, Laura M, Magri S, Pipis M, Pisciotta C, Powell E, Rossor AM, Saveri P, Sowden JE, Tozza S, Vandrovcova J, Dallman J, Grignani E, Marchioni E, Scherer SS, Tang B, Lin Z, Al-Ajmi A, Schule R, Synofzik M, Maisonobe T, Stojkovic T, Auer-Grumbach M, Abdelhamed MA, Hamed SA, Zhang R, Manganelli F, Santoro L, Taroni F, Pareyson D, Houlden H, Herrmann DN, Reilly MM, Shy ME, Zhai RG, Zuchner S. Biallelic mutations in SORD cause a common and potentially treatable hereditary neuropathy with implications for diabetes. Nat Genet 2020;52:473–81.

36. Johansson AS, Mannervik B. Human glutathione transferase A3-3, a highly efficient catalyst of double-bond isomerization in the biosynthetic pathway of steroid hormones. J Biol Chem 2001;276:33061–5.

37. Sawyer L. beta-Lactoglobulin and Glycodelin: Two Sides of the Same Coin? Front Physiol 2021;12:678080.

38. Szklarczyk D, Gable AL, Nastou KC, Lyon D, Kirsch R, Pyysalo S, Doncheva NT, Legeay M, Fang T, Bork P, Jensen LJ, von Mering C. The STRING database in 2021: customizable protein-protein networks, and functional characterization of user-uploaded gene/measurement sets. Nucleic Acids Res 2021;49:D605–D12.

39. Wild RA, Rizzo M, Clifton S, Carmina E. Lipid levels in polycystic ovary syndrome: systematic review and meta-analysis. Fertil Steril 2011;95:1073–9 e1-11.

40. Guo F, Gong Z, Fernando T, Zhang L, Zhu X, Shi Y. The Lipid Profiles in Different Characteristics of Women with PCOS and the Interaction Between Dyslipidemia and Metabolic Disorder States: A Retrospective Study in Chinese Population. Front Endocrinol (Lausanne) 2022;13:892125.

41. Mikhaylova IV, Kuulasmaa T, Jaaskelainen J, Voutilainen R. Tumor necrosis factor-alpha regulates steroidogenesis, apoptosis, and cell viability in the human adrenocortical cell line NCI-H295R. Endocrinol 2007;148:386–92.

42. Oh HS, Rutledge J, Nachun D, Palovics R, Abiose O, Moran-Losada P, Channappa D, Urey DY, Kim K, Sung YJ, Wang L, Timsina J, Western D, Liu M, Kohlfeld P, Budde J, Wilson EN, Guen Y, Maurer TM, Haney M, Yang AC, He Z, Greicius MD, Andreasson KI, Sathyan S, Weiss EF, Milman S, Barzilai N, Cruchaga C, Wagner AD, Mormino E, Lehallier B, Henderson VW, Longo FM, Montgomery SB, Wyss-Coray T. Organ aging signatures in the plasma proteome track health and disease. Nature 2023;624:164–72.

43. Joo YY, Actkins K, Pacheco JA, Basile AO, Carroll R, Crosslin DR, Day F, Denny JC, Velez Edwards DR, Hakonarson H, Harley JB, Hebbring SJ, Ho K, Jarvik GP, Jones M, Karaderi T, Mentch FD, Meun C, Namjou B, Pendergrass S, Ritchie MD, Stanaway IB, Urbanek M, Walunas TL, Smith M, Chisholm RL, Kho AN, Davis L, Hayes MG, International PCOS consortium. A Polygenic and Phenotypic Risk Prediction for Polycystic Ovary Syndrome Evaluated by Phenome-Wide Association Studies. J Clin Endocrinol Metab 2020;105:1918–36.

44. Teede HJ, Tay CT, Laven JJE, Dokras A, Moran LJ, Piltonen TT, Costello MF, Boivin J, Redman LM, Boyle JA, Norman RJ, Mousa A, Joham AE. Recommendations From the 2023 International Evidence-based Guideline for the Assessment and Management of Polycystic Ovary Syndrome. J Clin Endocrinol Metab 2023;108:2447–69.

45. Ge T, Chen CY, Ni Y, Feng YA, Smoller JW. Polygenic prediction via Bayesian regression and continuous shrinkage priors. Nat Commun 2019;10:1776.

46. Dokras A, Stener-Victorin E, Yildiz BO, Li R, Ottey S, Shah D, Epperson N, Teede H. Androgen Excess-Polycystic Ovary Syndrome Society: position statement on depression, anxiety, quality of life, and eating disorders in polycystic ovary syndrome. Fertil Steril 2018;109:888–99.

47. Teede HJ, Misso ML, Costello MF, Dokras A, Laven J, Moran L, Piltonen T, Norman RJ, International PN. Recommendations from the international evidence-based guideline for the assessment and management of polycystic ovary syndrome. Hum Reprod 2018;33:1602–18.

48. Bolcun-Filas E, Rinaldi VD, White ME, Schimenti JC. Reversal of female infertility by Chk2 ablation reveals the oocyte DNA damage checkpoint pathway. Science 2014;343:533–6.

49. Sorensen E, Christiansen L, Wilkowski B, Larsen MH, Burgdorf KS, Thorner LW, Nissen J, Pedersen OB, Banasik K, Brunak S, Bundgaard H, Stefansson H, Stefansson K, Melbye M, Ullum H. Data Resource Profile: The Copenhagen Hospital Biobank (CHB). Int J Epidemiol 2021;50:719–20e.

50. Mills MC, Tropf FC, Brazel DM, van Zuydam N, Vaez A, e QC, Consortium B, Human Reproductive Behaviour C, Pers TH, Snieder H, Perry JRB, Ong KK, den Hoed M, Barban N, Day FR. Identification of 371 genetic variants for age at first sex and birth linked to externalising behaviour. Nat Hum Behav 2021;5:1717–30.

51. Mathieson I, Day FR, Barban N, Tropf FC, Brazel DM, e QC, Consortium B, Vaez A, van Zuydam N, Bitarello BD, Gardner EJ, Akimova ET, Azad A, Bergmann S, Bielak LF, Boomsma DI, Bosak K, Brumat M, Buring JE, Cesarini D, Chasman DI, Chavarro JE, Cocca M, Concas MP, Davey Smith G, Davies G, Deary IJ, Esko T, Faul JD, FinnGen S, Franco O, Ganna A, Gaskins AJ, Gelemanovic A, de Geus EJC, Gieger C, Girotto G, Gopinath B, Grabe HJ, Gunderson EP, Hayward C, He C, van Heemst D, Hill WD, Hoffmann ER, Homuth G, Hottenga JJ, Huang H, HyppLnen E, Ikram MA, Jansen R, Johannesson M, Kamali Z, Kardia SLR, Kavousi M, Kifley A, Kiiskinen T, Kraft P, Kuhnel B, Langenberg C, Liew G, Lifelines Cohort S, Lind PA, Luan J, Magi R, Magnusson PKE, Mahajan A, Martin NG, Mbarek H, McCarthy MI, McMahon G, Medland SE, Meitinger T, Metspalu A, Mihailov E, Milani L, Missmer SA, Mitchell P, Mollegaard S, Mook-Kanamori DO, Morgan A, van der Most PJ, de Mutsert R, Nauck M, Nolte IM, Noordam R, Penninx B, Peters A, Peyser PA, Polasek O, Power C, Pribisalic A, Redmond P, Rich-Edwards JW, Ridker PM, Rietveld CA, Ring SM, Rose LM, Rueedi R, Shukla V, Smith JA, Stankovic S, Stefansson K, Stockl D, Strauch K, Swertz MA, Teumer A, Thorleifsson G, Thorsteinsdottir U, Thurik AR, Timpson NJ, Turman C, Uitterlinden AG, Waldenberger M, Wareham NJ, Weir DR, Willemsen G, Zhao JH, Zhao W, Zhao Y, Snieder H, den Hoed M, Ong KK, Mills MC, Perry JRB. Genome-wide analysis identifies genetic effects on reproductive success and ongoing natural selection at the FADS locus. Nat Hum Behav 2023;7:790–801.

52. Censin JC, Bovijn J, Holmes MV, Lindgren CM. Colocalization analysis of polycystic ovary syndrome to identify potential disease-mediating genes and proteins. Eur J Hum Genet 2021;29:1446–54.

53. Welt CK, Pagan YL, Smith PC, Rado KB, Hall JE. Control of follicle-stimulating hormone by estradiol and the inhibins: critical role of estradiol at the hypothalamus during the luteal-follicular transition. J Clin Endocrinol Metab 2003;88:1766–71.

54. Yding Andersen C. Inhibin-B secretion and FSH isoform distribution may play an integral part of follicular selection in the natural menstrual cycle. Molecular Hum Reprod 2017;23:16–24.

55. Dewailly D, Barbotin AL, Dumont A, Catteau-Jonard S, Robin G. Role of Anti-Mullerian Hormone in the Pathogenesis of Polycystic Ovary Syndrome. Front Endocrinol (Lausanne) 2020;11:641.

56. Laurent MR, Hammond GL, Blokland M, Jardi F, Antonio L, Dubois V, Khalil R, Sterk SS, Gielen E, Decallonne B, Carmeliet G, Kaufman JM, Fiers T, Huhtaniemi IT, Vanderschueren D, Claessens F. Sex hormone-binding globulin regulation of androgen bioactivity in vivo: validation of the free hormone hypothesis. Sci Rep 2016;6:35539.

57. Drummond AE. The role of steroids in follicular growth. Reprod Biol Endocrinol: RB&E 2006;4:16.

58. Bristol-Gould SK, Kreeger PK, Selkirk CG, Kilen SM, Cook RW, Kipp JL, Shea LD, Mayo KE, Woodruff TK. Postnatal regulation of germ cells by activin: the establishment of the initial follicle pool. Dev Biol 2006;298:132–48.

59. Ruth KS, Beaumont RN, Tyrrell J, Jones SE, Tuke MA, Yaghootkar H, Wood AR, Freathy RM, Weedon MN, Frayling TM, Murray A. Genetic evidence that lower circulating FSH levels lengthen menstrual cycle, increase age at menopause and impact female reproductive health. Hum Reprod 2016;31:473–81.

60. Hudecova M, Holte J, Olovsson M, Sundstrom Poromaa I. Long-term follow-up of patients with polycystic ovary syndrome: reproductive outcome and ovarian reserve. Hum Reprod 2009;24:1176–83.

61. Koivunen R, Pouta A, Franks S, Martikainen H, Sovio U, Hartikainen AL, McCarthy MI, Ruokonen A, Bloigu A, Jarvelin MR, Morin-Papunen L, Northern Finland Birth Cohort S. Fecundability and spontaneous abortions in women with self-reported oligo-amenorrhea and/or hirsutism: Northern Finland Birth Cohort 1966 Study. Hum Reprod 2008;23:2134–9.

62. West S, Vahasarja M, Bloigu A, Pouta A, Franks S, Hartikainen AL, Jarvelin MR, Corbett S, Vaarasmaki M, Morin-Papunen L. The impact of self-reported oligo-amenorrhea and hirsutism on fertility and lifetime reproductive success: results from the Northern Finland Birth Cohort 1966. Hum Reprod 2014;29:628–33.

63. Persson S, Elenis E, Turkmen S, Kramer MS, Yong EL, Sundstrom-Poromaa I. Fecundity among women with polycystic ovary syndrome (PCOS)-a population-based study. Hum Reprod 2019;34:2052–60.

64. Elting MW, Korsen TJ, Rekers-Mombarg LT, Schoemaker J. Women with polycystic ovary syndrome gain regular menstrual cycles when ageing. Hum Reprod 2000;15:24–8.

65. Carmina E, Campagna AM, Lobo RA. A 20-year follow-up of young women with polycystic ovary syndrome. Obstet Gynecol 2012;119:263–9.

66. Pigny P, Merlen E, Robert Y, Cortet-Rudelli C, Decanter C, Jonard S, Dewailly D. Elevated serum level of anti-mullerian hormone in patients with polycystic ovary syndrome: relationship to the ovarian follicle excess and to the follicular arrest. J Clin Endocrinol Metab 2003;88:5957–62.

67. Webber LJ, Stubbs S, Stark J, Trew GH, Margara R, Hardy K, Franks S. Formation and early development of follicles in the polycystic ovary. Lancet 2003;362:1017–21.

68. Emerging Risk Factors C, Erqou S, Kaptoge S, Perry PL, Di Angelantonio E, Thompson A, White IR, Marcovina SM, Collins R, Thompson SG, Danesh J. Lipoprotein(a) concentration and the risk of coronary heart disease, stroke, and nonvascular mortality. JAMA 2009;302:412–23.

69. Bots SH, Peters SAE, Woodward M. Sex differences in coronary heart disease and stroke mortality: a global assessment of the effect of ageing between 1980 and 2010. BMJ Glob Health 2017;2:e000298.

70. Vitale C, Fini M, Speziale G, Chierchia S. Gender differences in the cardiovascular effects of sex hormones. Fundam Clin Pharmacol 2010;24:675–85.

71. Schmidt J, Brannstrom M, Landin-Wilhelmsen K, Dahlgren E. Reproductive hormone levels and anthropometry in postmenopausal women with polycystic ovary syndrome (PCOS): a 21-year follow-up study of women diagnosed with PCOS around 50 years ago and their age-matched controls. J Clin Endocrinol Metab 2011;96:2178–85.

72. Puurunen J, Piltonen T, Morin-Papunen L, Perheentupa A, Jarvela I, Ruokonen A, Tapanainen JS. Unfavorable hormonal, metabolic, and inflammatory alterations persist after menopause in women with PCOS. J Clin Endocrinol Metab 2011;96:1827–34.

73. Winkler TW, Day FR, Croteau-Chonka DC, Wood AR, Locke AE, Magi R, Ferreira T, Fall T, Graff M, Justice AE, Luan J, Gustafsson S, Randall JC, Vedantam S, Workalemahu T, Kilpelainen TO, Scherag A, Esko T, Kutalik Z, Heid IM, Loos RJ, Genetic Investigation of Anthropometric Traits C. Quality control and conduct of genome-wide association meta-analyses. Nat Protoc 2014;9:1192–212.

74. Magi R, Morris AP. GWAMA: software for genome-wide association meta-analysis. BMC Bioinformatics 2010;11:288.

75. Willer CJ, Li Y, Abecasis GR. METAL: fast and efficient meta-analysis of genomewide association scans. Bioinformatics 2010;26:2190–1.

76. Watanabe K, Taskesen E, van Bochoven A, Posthuma D. Functional mapping and annotation of genetic associations with FUMA. Nat Commun 2017;8:1826.

77. Benner C, Spencer CC, Havulinna AS, Salomaa V, Ripatti S, Pirinen M. FINEMAP: efficient variable selection using summary data from genome-wide association studies. Bioinformatics 2016;32:1493–501..

78. Dewey FE, Murray MF, Overton JD, Habegger L, Leader JB, Fetterolf SN, O’Dushlaine C, Van Hout CV, Staples J, Gonzaga-Jauregui C, Metpally R, Pendergrass SA, Giovanni MA, Kirchner HL, Balasubramanian S, Abul-Husn NS, Hartzel DN, Lavage DR, Kost KA, Packer JS, Lopez AE, Penn J, Mukherjee S, Gosalia N, Kanagaraj M, Li AH, Mitnaul LJ, Adams LJ, Person TN, Praveen K, Marcketta A, Lebo MS, Austin-Tse CA, Mason-Suares HM, Bruse S, Mellis S, Phillips R, Stahl N, Murphy A, Economides A, Skelding KA, Still CD, Elmore JR, Borecki IB, Yancopoulos GD, Davis FD, Faucett WA, Gottesman O, Ritchie MD, Shuldiner AR, Reid JG, Ledbetter DH, Baras A, Carey DJ. Distribution and clinical impact of functional variants in 50,726 whole-exome sequences from the DiscovEHR study. Science 2016;354.

79. Dennis JK, Sealock JM, Straub P, Lee YH, Hucks D, Actkins K, Faucon A, Feng YA, Ge T, Goleva SB, Niarchou M, Singh K, Morley T, Smoller JW, Ruderfer DM, Mosley JD, Chen G, Davis LK. Clinical laboratory test-wide association scan of polygenic scores identifies biomarkers of complex disease. Genome Med 2021;13:6.

80. Staley JR, Blackshaw J, Kamat MA, Ellis S, Surendran P, Sun BB, Paul DS, Freitag D, Burgess S, Danesh J, Young R, Butterworth AS. PhenoScanner: a database of human genotype-phenotype associations. Bioinformatics 2016;32:3207–9.

81. Kamat MA, Blackshaw JA, Young R, Surendran P, Burgess S, Danesh J, Butterworth AS, Staley JR. PhenoScanner V2: an expanded tool for searching human genotype-phenotype associations. Bioinformatics 2019;35:4851–3.

82. Sun BB, Chiou J, Traylor M, Benner C, Hsu YH, Richardson TG, Surendran P, Mahajan A, Robins C, Vasquez-Grinnell SG, Hou L, Kvikstad EM, Burren OS, Davitte J, Ferber KL, Gillies CE, Hedman AK, Hu S, Lin T, Mikkilineni R, Pendergrass RK, Pickering C, Prins B, Baird D, Chen CY, Ward LD, Deaton AM, Welsh S, Willis CM, Lehner N, Arnold M, Worheide MA, Suhre K, Kastenmuller G, Sethi A, Cule M, Raj A, Alnylam Human G, AstraZeneca Genomics I, Biogen Biobank T, Bristol Myers S, Genentech Human G, GlaxoSmithKline Genomic S, Pfizer Integrative B, Population Analytics of Janssen Data S, Regeneron Genetics C, Burkitt-Gray L, Melamud E, Black MH, Fauman EB, Howson JMM, Kang HM, McCarthy MI, Nioi P, Petrovski S, Scott RA, Smith EN, Szalma S, Waterworth DM, Mitnaul LJ, Szustakowski JD, Gibson BW, Miller MR, Whelan CD. Plasma proteomic associations with genetics and health in the UK Biobank. Nature 2023;622:329–38.

83. Sun BB, Kurki MI, Foley CN, Mechakra A, Chen CY, Marshall E, Wilk JB, Biogen Biobank T, Chahine M, Chevalier P, Christe G, FinnGen, Palotie A, Daly MJ, Runz H. Genetic associations of protein-coding variants in human disease. Nature 2022;603:95–102.

84. Weeks EM, Ulirsch JC, Cheng NY, Trippe BL, Fine RS, Miao J, Patwardhan TA, Kanai M, Nasser J, Fulco CP, Tashman KC, Aguet F, Li T, Ordovas-Montanes J, Smillie CS, Biton M, Shalek AK, Ananthakrishnan AN, Xavier RJ, Regev A, Gupta RM, Lage K, Ardlie KG, Hirschhorn JN, Lander ES, Engreitz JM, Finucane HK. Leveraging polygenic enrichments of gene features to predict genes underlying complex traits and diseases. Nat Genet 2023;55:1267–76.

85. Sudlow C, Gallacher J, Allen N, Beral V, Burton P, Danesh J, Downey P, Elliott P, Green J, Landray M, Liu B, Matthews P, Ong G, Pell J, Silman A, Young A, Sprosen T, Peakman T, Collins R. UK biobank: an open access resource for identifying the causes of a wide range of complex diseases of middle and old age. PLoS Med 2015;12:e1001779.

86. Choi SW, Mak TS, O’Reilly PF. Tutorial: a guide to performing polygenic risk score analyses. Nat Protoc 2020;15:2759–72.

87. Prive F, Arbel J, Vilhjalmsson BJ. LDpred2: better, faster, stronger. Bioinformatics 2020;36:5424–31.

88. Bliddal M, Broe A, Pottegard A, Olsen J, Langhoff-Roos J. The Danish Medical Birth Register. Eur J Epidemiol 2018;33:27–36.

89. Schmidt M, Schmidt SA, Sandegaard JL, Ehrenstein V, Pedersen L, Sorensen HT. The Danish National Patient Registry: a review of content, data quality, and research potential. Clin Epidemiol 2015;7:449–90.

90. Burgess S, Butterworth A, Thompson SG. Mendelian randomization analysis with multiple genetic variants using summarized data. Genet Epidemiol 2013;37:658–65.

91. Burgess S, Thompson SG. Interpreting findings from Mendelian randomization using the MR-Egger method. Eur J Epidemiol 2017;32:377–89.

92. Burgess S, Thompson SG. Multivariable Mendelian randomization: the use of pleiotropic genetic variants to estimate causal effects. Am J Epidemiol 2015;181:251–60.

93. Yengo L, Sidorenko J, Kemper KE, Zheng Z, Wood AR, Weedon MN, Frayling TM, Hirschhorn J, Yang J, Visscher PM, Consortium G. Meta-analysis of genome-wide association studies for height and body mass index in approximately 700000 individuals of European ancestry. Hum Molec Genet 2018;27:3641–9.

