## Supplementary Information for "Genomic and proteomic evidence for hormonal and metabolic foundations of polycystic ovary syndrome"

### Subjects

The first stage of the meta-analysis included 10,563 cases and 445,078 controls from twenty cohorts of European, African American, Hispanic American and South Asian descent. Cases were diagnosed with PCOS based on NIH or Rotterdam Criteria, by self-report or using International Classification of Disease codes for PCOS or irregular menses and hirsutism (**Supplementary Table 1**). The NIH criteria require the presence of both OD and clinical and/or biochemical HA for a diagnosis of PCOS^1^. The Rotterdam criteria require two out of three features 1) OD defined by oligo- or amenorrhea (chronic menstrual cycle interval >35 days in all cohorts), 2) clinical and/or biochemical hyperandrogenism (HA) and/or 3) PCOM for a diagnosis of PCOS^1^. Non-NIH Rotterdam was defined by OD and PCOM or clinical and/or biochemical hyperandrogenism (HA) and PCOM. This was combined with previous published data. Self-reported female cases from research participants in the 23andMe, Inc. (Mountain View, CA, USA) cohort either responded “yes” to the question “Have you ever been diagnosed with polycystic ovary syndrome?” or indicated a diagnosis of PCOS when asked about fertility (“Have you ever been diagnosed with PCOS?” or “What was your diagnosis? Please check all that apply.” Answer=PCOS), hair loss in men or women (“Have you been diagnosed with any of the following? Please check all that apply.” Answer=PCOS) or research question (“Have you ever been diagnosed with PCOS?”)^5^. 23andMe controls were female, only.

HA was defined as hirsutism and quantified by the Ferriman-Gallwey (FG) score. The FG score assesses terminal hair growth in a male pattern in females, and a score above the upper limit of normal controls (>8) is considered hirsutism^3^. Hyperandrogenemia was defined as testosterone, androstenedione or DHEAS greater than the 95% confidence limits in control subjects in the individual population. OD was defined as cycle interval <21 or >35 days. PCOM was defined as 12 or more follicles of 2-9 mm in at least one ovary or an ovarian volume >10 mL^4^. The quantitative PCOS traits included levels of total testosterone (T), follicle-stimulating hormone (FSH), and luteinizing hormone (LH) and ovarian volume. An overview of the cohorts, diagnostic criteria and number of subjects included in each subphenotype or trait analysis are summarised in Supplementary Table 27.

**Supplementary Table 27. International Classification of Disease (ICD) codes used to identify women with PCOS in electronic medical records.**

| **Diagnosis** | **ICD8** | **ICD9** | **ICD10** |
| --- | --- | --- | --- |
| **INCLUSION DIAGNOSES** | | | |
| PCOS | 256.9 | 256.4 | E28.2 |
| Irregular Menses | 626 | 626.X | N92 |
| Hirsutism | 704 | 704.1 | L68.0 |
| **EXCLUSION DIAGNOSES** | | | |
| Premature Ovarian Failure | 256.1, 256.3 | 256.3, 256.31 | E28.39 |
| Cushing Syndrome | 255, 258 | 255.0, 258.0 | E24 |
| Hypothalamic amenorrhea | 256.1 | 256.8 | E23.3 |
| Congenital Adrenal Hyperplasia | 255 | 255.2 | E25 |
| Eating disorder | 306.5 | 307.1x, 307.5x | F50.9, F50.2, F50.01 |
| Chronic opioid use | 304 | 304.X | F11 |
| Fibroids | 218 | 654.1X, 218.9 | O34.1, D25.9 |
| Pituitary adenoma | 226.2 | 253.X | D352 |
| Pituitary hypersecretion | 253 | 253.X | E22 |
| Hyperprolactinemia | 253 | 253.1 | E22 |
| Ovarian tumor | 183 | 239.5 | C56 |
| Benign neoplasm of the ovary (Leydig cell tumor, hilus cell tumor) | 220 | 220 | D27.9 |
| Turner syndrome | 759.5 | 758.6 | Q96 |
| Galactorrhea | 253 | 253.1 | N64.3 |
| Suprarenal tumor | 226.0 | 227.0, 194.0 | C74 |

### Additional Cohort-specific Details

##### FinnGen

We want to acknowledge the participants and investigators of FinnGen study. The FinnGen project is funded by two grants from Business Finland (HUS 4685/31/2016 and UH 4386/31/2016) and the following industry partners: AbbVie Inc., AstraZeneca UK Ltd, Biogen MA Inc., Bristol Myers Squibb (and Celgene Corporation & Celgene International II Sàrl), Genentech Inc., Merck Sharp & Dohme LCC, Pfizer Inc., GlaxoSmithKline Intellectual Property Development Ltd., Sanofi US Services Inc., Maze Therapeutics Inc., Janssen Biotech Inc, Novartis Pharma AG, and Boehringer Ingelheim International GmbH. Following biobanks are acknowledged for delivering biobank samples to FinnGen: Auria Biobank (www.auria.fi/biopankki), THL Biobank (www.thl.fi/biobank), Helsinki Biobank (www.helsinginbiopankki.fi), Biobank Borealis of Northern Finland (<https://www.ppshp.fi/Tutkimus-ja-opetus/Biopankki/Pages/Biobank-Borealis-briefly-in-English.aspx>), Finnish Clinical Biobank Tampere (www.tays.fi/en-US/Research_and_development/Finnish_Clinical_Biobank_Tampere), Biobank of Eastern Finland (www.ita-suomenbiopankki.fi/en), Central Finland Biobank (www.ksshp.fi/fi-FI/Potilaalle/Biopankki), Finnish Red Cross Blood Service Biobank ([www.veripalvelu.fi/verenluovutus/biopankkitoiminta](http://www.veripalvelu.fi/verenluovutus/biopankkitoiminta)), Terveystalo Biobank ([www.terveystalo.com/fi/Yritystietoa/Terveystalo-Biopankki/Biopankki/](http://www.terveystalo.com/fi/Yritystietoa/Terveystalo-Biopankki/Biopankki/)) and Arctic Biobank (<https://www.oulu.fi/en/university/faculties-and-units/faculty-medicine/northern-finland-birth-cohorts-and-arctic-biobank>). All Finnish Biobanks are members of BBMRI.fi infrastructure ([www.bbmri.fi](http://www.bbmri.fi/)). Finnish Biobank Cooperative -FINBB (<https://finbb.fi/>) is the coordinator of BBMRI-ERIC operations in Finland. The Finnish biobank data can be accessed through the Fingenious^®^ services (<https://site.fingenious.fi/en/>) managed by FINBB.

##### Estonian BioBank

The Estonian Biobank (EstBB) is a population-based biobank with over 200,000 participants, currently including approximately 135,000 women (20% of the female Estonian population). The 150K data freeze was used for the analyses described in this paper. All biobank participants have signed a broad informed consent form. Individuals with PCOS were identified using the ICD-10 code E28.2, and all female biobank participants who did not have this diagnosis served as controls. This included a total of 2,812 cases and 89,230 controls. Information on the ICD codes was obtained via regular linking with the national Health Insurance Fund and other relevant databases.

**Estonian Biobank research team** (Affiliation: Estonian Genome Centre, Institute of Genomics, University of Tartu /): Andres Metspalu, Lili Milani, Tõnu Esko, Reedik Mägi, Mari Nelis and Georgi Hudjashov.

Leitsalu L, Haller T, Esko T, Tammesoo ML, Alavere H, Snieder H, et al. Cohort Profile: Estonian Biobank of the Estonian Genome Center, University of Tartu. Int J Epidemiol 2015 Aug;44(4):1137- 1147.

##### Genes and Health

Genes & Health is/has recently been core-funded by Wellcome (WT102627, WT210561), the Medical Research Council (UK) (M009017, MR/X009777/1), Higher Education Funding Council for England Catalyst, Barts Charity (845/1796), Health Data Research UK (for London substantive site), and research delivery support from the NHS National Institute for Health Research Clinical Research Network (North Thames). Genes & Health is/has recently been funded by Alnylam Pharmaceuticals, Genomics PLC; and a Life Sciences Industry Consortium of Astra Zeneca PLC, Bristol-Myers Squibb Company, GlaxoSmithKline Research and Development Limited, Maze Therapeutics Inc, Merck Sharp & Dohme LLC, Novo Nordisk A/S, Pfizer Inc, Takeda Development Centre Americas Inc.

We thank Social Action for Health, Centre of The Cell, members of our Community Advisory Group, and staff who have recruited and collected data from volunteers. We thank the NIHR National Biosample Centre (UK Biocentre), the Social Genetic & Developmental Psychiatry Centre (King's College London), Wellcome Sanger Institute, and Broad Institute for sample processing, genotyping, sequencing and variant annotation. We thank: Barts Health NHS Trust, NHS Clinical Commissioning Groups (City and Hackney, Waltham Forest, Tower Hamlets, Newham, Redbridge, Havering, Barking and Dagenham), East London NHS Foundation Trust, Bradford Teaching Hospitals NHS Foundation Trust, Public Health England (especially David Wyllie), Discovery Data Service/Endeavour Health Charitable Trust (especially David Stables), NHS Digital - for GDPR-compliant data sharing backed by individual written informed consent. Most of all we thank all of the volunteers participating in Genes & Health.

##### Danish Blood Donors Study (DBDS) Genomic Consortium

Karina Banasik^1^, Jakob Bay^2^, Jens Kjaergaard Boldsen^5^, Thosten Brodersen^2^, Soren Brunak ^1^, Kristoffer Burgdorf ^1^, Mona Ameri Chalmer^3^, Maria Didriksen^7^, Khoa Manh Dinh ^5^, Joseph Dowsett^7^, Bjarke Feenstra ^7,8^, Frank Geller ^7,8^, Daniel Gudbjartsson ^4^, Thomas Folkmann Hansen^3^, Lotte Hindhede ^5^, Henrik Hjalgrim^14,15^, Rikke Louise Jacobsen ^7^, Gregor Jemec^10^, Bertram Dalskov Kjerulff^5^, Lisette Kogelman^3^, Margit Anita Horup Larsen ^7^, Ioannis Louloudis^1^, Agnete Lundgaard ^1^, Susan Mikkelsen^5^, Ioanna Nissen ^7^, Alexander Pil Henriksen^1^, Palle Duun Rohde^11^, Klaus Rostgaard^14,15^, Michael Schwinn ^7^, Kari Stefansson^4^, Hreinn Stefansson ^4^, Erik Sorensen ^7^, Unnur Thorsteinsdottir ^4^, Lise Wegner Thorner^7^, Thomas Werge^12,13^, Mette Nyegaard^11^, Sisse R. Ostrowski ^7^, Ole B.V. Pedersen^2^, Christina Mikkelsen ^7^, Christian Erikstrup ^5,6^, Katrine Kaspersen^5^, Mie T. Bruun^9^, Bitten Aagaard^5^, Henrik Ullum^16^, David Westergaard^1^

1 Novo Nordisk Foundation Center for Protein Research, Faculty of Health and Medical Sciences, University of Copenhagen, Copenhagen, Denmark

2 Department of Clinical Immunology, Zealand University Hospital, Køge, Denmark

3 Danish Headache Center, Department of Neurology, Copenhagen University Hospital, Rigshospitalet-Glostrup, Copenhagen, Denmark

4 deCODE Genetics, Reykjavik, Iceland

5 Department of Clinical Immunology, Aalborg University Hospital, Aalborg, Denmark

6 Department of Clinical Medicine, Health, Aarhus University, Aarhus, Denmark

7 Department of Clinical Immunology, Copenhagen University Hospital, Rigshospitalet, Copenhagen, Denmark

8 Department of Epidemiology Research, Statens Serum Institut, Copenhagen, Denmark

9 Department of Clinical Immunology, Odense University Hospital, Odense, Denmark

10 Department of Dermatology, Zealand University hospital, Roskilde, Denmark

11 Department of Health Science and Technology, Faculty of Medicine, Aalborg University, Aalborg, Denmark

12 Institute of Biological Psychiatry, Mental Health Centre, Sct. Hans, Copenhagen University Hospital, Roskilde, Denmark

13 Department of Clinical Medicine, Faculty of Health and Medical Sciences, University of Copenhagen, Copenhagen, Denmark

14 Danish Cancer Society Research Center, Copenhagen, Denmark

15 Department of Epidemiology Research, Statens Serum Institut, Copenhagen, Denmark

16 Statens Serum Institut, Copenhagen, Denmark

#### Other Funding and Acknowledgements

Funding: The WGHS is supported by the NHLBI (HL043851 and HL080467) and the NCI (CA047988 and UM1CA182913). This work has been supported by F32 HD103317, K08 HD110723 from the National Institute of Child Health and Human Development, 23CDA1054471 from the American Heart Association, Pediatric Endocrine Society Clinical Scholar Award and Boston Children’s Hospital Office of Faculty Development Career Development Fellowship (JZ), MATER Marie Sklodowska-Curie which received funding from the European Union's Horizon 2020 research and innovation program under grant agreement No. 813707 (NPG), MRC grant MC_U106179472 YZ, KAK, FRD, KKO, JRBP), Samuel Oschin Comprehensive Cancer Institute Developmental Funds, Center for Bioinformatics and Functional Genomics and Department of Biomedical Sciences Developmental Funds (MRJ), Novo Nordisk Foundation (grants NNF17OC0027594 and NNF14CC0001 (KB and SB), NCI P30CA177558 (CH), NIDDK R01DK075787 (JNH), NCI UM1CA186107 (PK), European Regional Development Fund (Project No. 2014-2020.4.01.15-0012) and the European Union’s Horizon 2020 research and innovation program under grant agreements No 692065 (TL, RM, AS) and 692145 (RM), Estonian Research Council grant PRG1076 (AS), Horizon 2020 innovation grant ERIN grant EU952516 (AS), Horizon Europe NESTOR grant 101120075 (AS), NICHD R01HD065029 (RS), Estonian Ministry of Education and Research (grant IUT34-16 to TL), *NICHD R01HD057450 (MU), NICHD R01HD100630( MU, RL, MGH, CW),* NICHD P50HD044405 (AD), NICHD R01HD057223 (AD), R01HD085227 (MGH, AD) and R01 HD100812 (AD) from the Eunice Kennedy Shriver National Institute of Child Health and Human Development, deCode Genetics (GT, UT, KS, US*), NHMRC Ideas Grant 2003629 and DoH Western Australia Merit Award 1186046 (BHM),* SCGOPHCG RAC 2015-16/034 (SGW, BGAS), 2016-17/018 (BGAS), NIHR BRC, Wellcome Trust, MRC (TDS), Eris M. Field Chair in Diabetes Research (MOG), NIDDK P30 DK063491 (MOG), NIDDK U01DK094431, U01DK048381 (DE), NICHD U10HD38992 (RL), Estonian Ministry of Education and Research (grant IUT34-16), Enterprise Estonia (grant EU48695); the EU-FP7 Marie Curie Industry-Academia Partnerships and Pathways (IAPP, grant SARM, EU324509 to AS), Wellcome (090532, 098381, 203141); European Commission (ENGAGE: HEALTH-F4-2007-201413 to MIM), MRC G0802782, MR/M012638/1 (SF), Li Ka Shing Foundation, WT-SSI/John Fell Funds, NIHR Biomedical Research Centre, Oxford, Widenlife and NICHD 5P50HD028138-27 (CML), NICHD R01HD065029, ADA 1-10-CT-57, Harvard Clinical and Translational Science Center, from the National Center for Research Resources 1UL1 RR025758 (CKW). Novo Nordisk Foundation Data Science Investigator grant NNF20OC0062294 (TK). The funders had no role in study design, data collection and analysis, decision to publish, or preparation of the manuscript.

##### Address Changes

##### Mark I. McCarthy, Genentech, 1 DNA Way, South San Francisco, CA 94080

##### Competing interests

Members of the 23andMe Research team are employees of and hold stock or stock options in 23andMe, Inc. SB reports ownerships in Intomics A/S, Hoba Therapeutics Aps, Novo Nordisk A/S, Lundbeck A/S, ALK abello A/S and managing board memberships in Proscion A/S and Intomics A/S. GT, UT, KS, and US are employees of deCODE genetics/Amgen Inc. MIM serves on advisory panels for Pfizer and NovoNordisk. MIM has received honoraria from Pfizer, NovoNordisk and EliLilly, and has received research funding from Pfizer, NovoNordisk, EliLilly, AstraZeneca, Sanofi Aventis, Boehringer Ingelheim, Merck, Roche, Janssen, Takeda, and Servier. MIM is now an employee of Genentech and a holder of Roche stock. JL has received consultancy fees from Danone, Metagenics inc., Titus Healthcare, Roche and Euroscreen. RA has received consultancy fees from Spruce Biosciences, Fortress Biotech, Rani Therapeutics, Core Access Surgical Technologies, and Arora Forge Advisors; and has equity in Martin Imaging. MOG has served on an advisory board for Nestle Health Science. BCJM Fauser has received fees and/or grant support during the last 4 years from the following organisations (in alphabetic order); Bain Capital, Controversies in Obstetrics & Gynecology (COGI), Dutch Heart Foundation (Nederlandse Hartstichting), Elsevier, European Society of Human Reproduction and Embryology (ESHRE), Ferring, International Federation of Fertility Societies (IFFS), London Womens Clinic, Myovant, Netherlands Organisation for Health Research and Development (ZonMW), Pantharei Bioscience, Partners Group, PregLem/Gideon Richter, Shieldler, Reproductive Biomedicine Online (RBMO), UpToDate. Co-founder and shareholder of Zoe Ltd (TDS).

##### Ethics statements

All research involving human participants has been approved by the authors’ Institutional Review Board (IRB) or an equivalent committee, and all clinical investigation was conducted according to the principles expressed in the Declaration of Helsinki. Written informed consent was obtained from all participants.

The Boston cohort was approved by the Partners IRB (# 2002P001924 and 2012P002417) and the University of Utah IRB (IRB_00076659). The deCODE cohort was approved by the National Bioethics Committee of Iceland (VSN 03–007), which was conducted in agreement with conditions issued by the Data Protection Authority of Iceland. Personal identities of the participants’ data and biological samples were encrypted by a third-party system (Identity Protection System), approved and monitored by the Data Protection Authority.

The UK cohort was approved by the Parkside Health Authority (Now—NHS Health Research Authority, NRES Committee—West London & GTAC, UK, London, UK) under EC2359 "The Molecular Genetics of Polycystic Ovaries."

The Rotterdam PCOS cohort, the COLA study, was approved by institutional review board (Medical Ethics Committee) of the PCOS genetics meta-analysis PLOS Genetics | https://doi.org/10.1371/journal.pgen.1007813 December 19, 2018 10 / 20 Erasmus Medical Center (04–263). Controls from the Rotterdam Study were approved by the Medical Ethics Committee of the Erasmus MC (registration number MEC 02.1015) and by the Dutch Ministry of Health, Welfare and Sport (Population Screening Act WBO, license number 1071272-159521-PG). The Rotterdam Study Personal Registration Data collection is filed with the Erasmus MC Data Protection Officer under registration number EMC1712001. The Rotterdam Study has been entered into the Netherlands National Trial Register (NTR; www. trialregister.nl) and into the WHO International Clinical Trials Registry Platform (ICTRP; www.who.int/ictrp/network/primary/en/) under shared catalogue number NTR6831.

The Chicago PCOS cohort was approved by the Northwestern IRB (#STU00008096). The control subjects from the NUgene study were approved by the Northwestern IRB (# STU00010003).

The Estonia cohort was approved by the Research Ethics Committee of the University of Tartu approved the study (198T-18).

The Twins UK study was approved by the St Thomas’ Hospital Research Ethics Committee (EC04/015).

The Nurses’ Health Study (NHS I and II) was approved by the Partners Human Research Committee (#1999-P-011114).

Patients and control subjects in FinnGen provided informed consent for biobank research, based on the Finnish Biobank Act. Alternatively, older research cohorts, collected prior the start of FinnGen (in August 2017), were collected based on study-specific consents and later transferred to the Finnish biobanks after approval by the National Supervisory Authority for Welfare and Health, Fimea. Recruitment procedures followed the biobank protocols approved by Fimea. The Coordinating Ethics Committee of the Hospital District of Helsinki and Uusimaa (HUS) approved the FinnGen study protocol (Nr HUS/990/2017). The FinnGen study was approved by Finnish Institute for Health and Welfare (permit numbers: THL/2031/6.02.00/2017, THL/1101/5.05.00/2017, THL/341/6.02.00/2018, THL/2222/6.02.00/2018, THL/283/6.02.00/2019, THL/1721/5.05.00/2019, THL/1524/5.05.00/2020, and THL/2364/14.02/2020); Digital and population data service agency (permit numbers: VRK43431/2017-3, VRK/6909/2018-3, VRK/4415/2019-3); the Social Insurance Institution (permit numbers: KELA 58/522/2017, KELA 131/522/2018, KELA 70/522/2019, KELA 98/522/2019, KELA 138/522/2019, KELA 2/522/2020, KELA 16/522/2020); and Statistics Finland (permit numbers: TK-53- 1041-17 and TK-53-90-20). The Biobank access decisions for FinnGen samples and data utilized in the FinnGen Data Freeze 6 include: THL Biobank BB2017_55, BB2017_111, BB2018_19, BB_2018_34, BB_2018_67, BB2018_71, BB2019_7, BB2019_8, BB2019_26, BB2020_1, Finnish Red Cross Blood Service Biobank 7.12.2017, Helsinki Biobank HUS/359/2017, Auria Biobank AB17-5154, Biobank Borealis of Northern Finland_2017_1013, Biobank of Eastern Finland 1186/2018, Finnish Clinical Biobank Tampere MH0004, Central Finland Biobank 1-2017, and Terveystalo Biobank STB 2018001.

Analyses in the EstBB were carried out under ethical approval 1.1-12/624 from the Estonian Committee on Bioethics and Human Research and data release N05 from the EstBB.

### Supplementary Methods

##### Summary-data-based Mendelian Randomisation (SMR) Analysis

In order to identify genes whose expression levels are associated with PCOS status, we performed an integrative analysis on the PCOS GWAS meta-analysis results and expression quantitative trait locus (eQTL) association data for PCOS-relevant GTEx tissues using the SMR software^5,6^. This package utilises Mendelian randomisation principles to assess for association between gene expression and a trait due to a shared genetic variant (pleiotropy). Dual association signals in the GWAS and eQTL datasets are identified by testing for association between gene expression and the trait of interest at the top eQTL variant for each gene. The software also performs a heterogeneity in dependent instruments (HEIDI) test, which compares the association signals for nearby co-inherited markers in the GWAS and eQTL datasets. If heterogeneity exists in the association profiles of the two datasets, as indicated by a significant HEIDI test result, the association signals present in each dataset are considered less likely to be driven by the same causal variant. Genes with at least 1 *cis*-eQTL association significant at P<5×10^-8^ were included in the SMR analysis, with linkage disequilibrium data from the 1000 Genomes Project phase 3 dataset used for the HEIDI test. The SMR analysis was performed using the all-ancestries age-adjusted (not including 23andMe) PCOS GWAS meta-analysis dataset and the GTEx V7 eQTL datasets^7^ for the following PCOS-relevant tissues: adipose subcutaneous, adipose visceral omentum, adrenal gland, ovary, pancreas, pituitary and testis. Correction for multiple testing was performed for each tissue analysed using the Benjamini-Hochberg procedure, with a conservative significance threshold of P<0.05 used for the HEIDI test (*P_HEIDI_*) as an indicator of heterogeneity.

After correction for multiple testing, 26 significant associations were identified, including five for adipose subcutaneous, six for adipose visceral omentum, one for adrenal gland, two for ovary, three for pancreas, three for pituitary and six for testis. These tissues represent a range of important organs in the aetiology of PCOS. Some genes showed particularly strong evidence for an association with PCOS. The *RPS26* gene, which encodes a ribosomal protein, demonstrated significant pleiotropic associations in all tissues analysed except for the adrenal gland. In each instance, expression of RPS26 was found to be positively associated with PCOS risk (β_SMR_=0.06). In some cases the SMR analyses revealed more complex biological processes, with gene *ARL14EP* prioritised on chromosome 11, despite strong *a-priori* evidence that this variant is acting through the nearby *FSHB*. Sometimes it provided better resolution of established signals, with the variant previously tagged as *GATA2* showing strong evidence of an effect via *NEIL2*. The *NEIL2* gene, encoding a DNA glycosylase enzyme, demonstrated significant pleiotropic associations in the adipose visceral omentum, ovary and pituitary tissues. In each of these tissues, expression of *NEIL2* was found to be negatively associated with PCOS risk (β_SMR_=-0.2 to -0.4). Finally, we were also able to show that increased expression of the established age-at-menopause gene *MSH6* ^6^ in the testis is positively associated with PCOS risk (β_SMR_=0.3).

##### Polygenic Risk Score (PRS) Phenotype Definitions in the UK Biobank

**1) Coronary artery disease (CAD)**^8^

Self-report: Age heart attack diagnosed

Ischemic Heart diseases

ICD-10 I21-I25.X

ICD-9 410-412.X

Coronary Revascularization:

OPCS4 K40,41,45,49,50,75

Date of MI: Combination of 5 outcome data fields

**2) Type 2 diabetes (T2D)**^9,10^

Baseline Visit Algorithm:

Diabetes diagnosis

Medication use

Age at diagnosis

Self-reported diagnosis

Repeat Visits

Diabetes diagnosis

Medication information

Exclusions

Age of diabetes <40 years

Controls >=55 years

**3) Breast Cancer**

Type of cancer:

ICD-9: 174X

ICD-10: C50.X

Cancer code, self-reported

1002

Hormonal cancer phenotypes: Breast cancer was based on self-report and/or ICD-9 codes 174.0-174.9 and ICD-10 codes C50.0-C50.9 in hospitalisation records.

**4) Childlessness**

Cases

Women: Field code 2734 (number of live births) = 0

Men: Field code 2405 (number of children fathered) = 0

Controls:

Women: Field code 2734 (number of live births) neither 0 or NA

Men: Field code 2405 (number of children fathered) neither 0 or NA

**5) Bipolar/depression status**^11^

Derived field 20126

Mental health phenotypes: Depression was defined by self-report of current and previous depressive symptoms and items from a Patient Health Questionnaire and on seeking evaluation for mental health, and controls were defined as individuals with no history of bipolar disorder or depression as previously reported^81^.

**6) Asthma**

Cases

1. Field code 6152_8 (doctor diagnosed asthma)

2. ICD-10: J45 (asthma)/J46 (severe asthma)

3. Self-reported asthma

Controls:

1. Free from field code 6152_8 (doctor diagnosed asthma)

2. Free from field code 6152_9 (doctor diagnosed allergic diseases)

3. Free from ICD10 J45/J46/J30 (hay fever)/L20 (dermatitis and eczema)

4. Free from self-reported asthma/hay fever/eczema/allergy/ allergy to house dust mite (HDM)

Asthma was defined by a composite of self-reported diagnosis of asthma, self-reported diagnosis of asthma by a doctor, and/or ICD-10 codes J45.0, J45.1, J45.8, J45.9, J46 from hospitalisation records. Controls were defined by individuals free from a diagnosis of hay fever (ICD-10 J30.0-J30.4), dermatitis and eczema (ICD-10 L20.8, L20.9), or self-reported diagnosis of asthma or other allergic disorders (hay fever, eczema, allergy, and allergy to house dust mite).

### Supplementary Figures

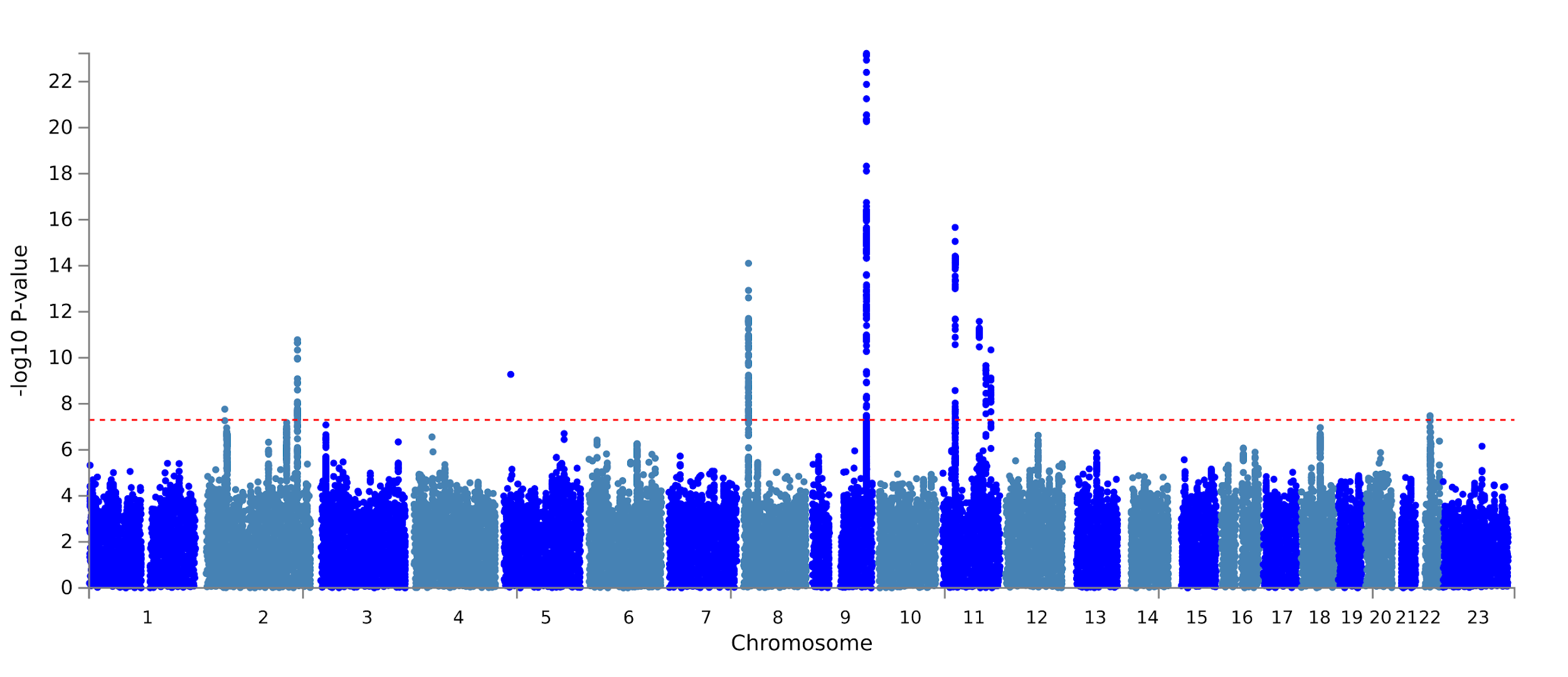

***Supplementary Figure 1*** *- Manhattan plot of the genome-wide data adjusting for BMI.*

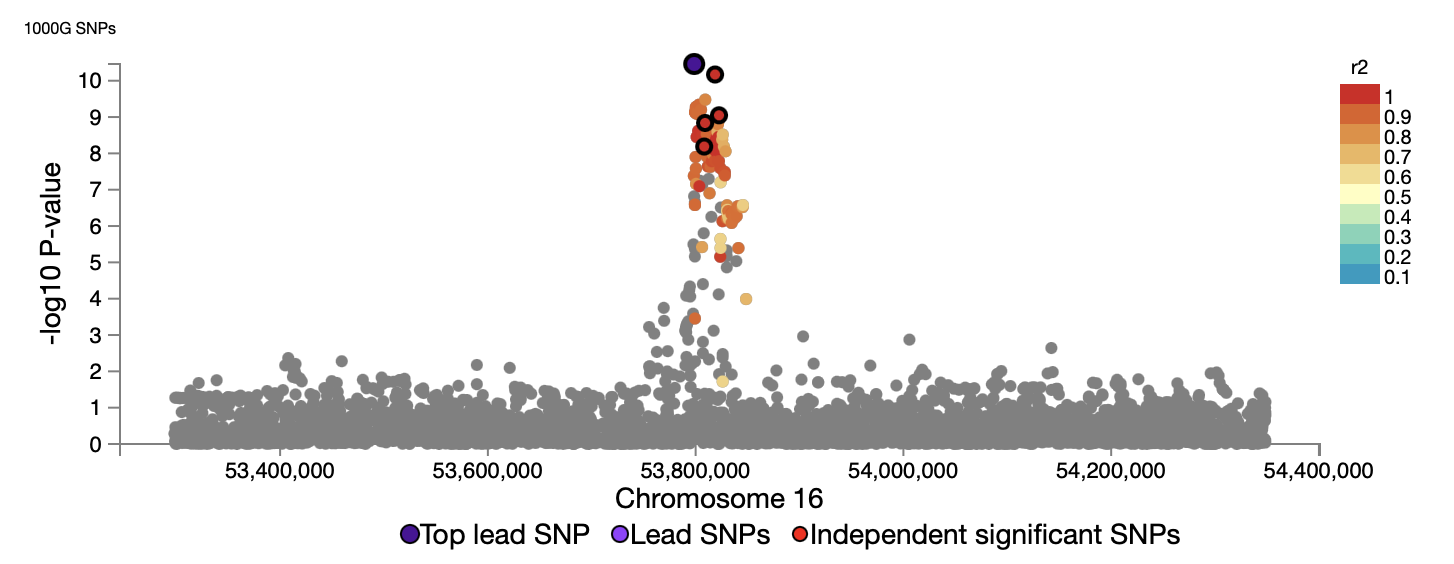

***Supplementary Figure 2*** *- Regional plot, generated by FUMA, for the FTO locus showing the top lead variant rs8047587 in purple in the age-adjusted meta-analysis (without 23andMe data).* A statistically significant association between the *FTO* variant and PCOS was not observed after BMI adjustment (p=0.02). Each variant is color-coded based on the highest r^2^ to one of the independent significant variants, if that is greater or equal to the user defined threshold (r^2^≥0.6). Other variants (i.e. below the user-defined r^2^) are colored in grey. The top lead variants in genomic risk loci, lead variants and independent significant variants are circled in black and colored in dark-purple, purple and red, respectively.

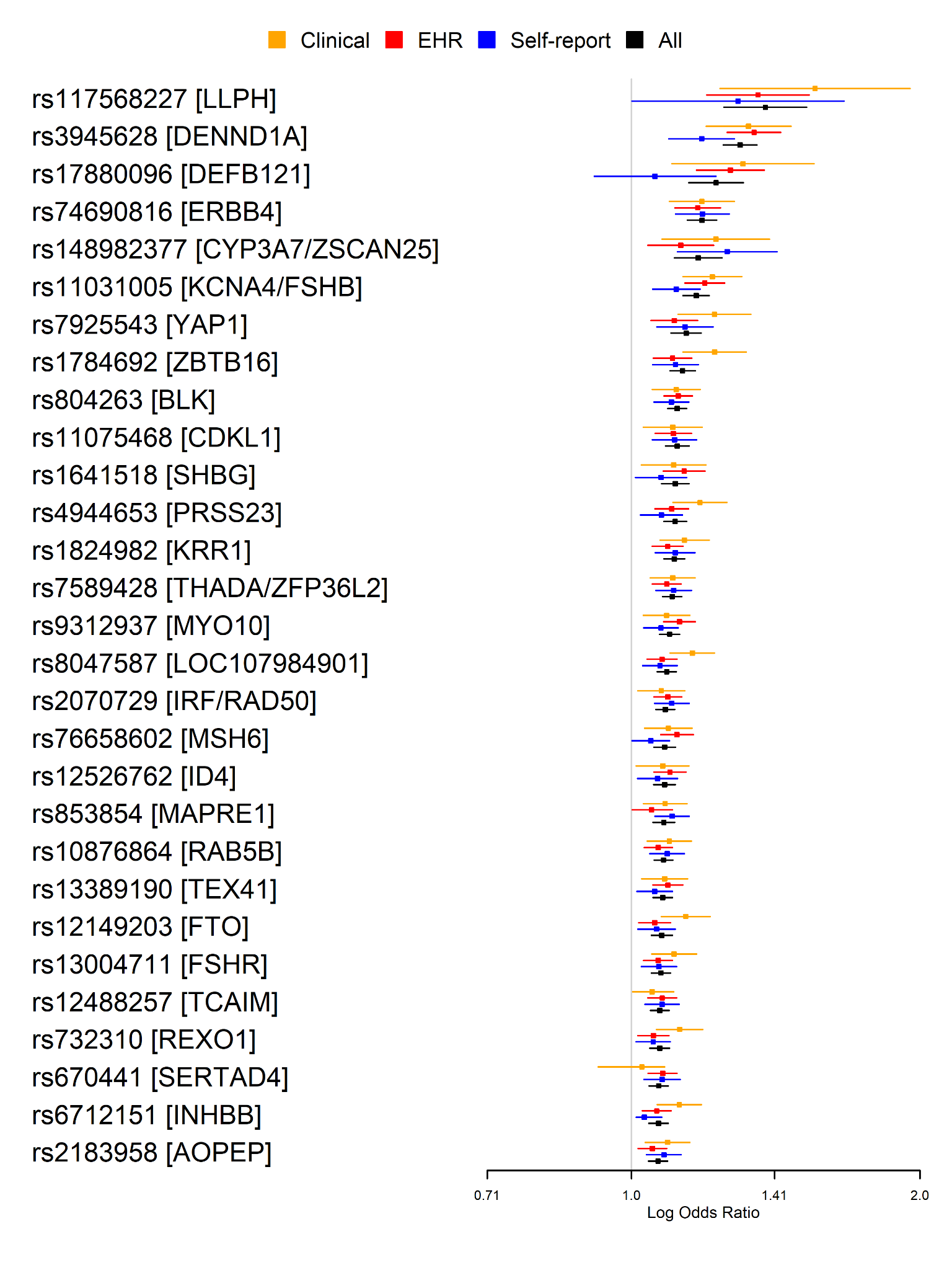

***Supplementary Figure 3*** *- Forest plot of the identified loci by source of PCOS designation.*

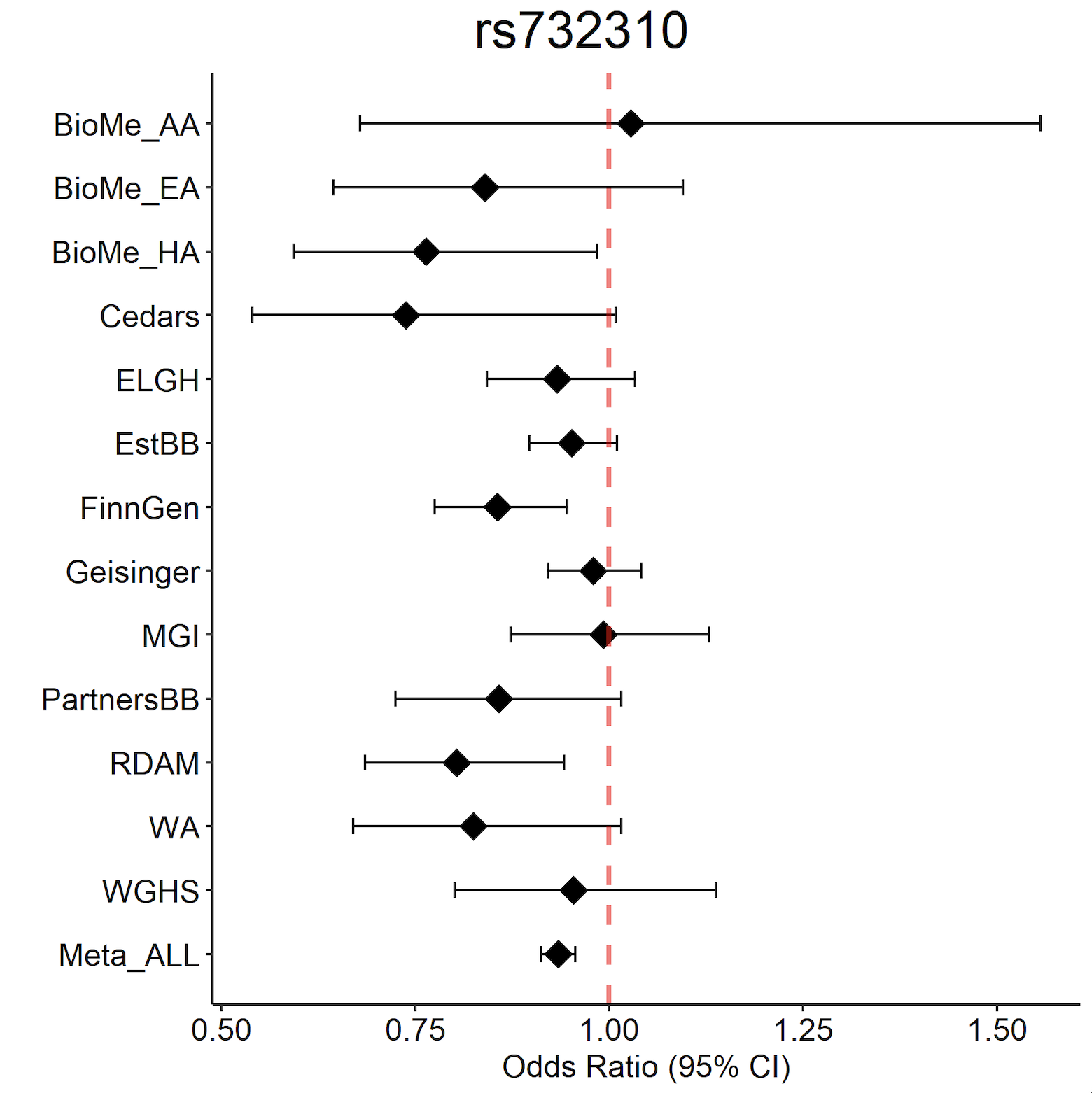

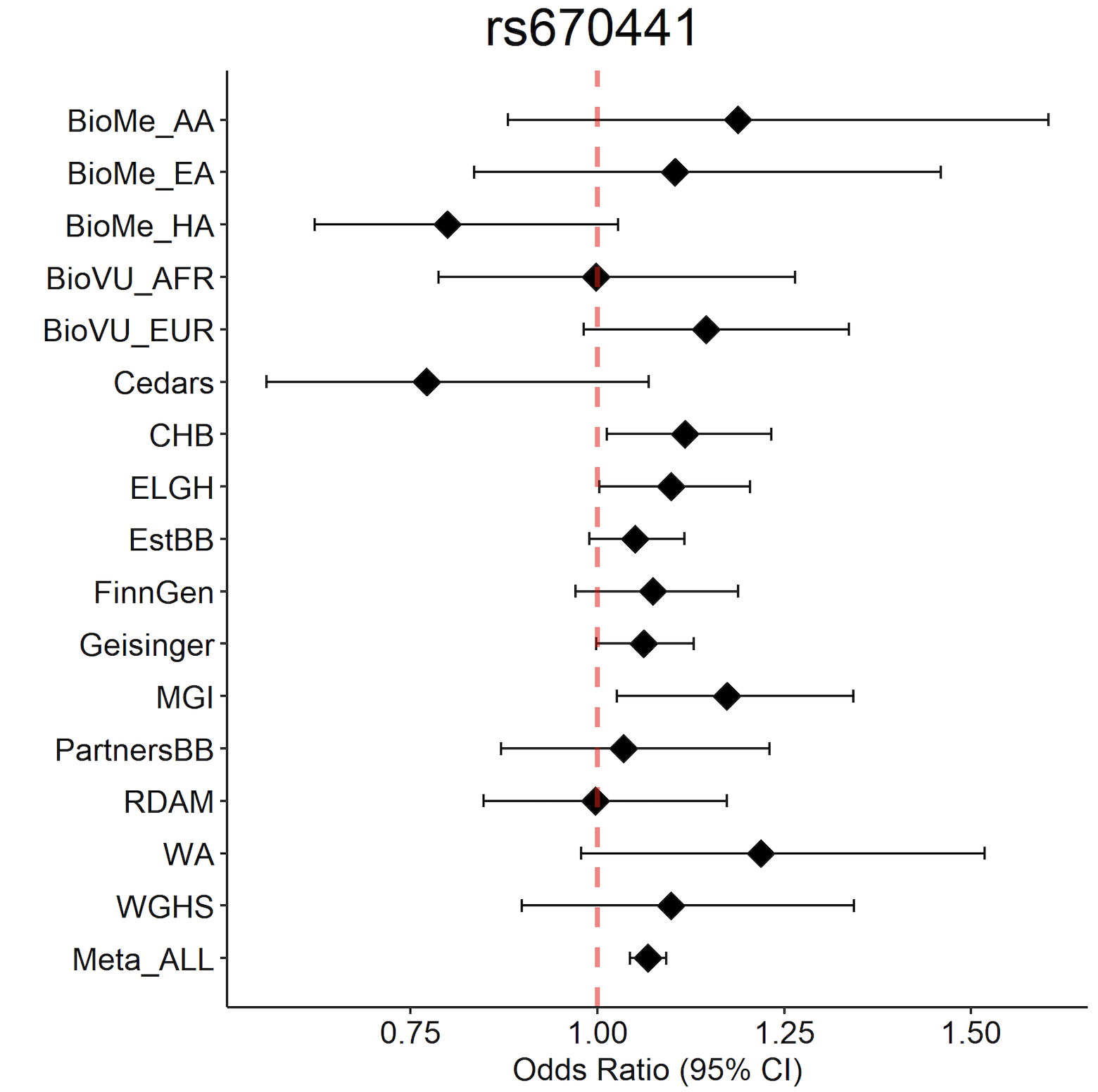

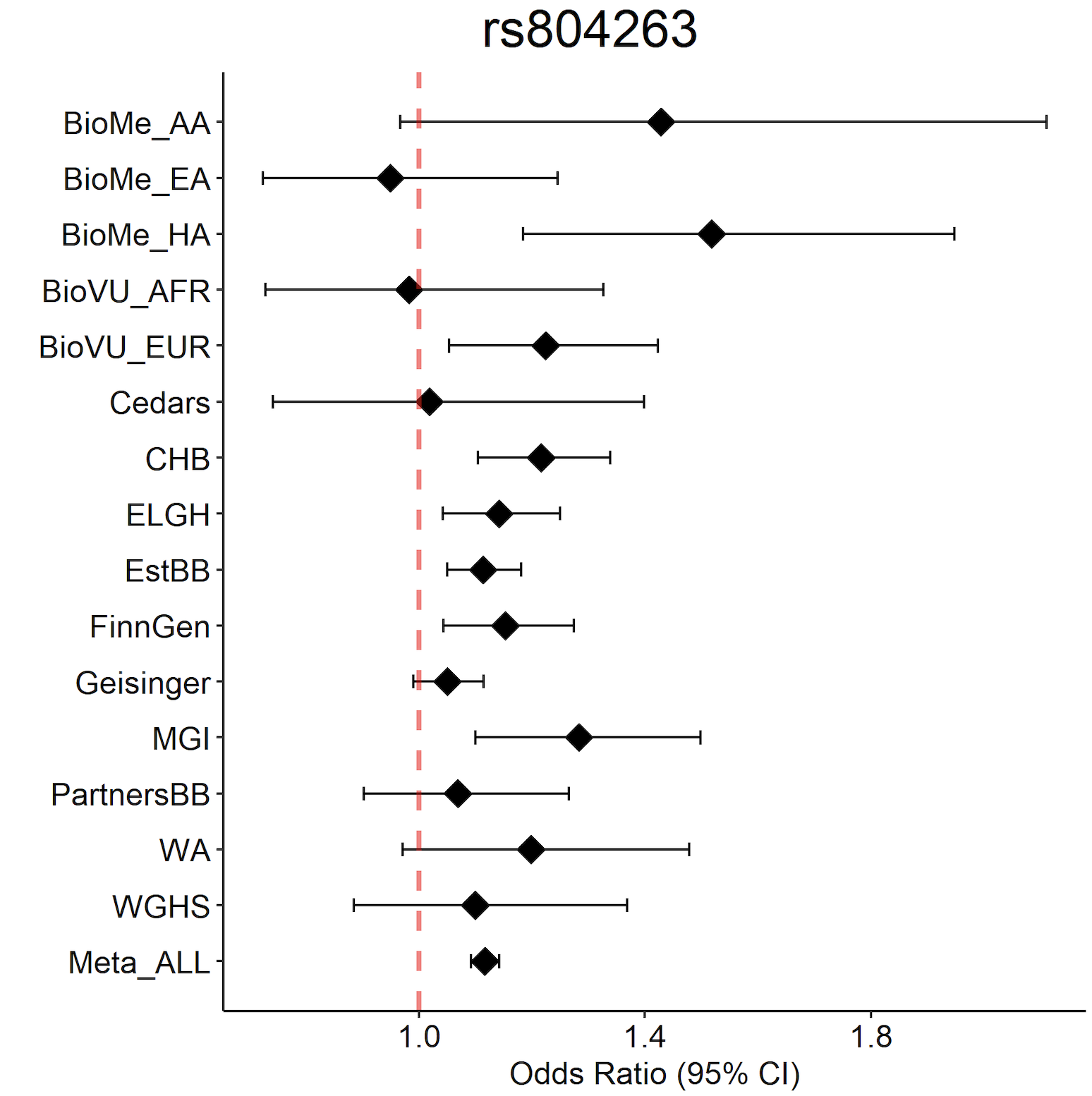

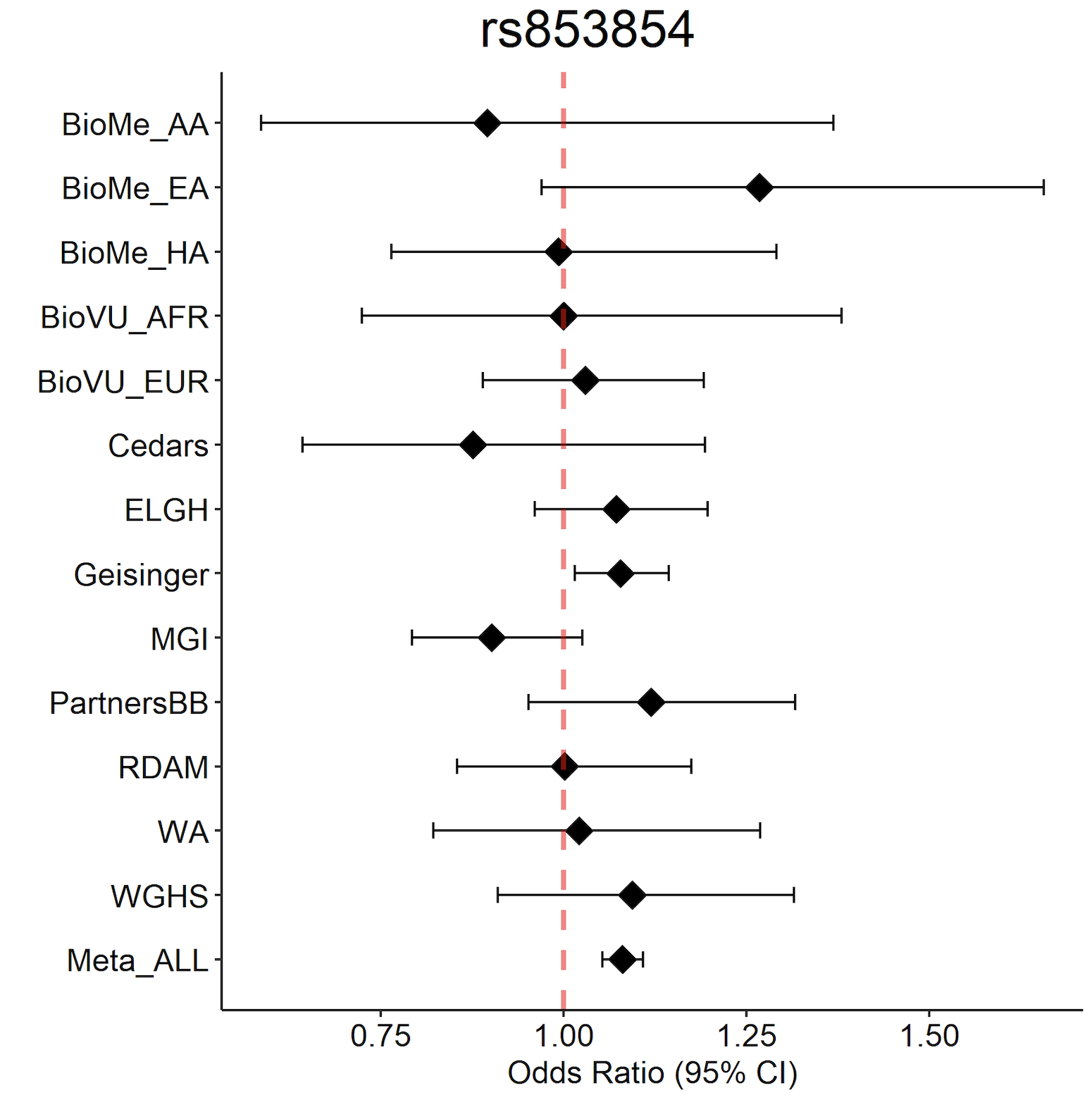

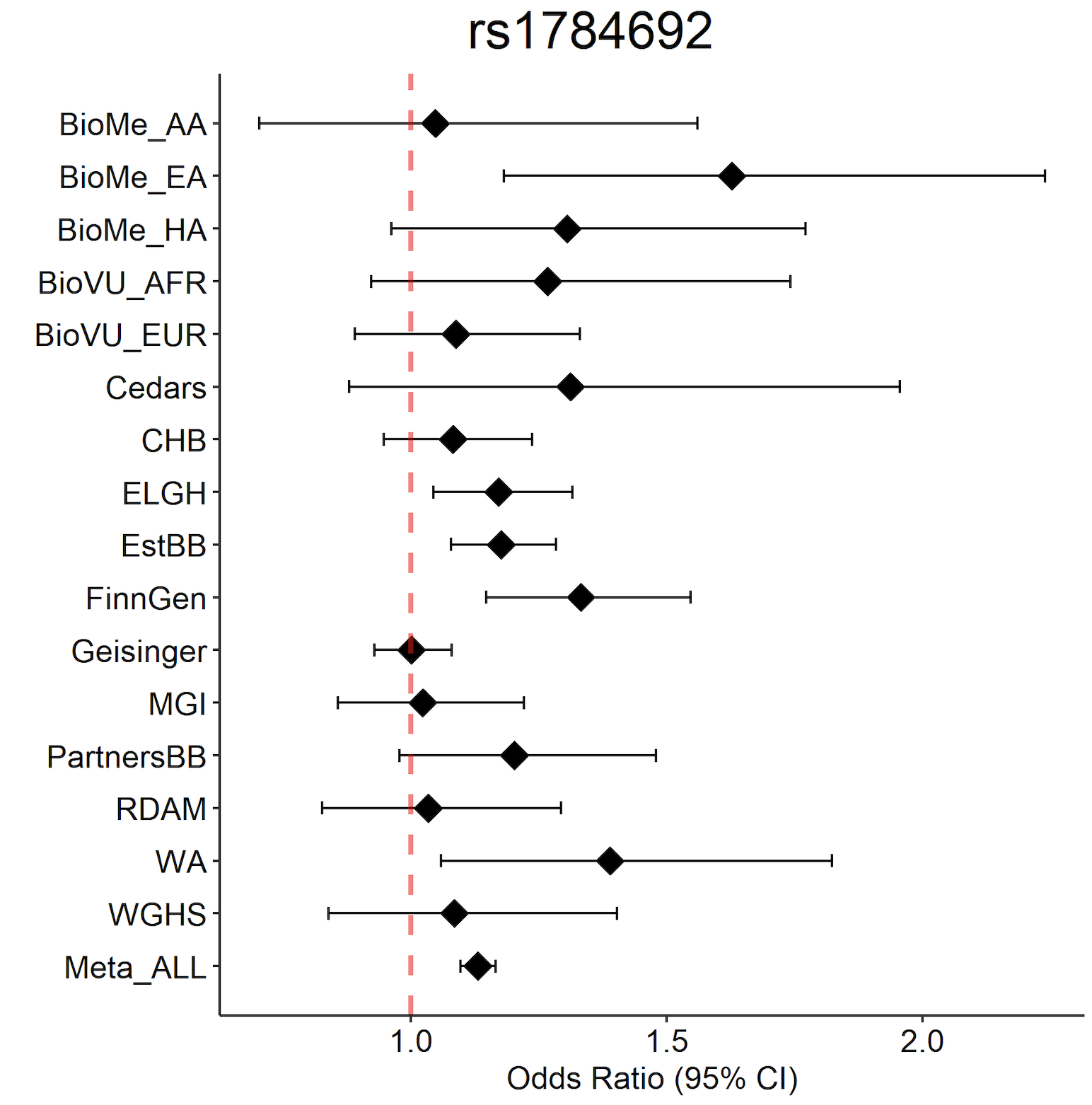

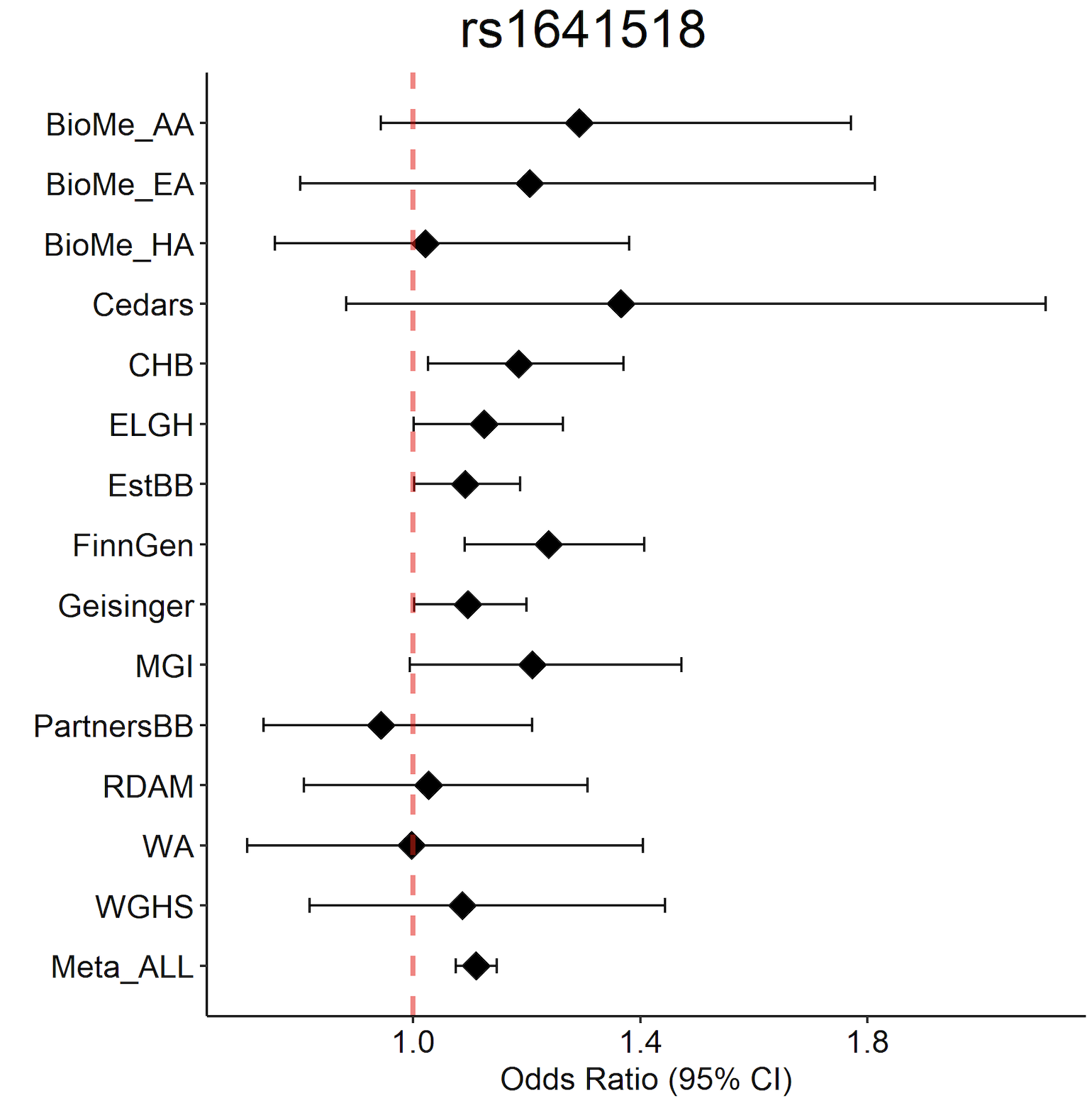

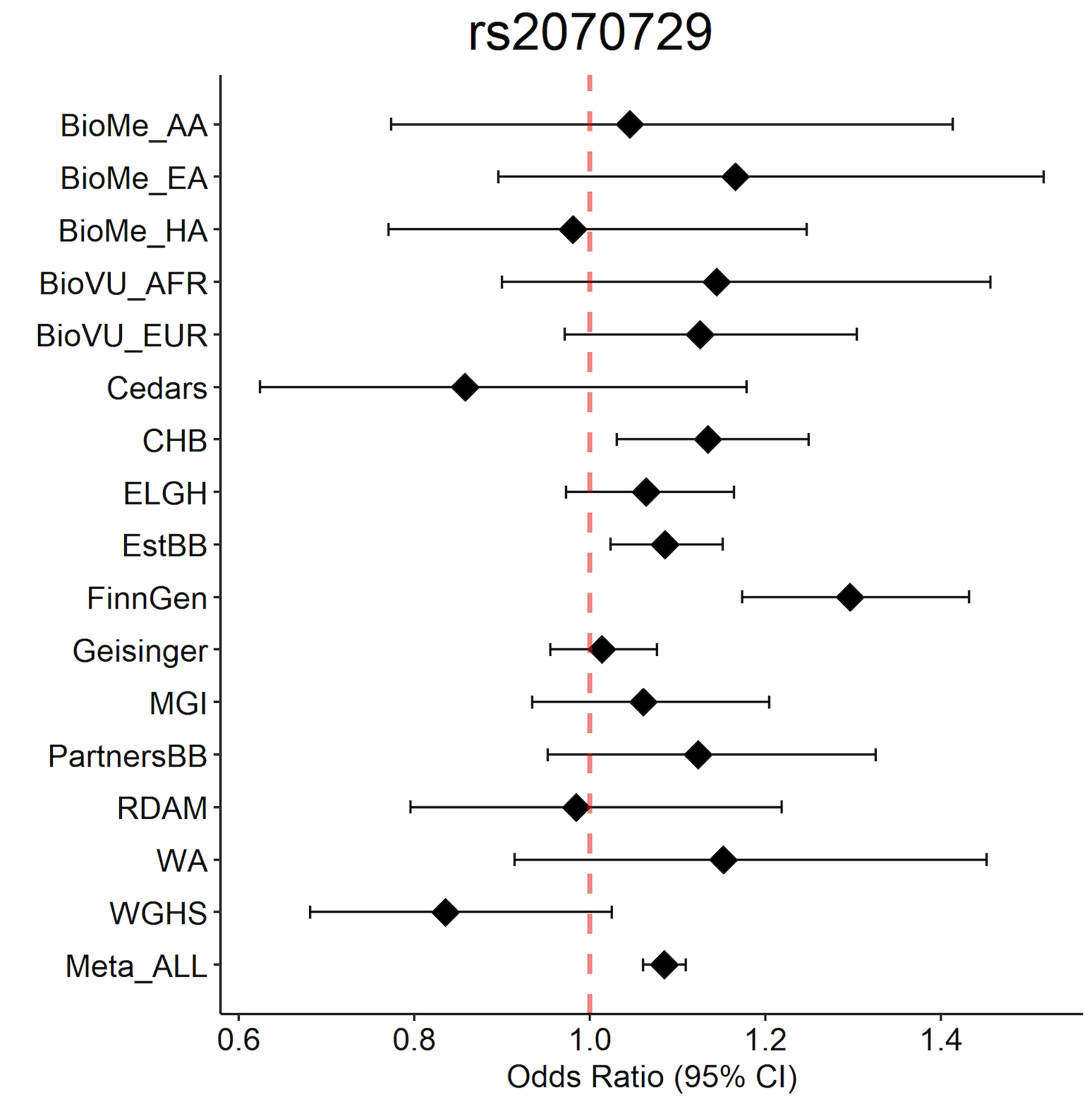

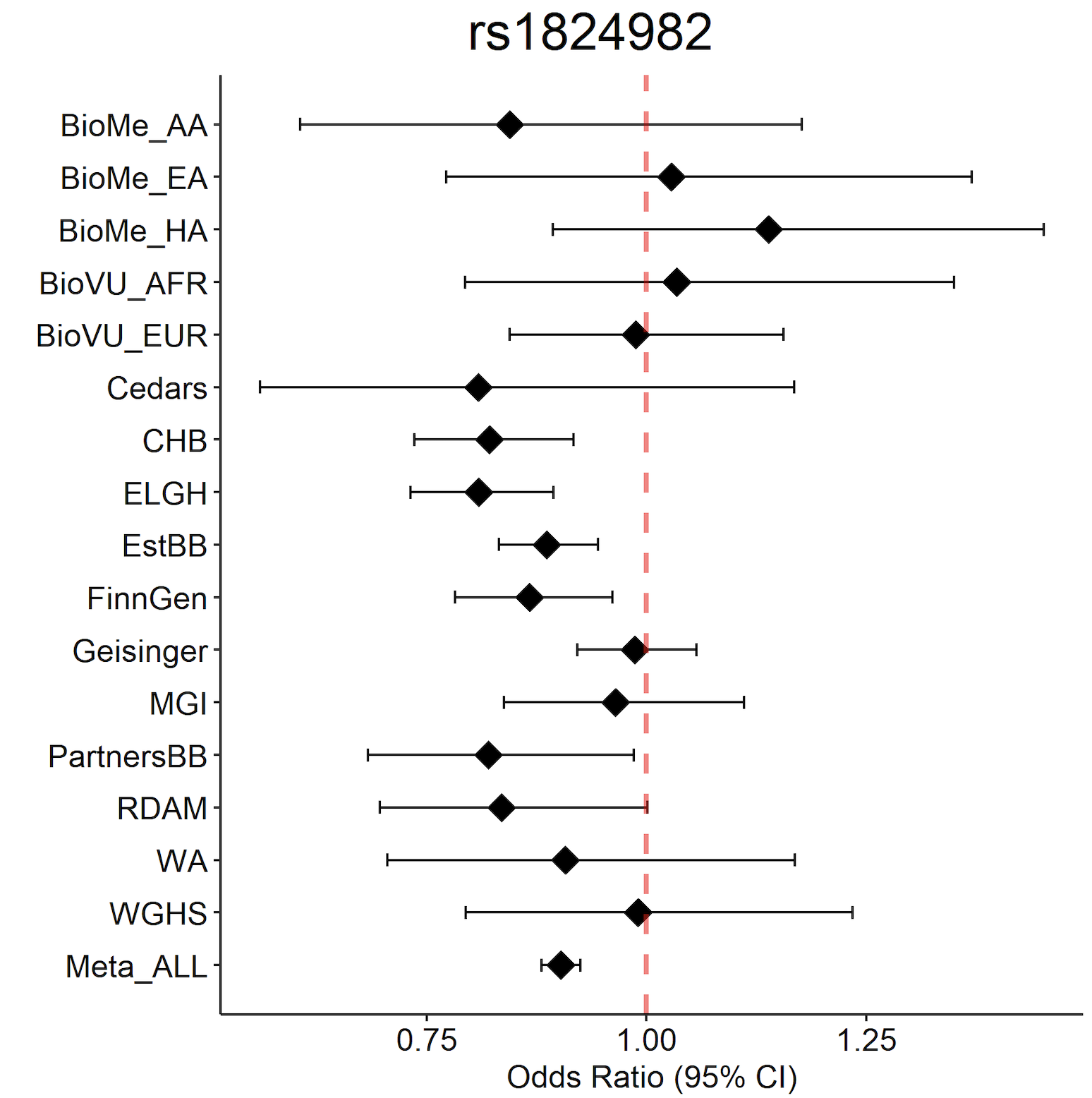

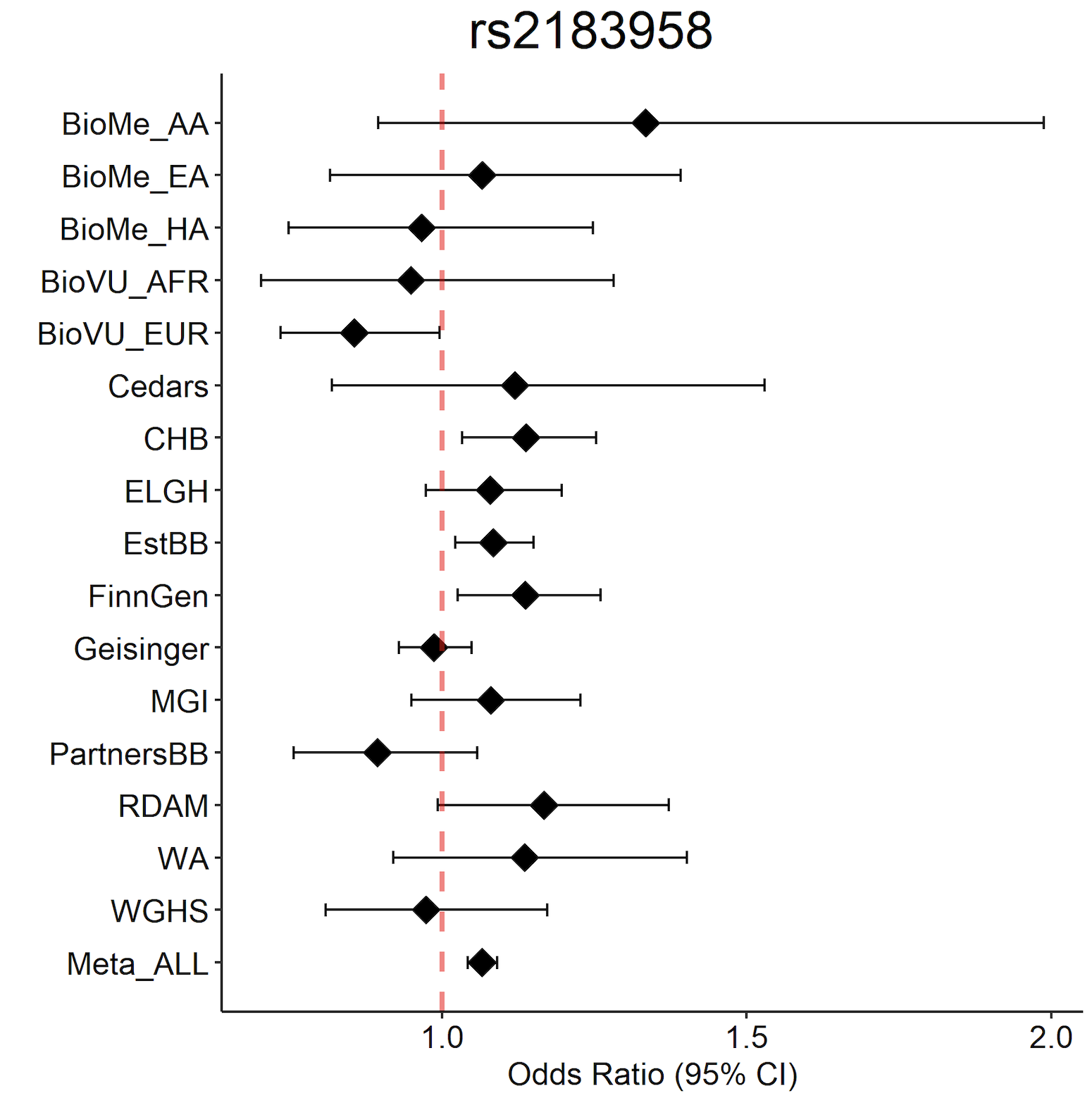

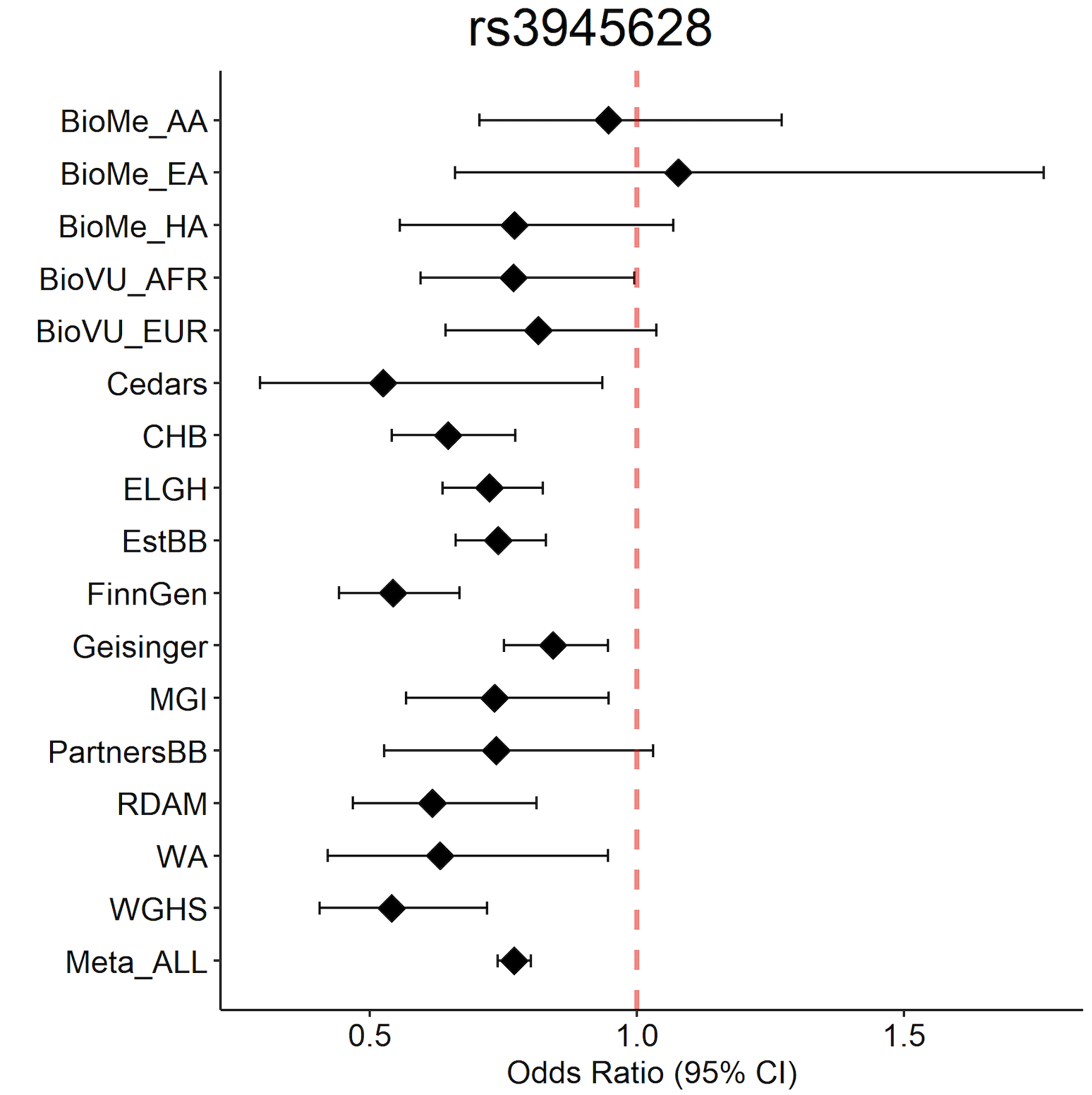

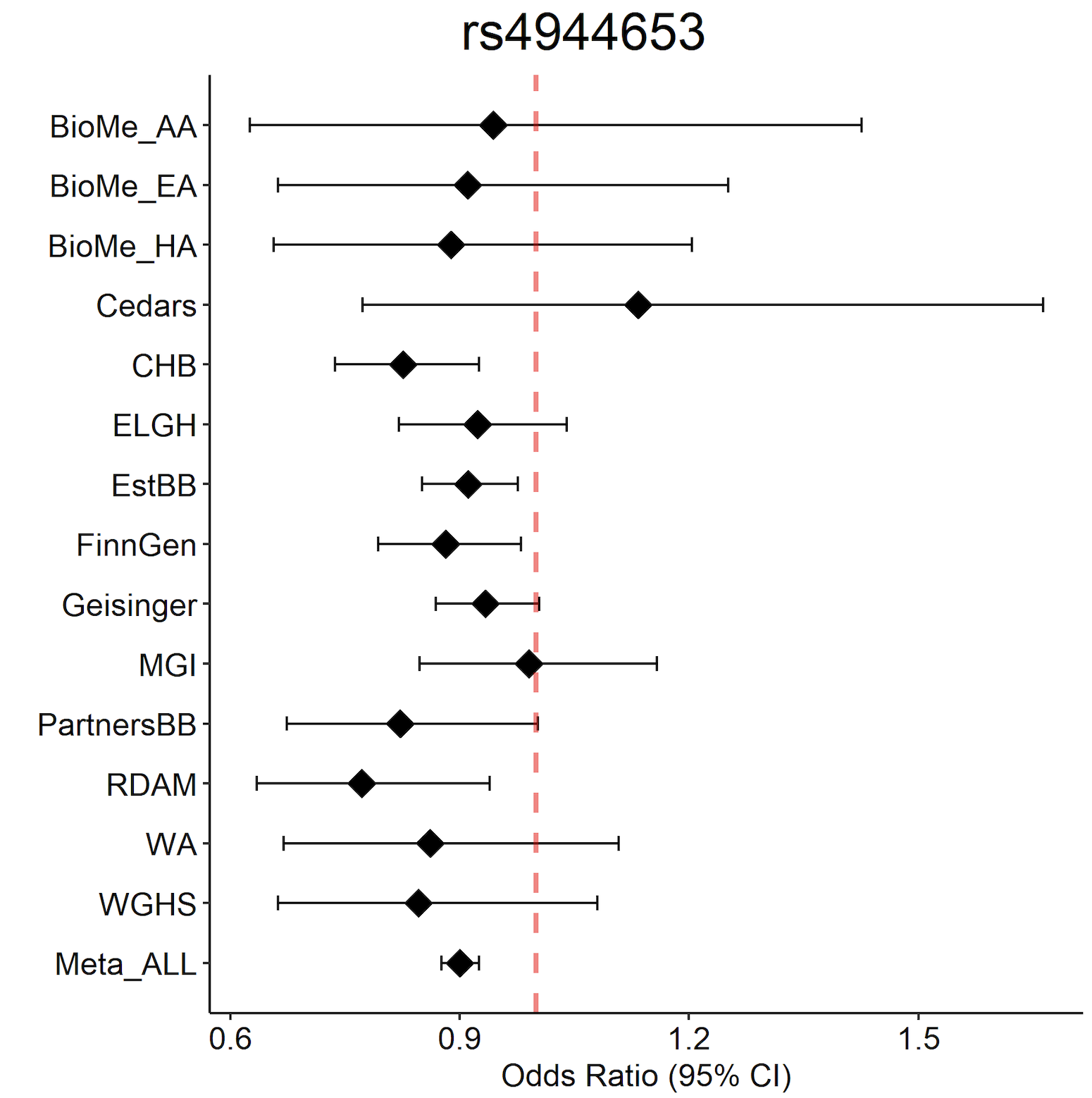

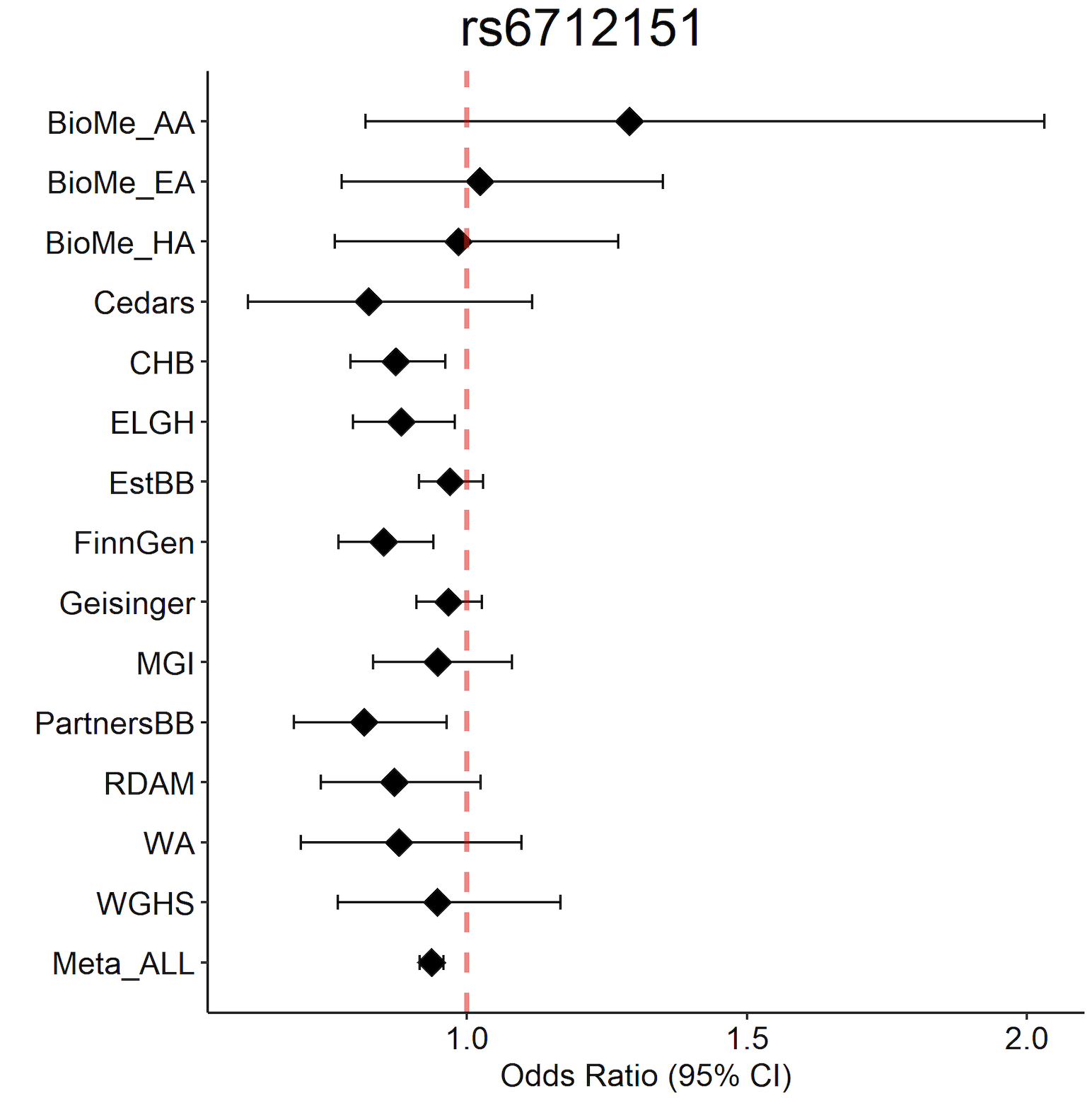

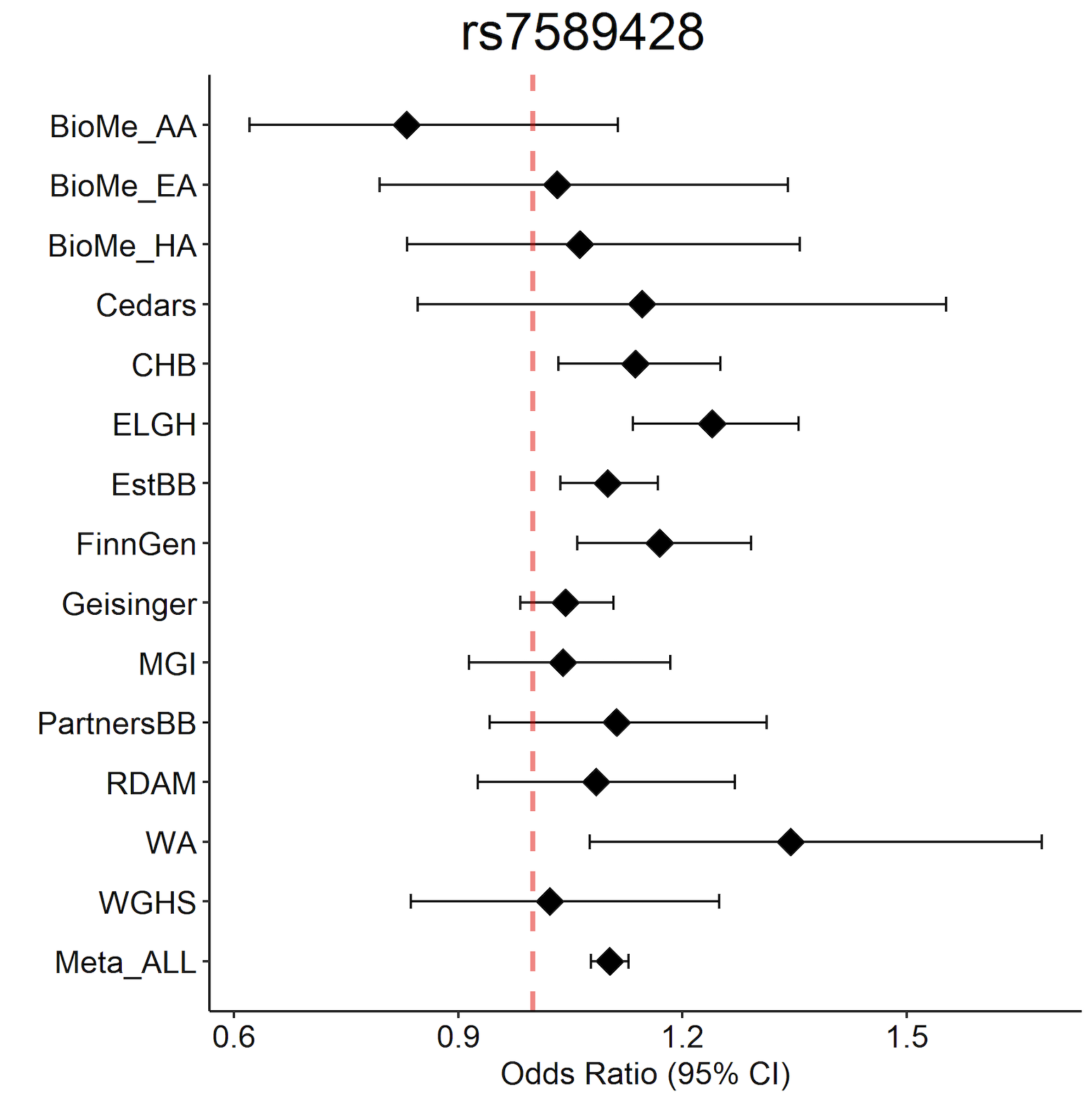

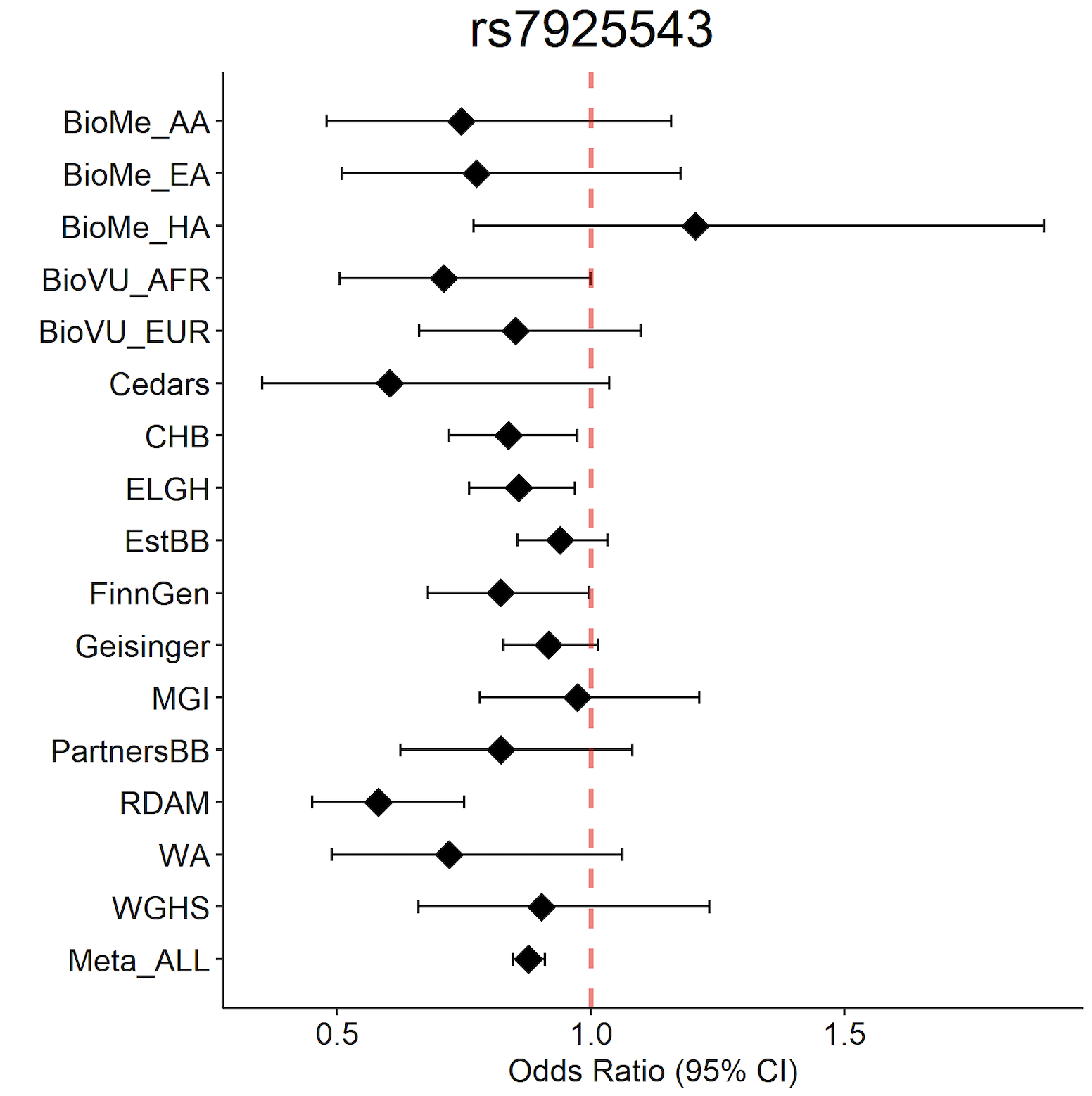

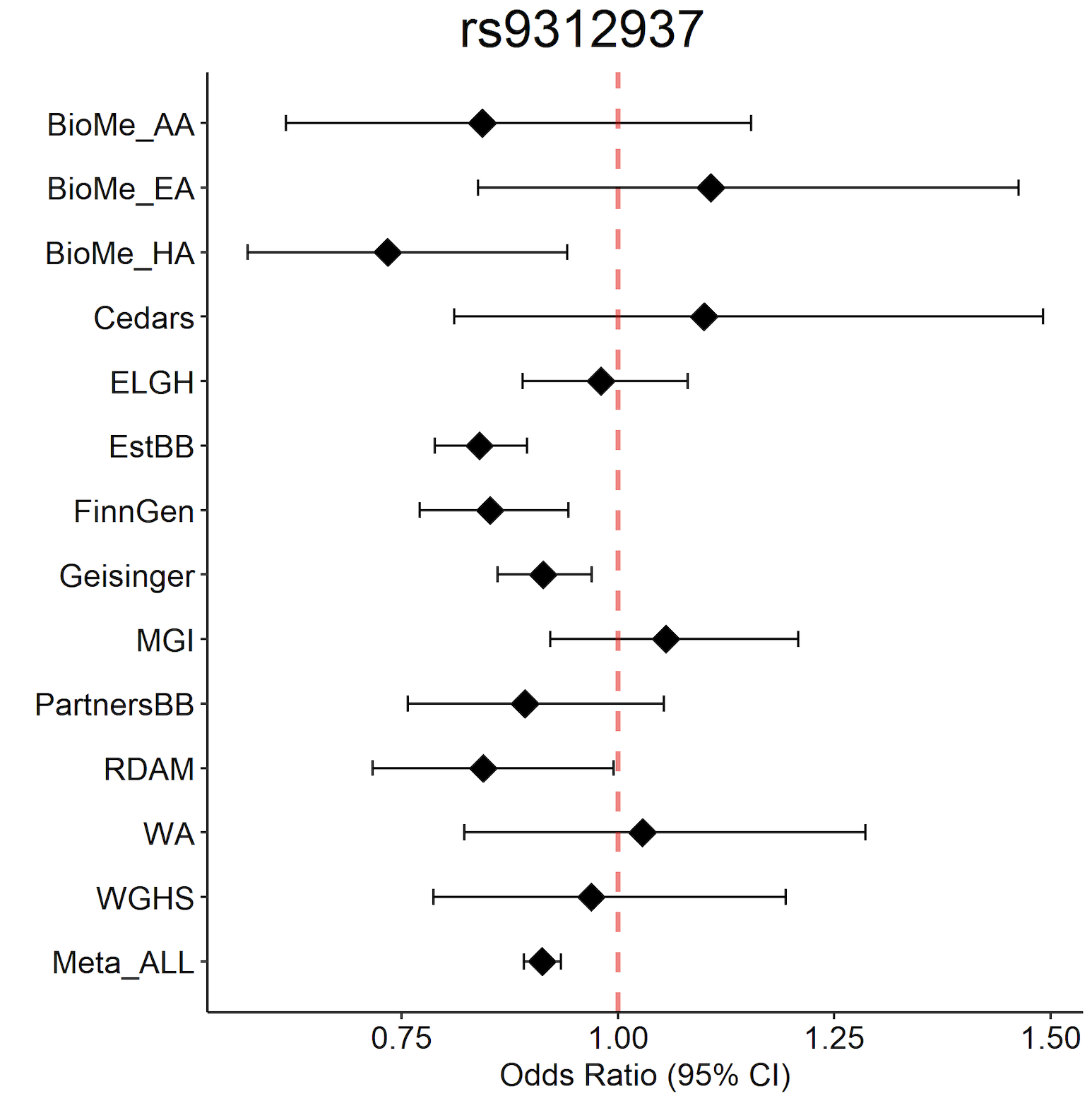

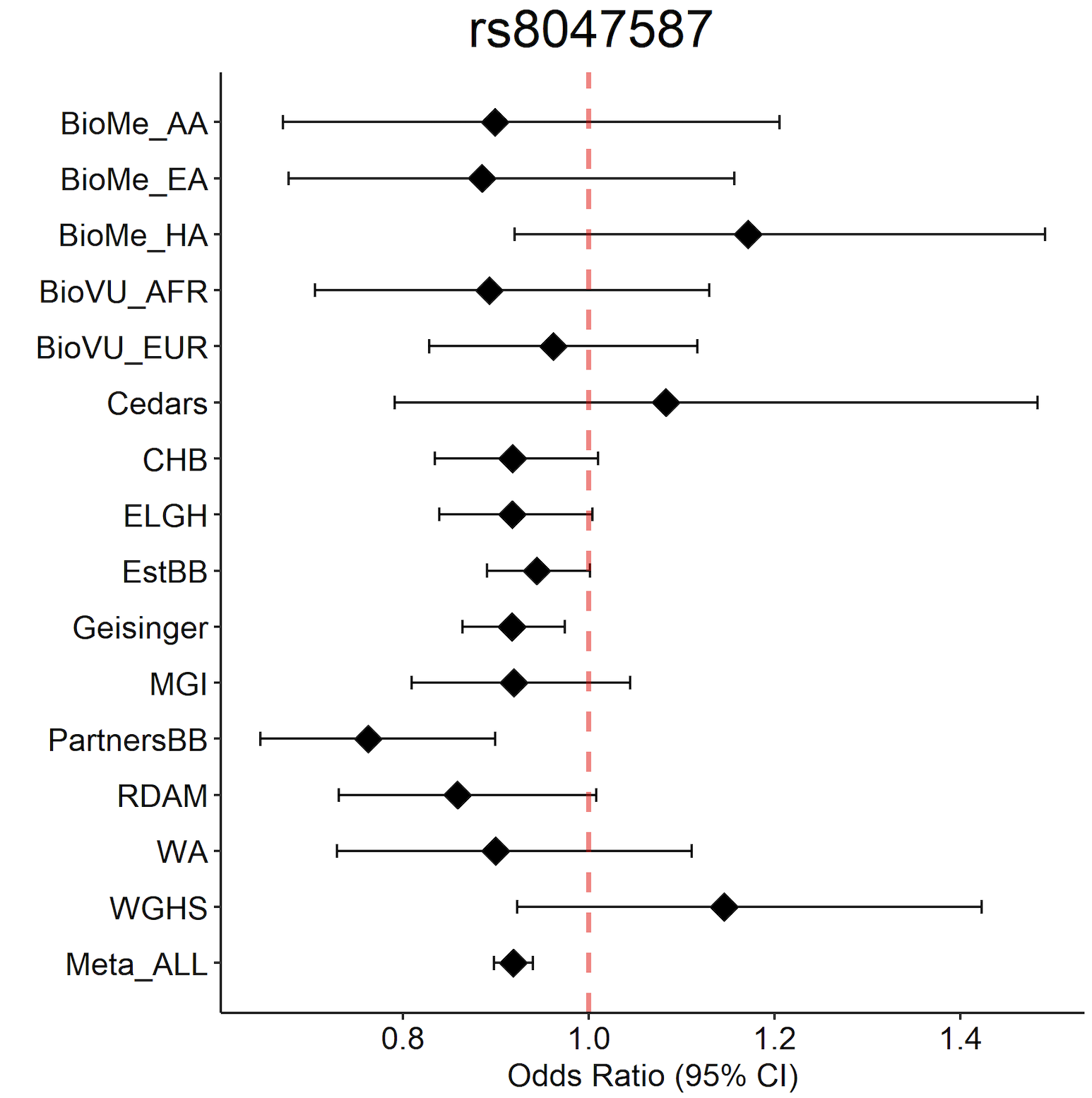

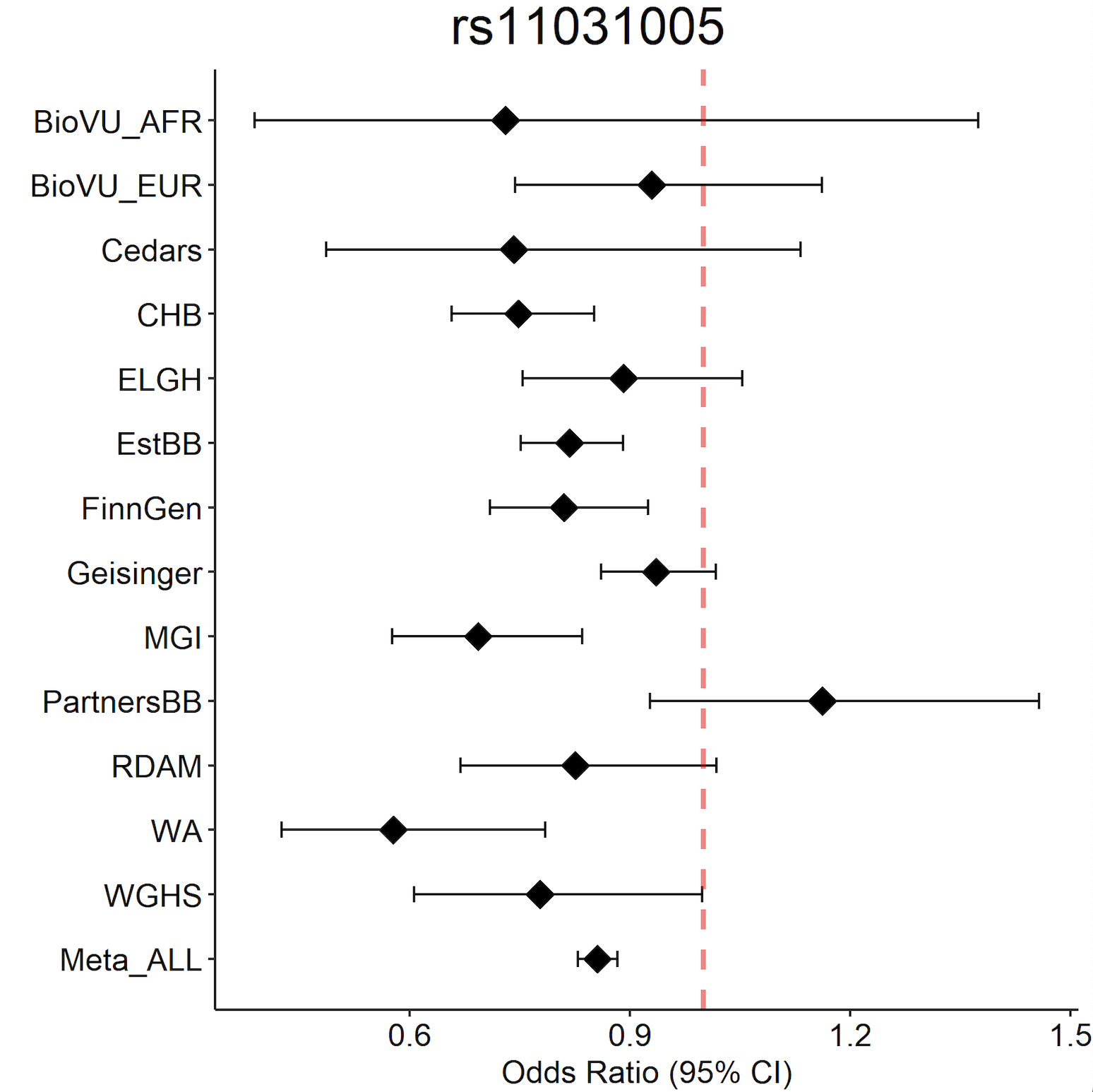

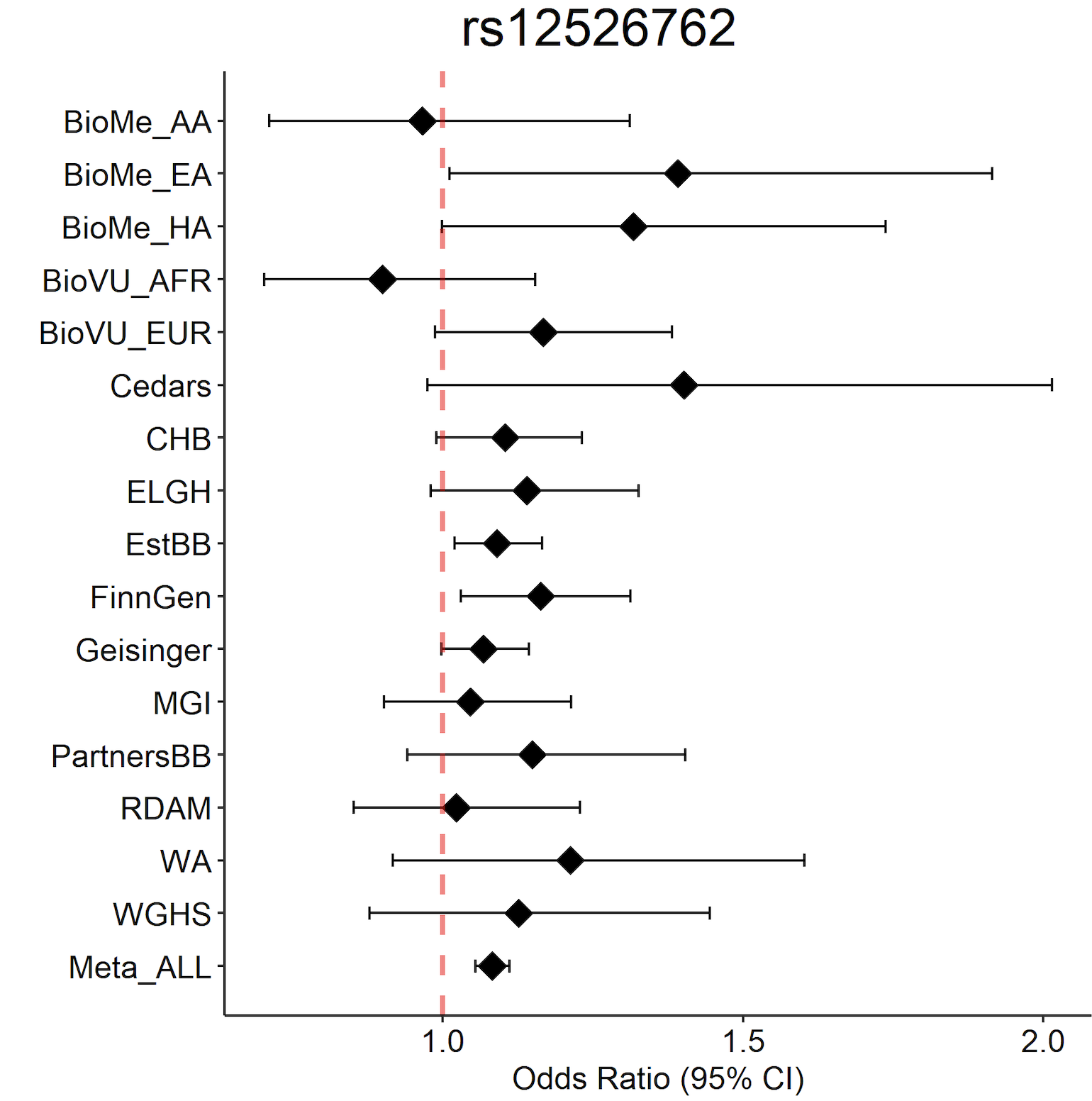

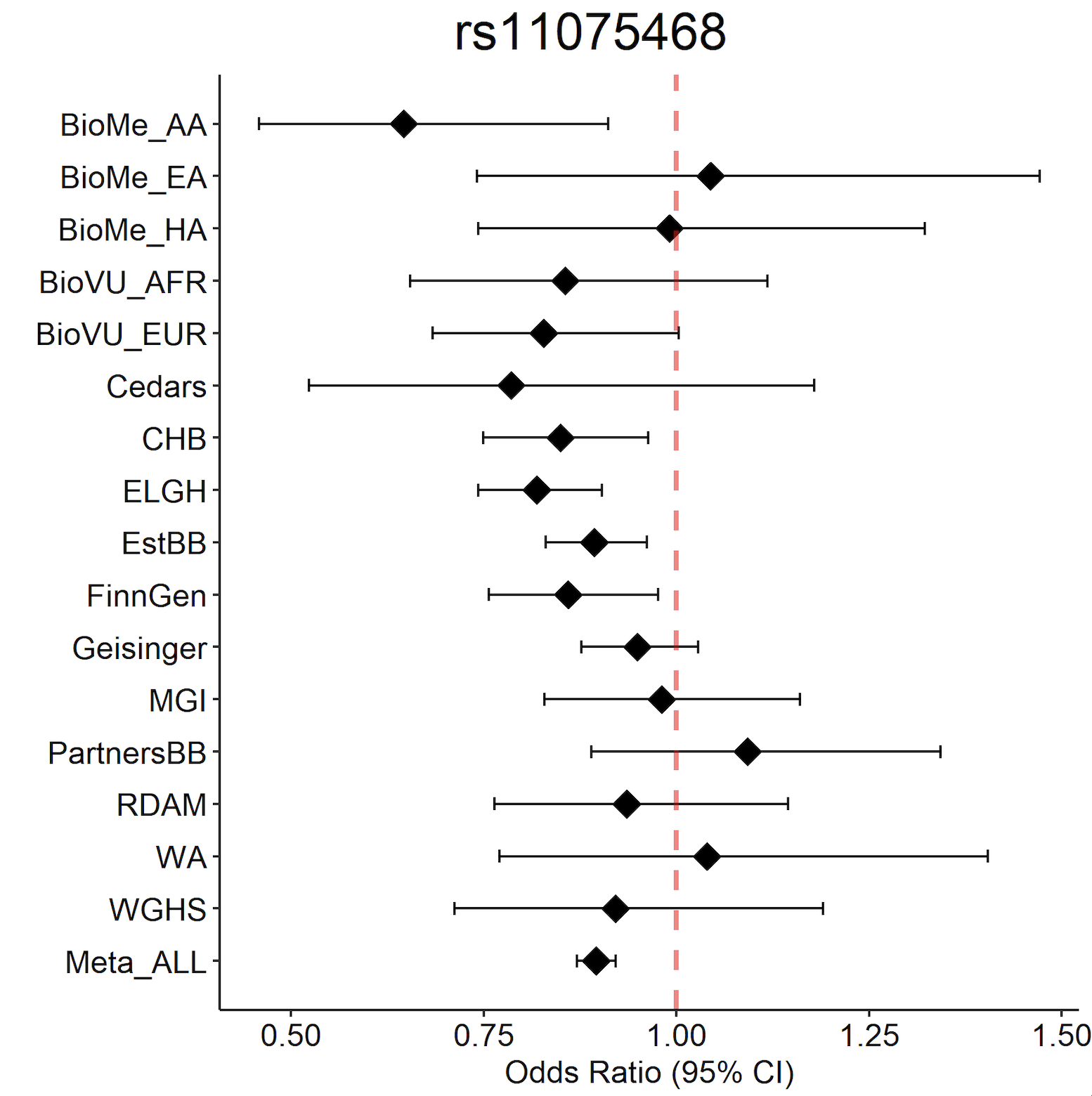

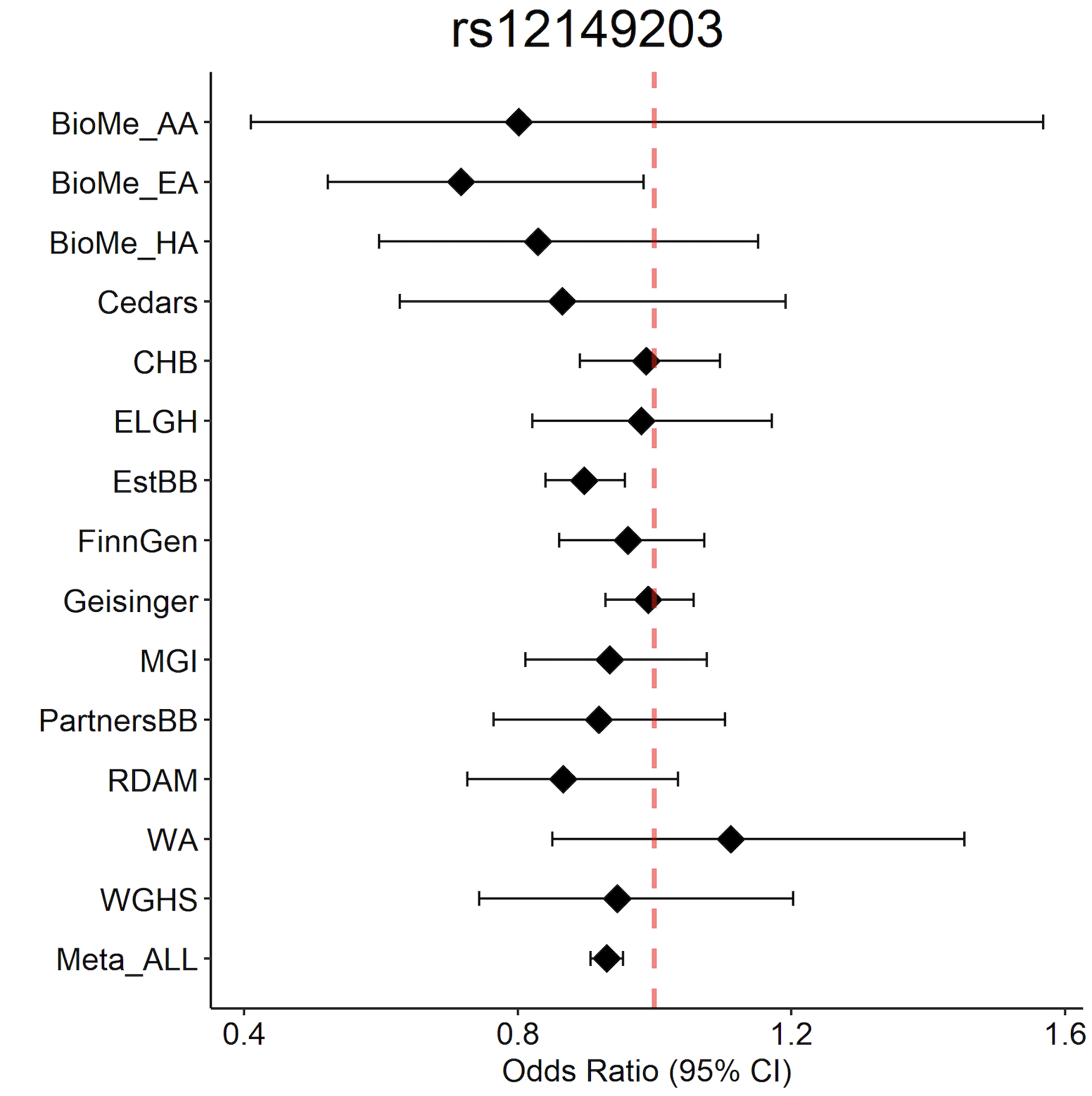

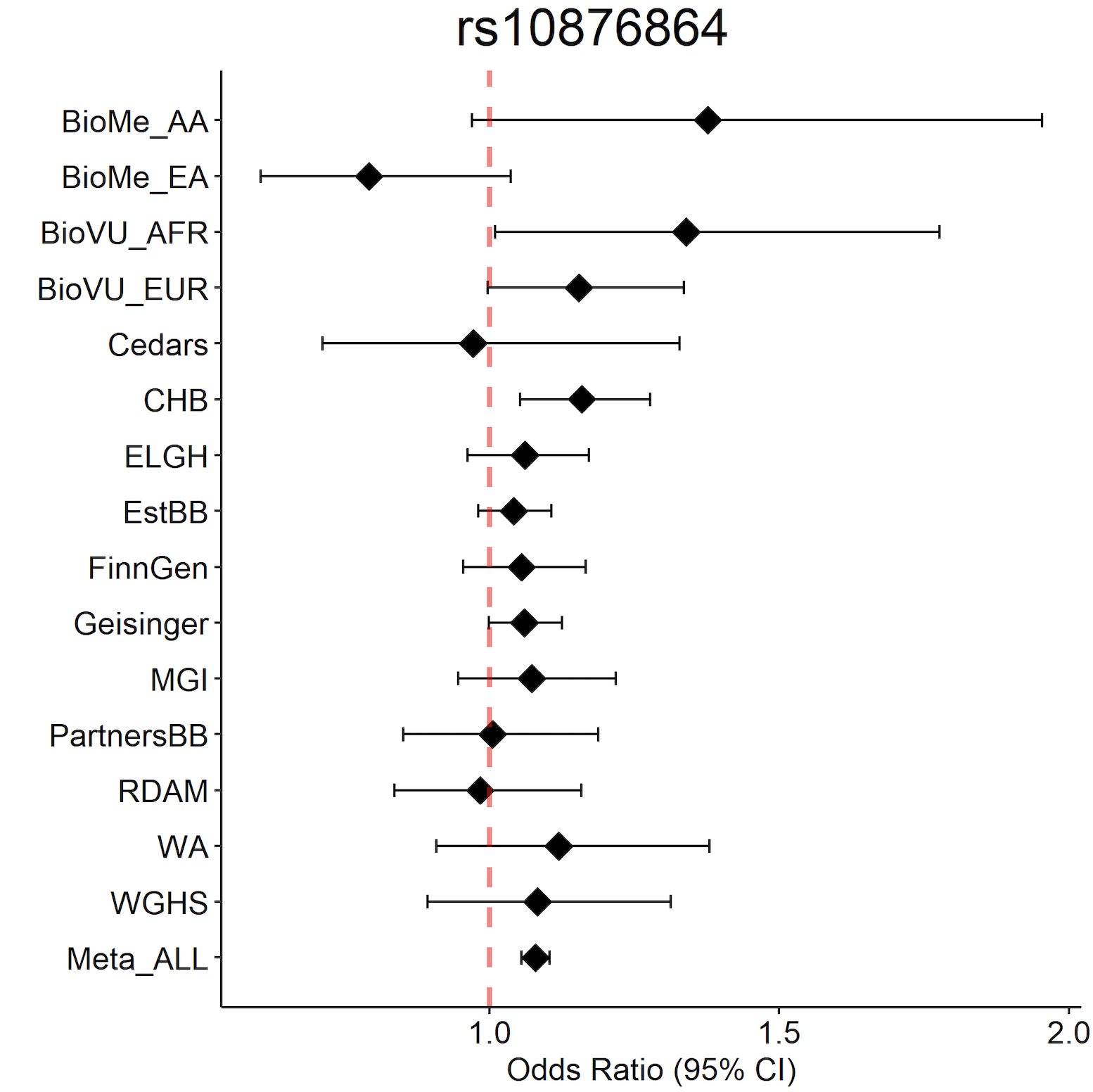

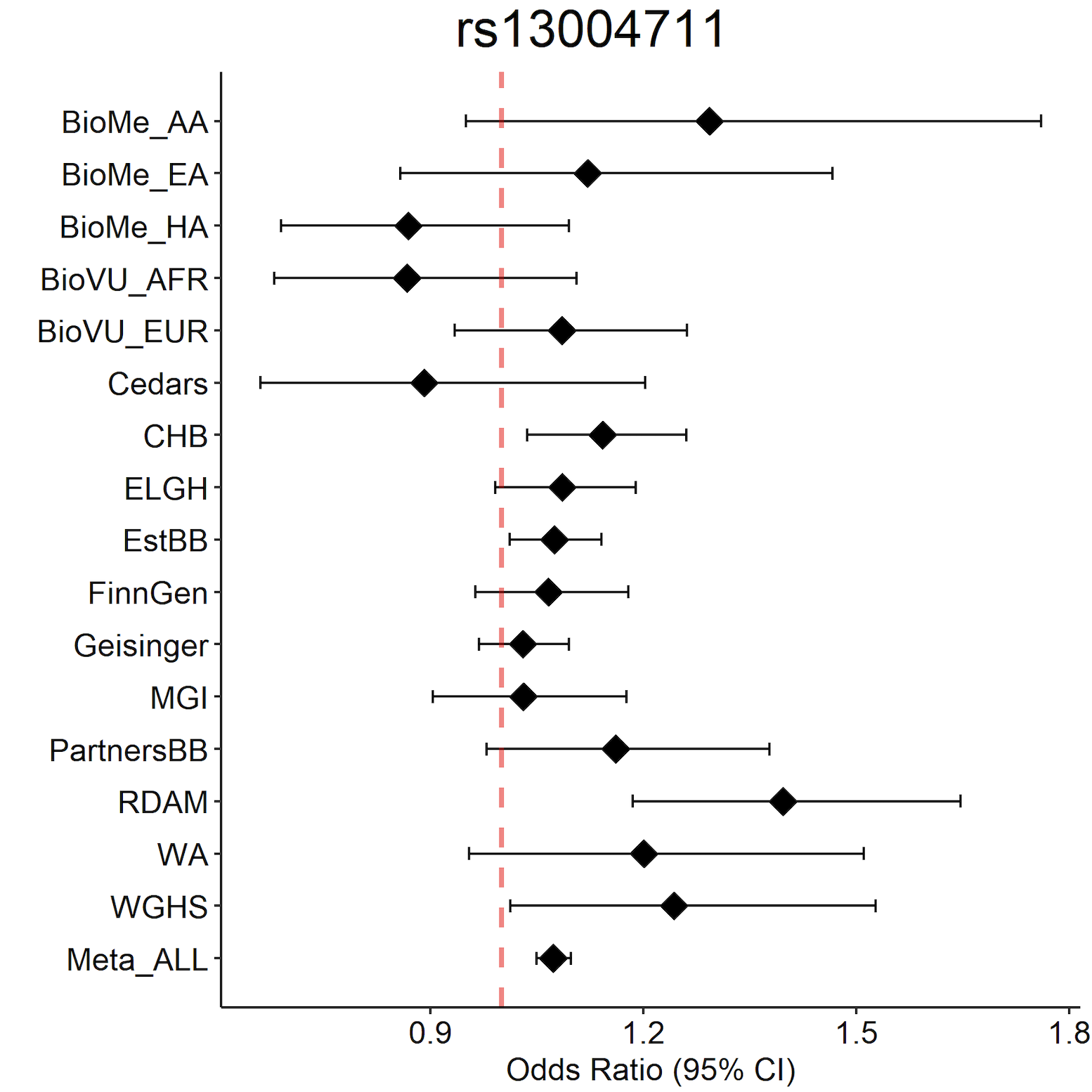

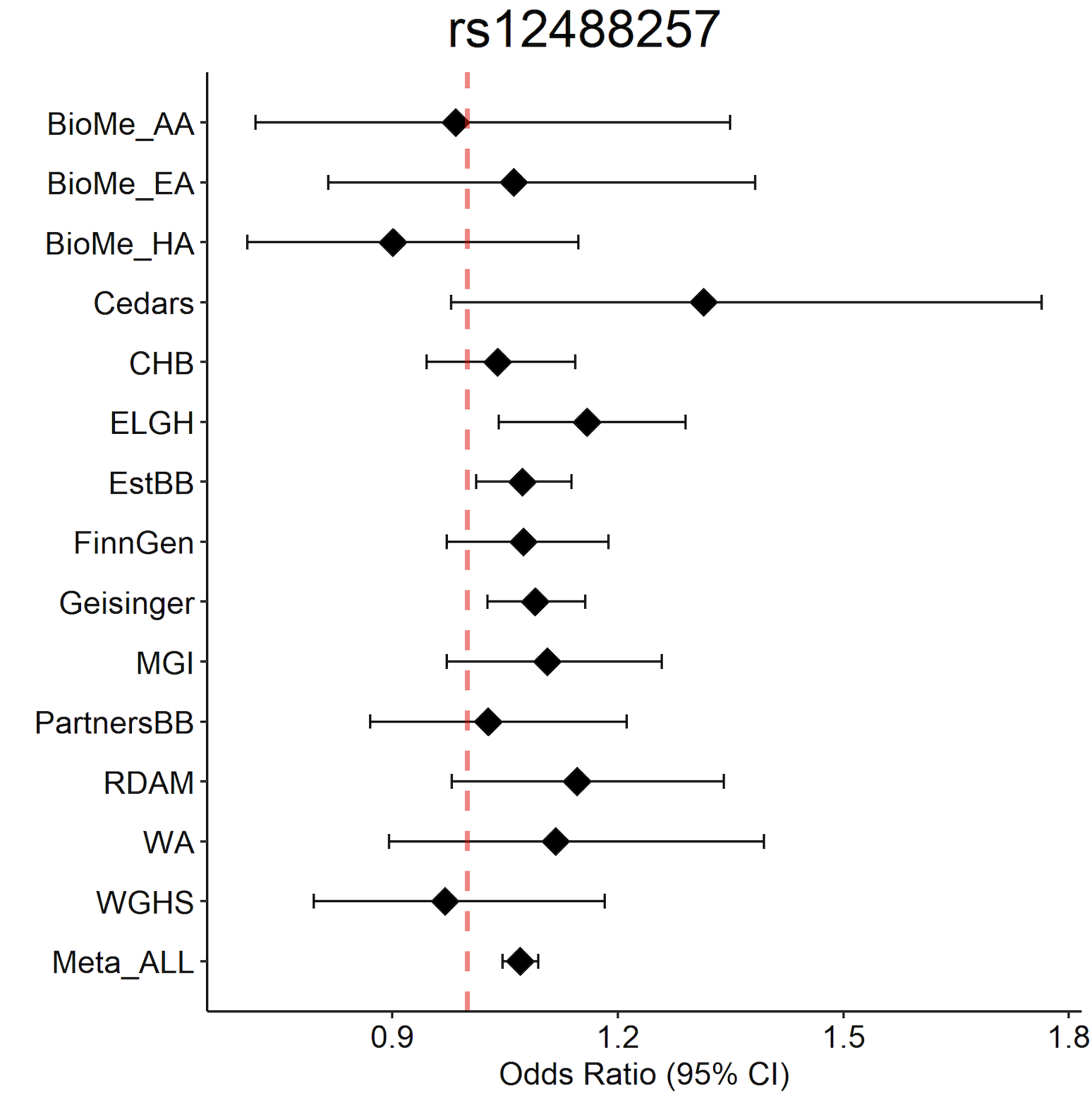

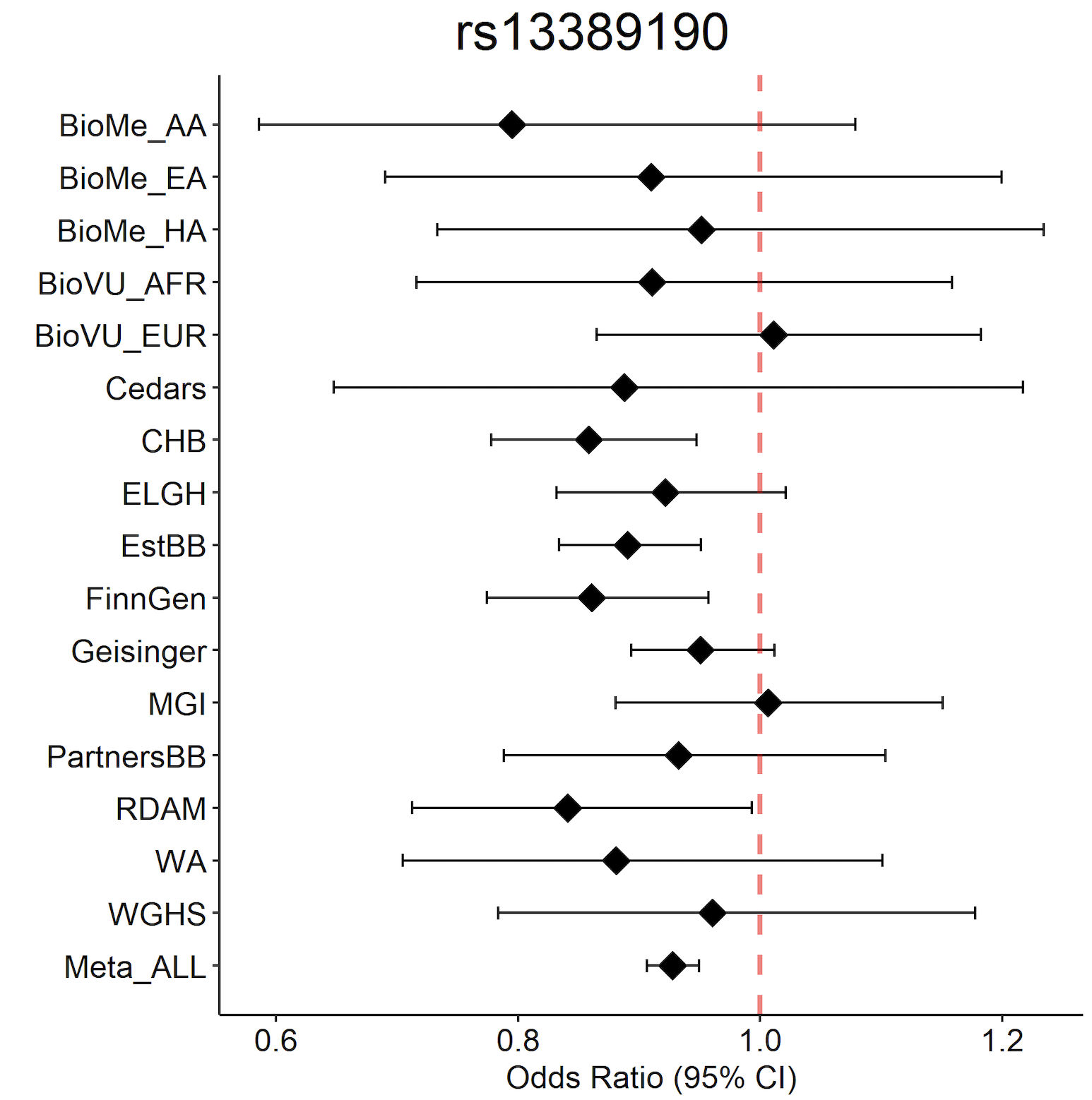

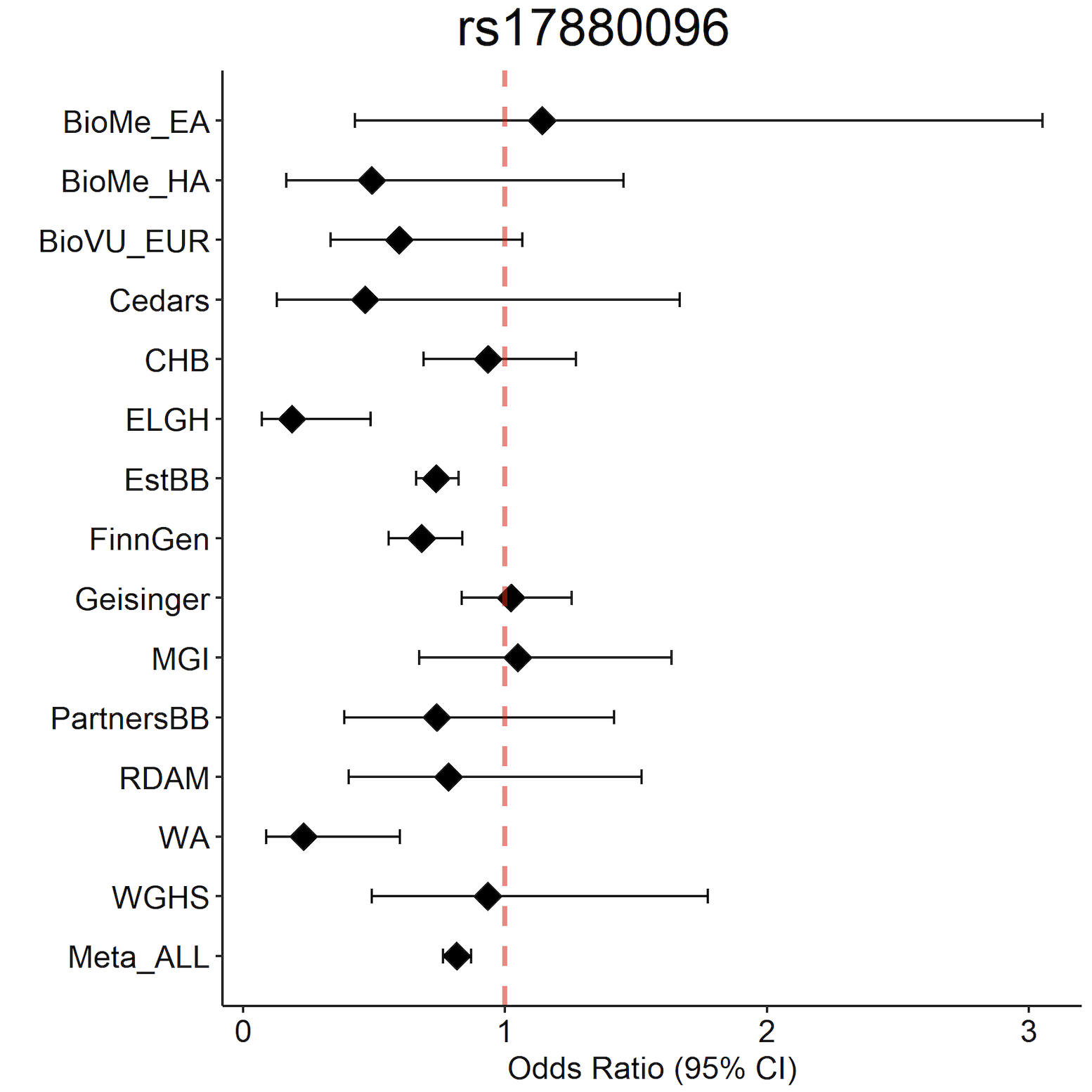

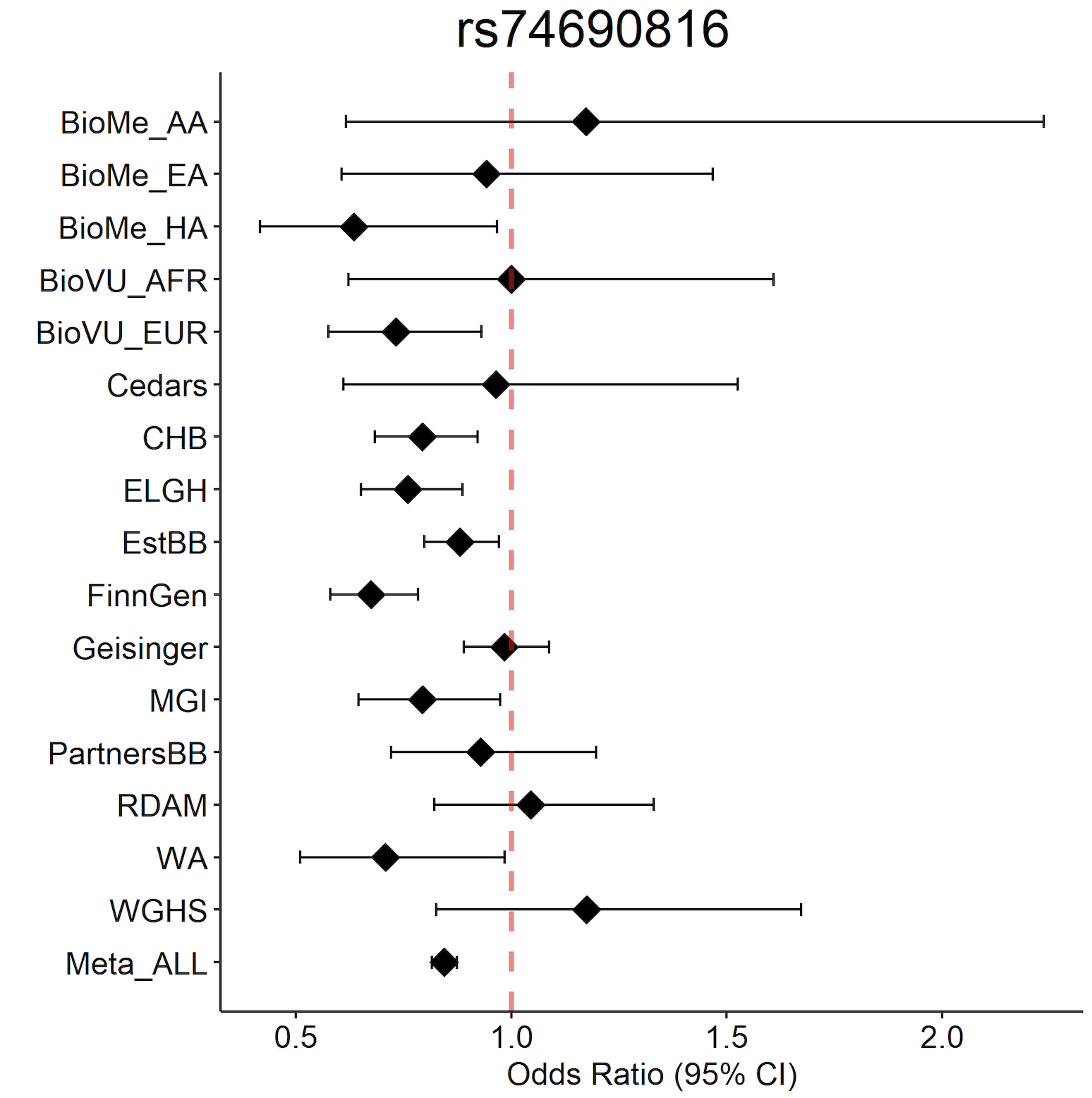

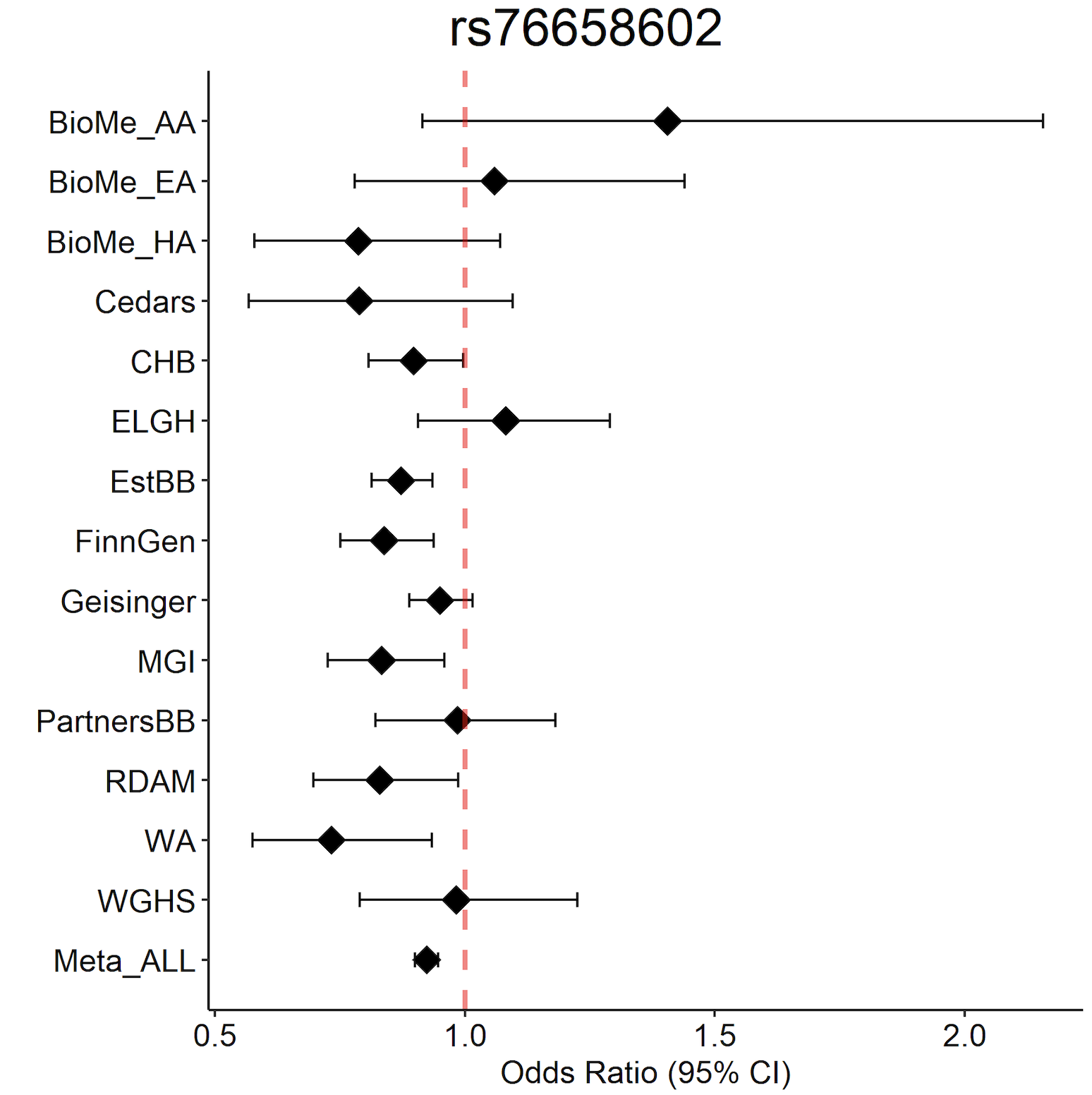

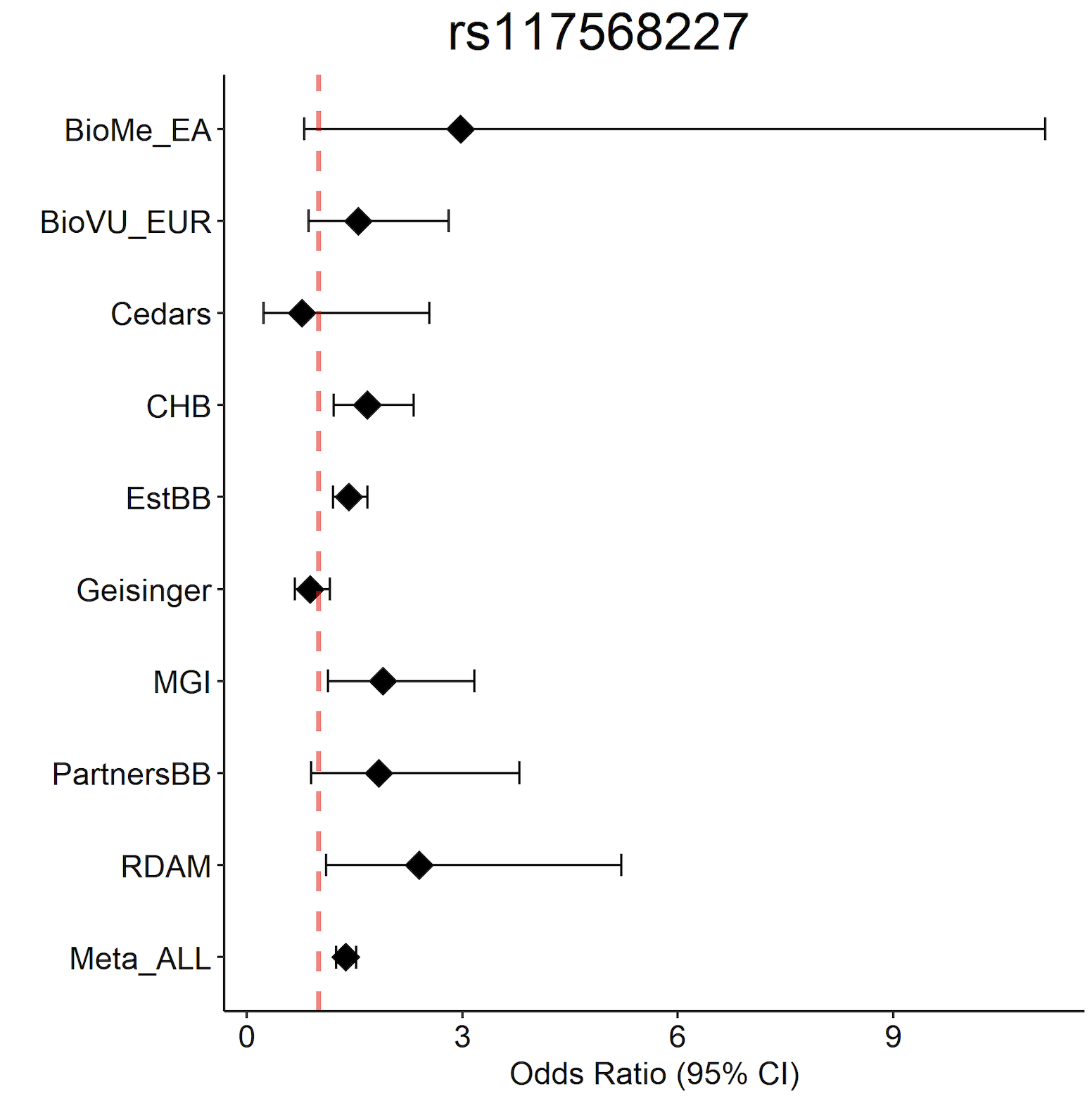

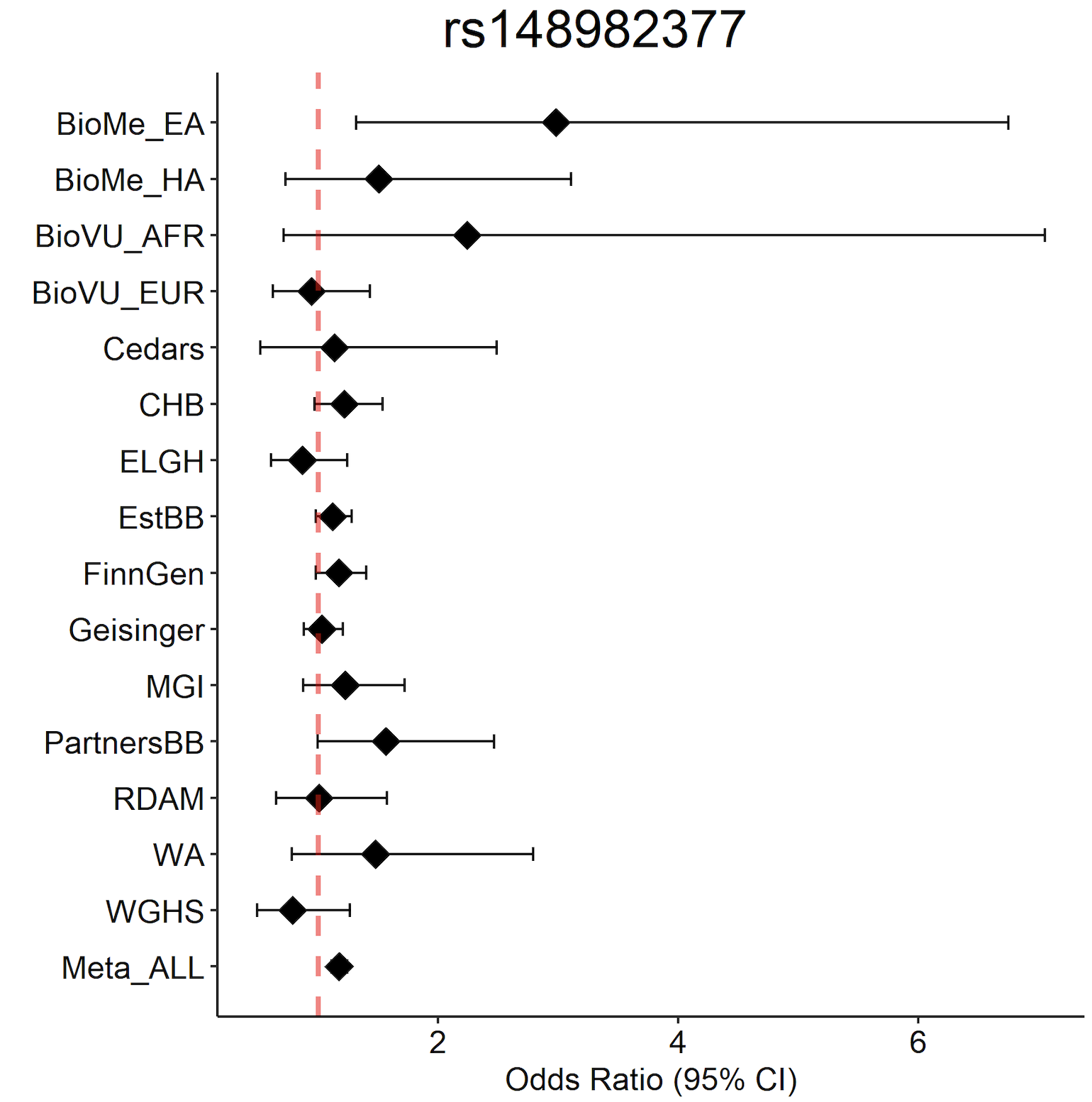

***Supplementary Figure 4*** *- Forest plots for top 29 PCOS associations showing effect sizes and 95% confidence intervals across the studies in the age-adjusted meta-analysis.*

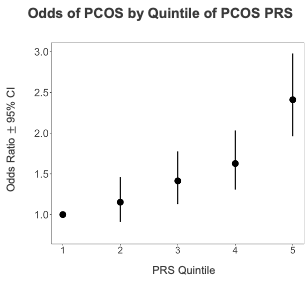

***Supplementary Figure 5*** *- Association of the PCOS-PRS with PCOS in the UK Biobank sample.*

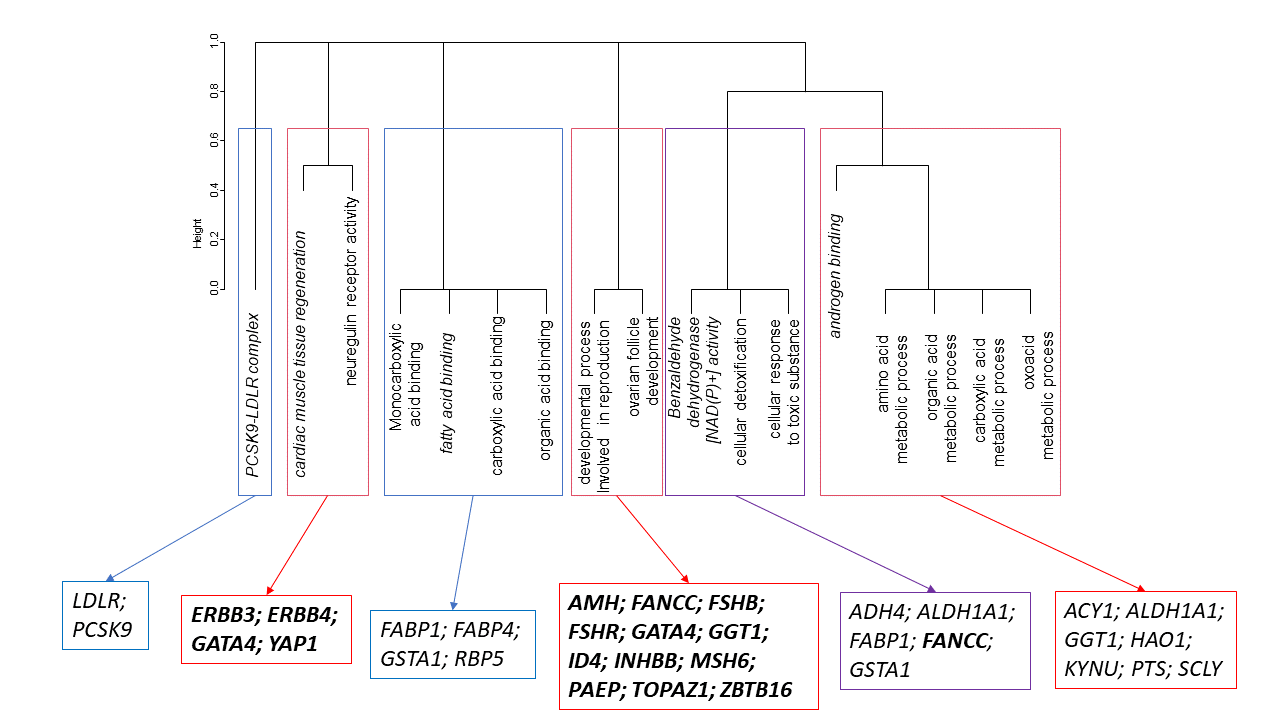

***Supplementary Figure 6*** *- Clustered Pathways based on a combined list of associated genetic loci and proteins associated with a diagnosis coded as E28.* Clusters were generated based on similarity of the intersection between pathways and PCOS genes. Clusters driven by genetic associations are shown in red, with genes in bold. Those driven by proteins are shown in blue, and the one cluster with interactions covering both discovery sets is shown in purple. In each case the strongest associated pathway in each cluster is in italic.

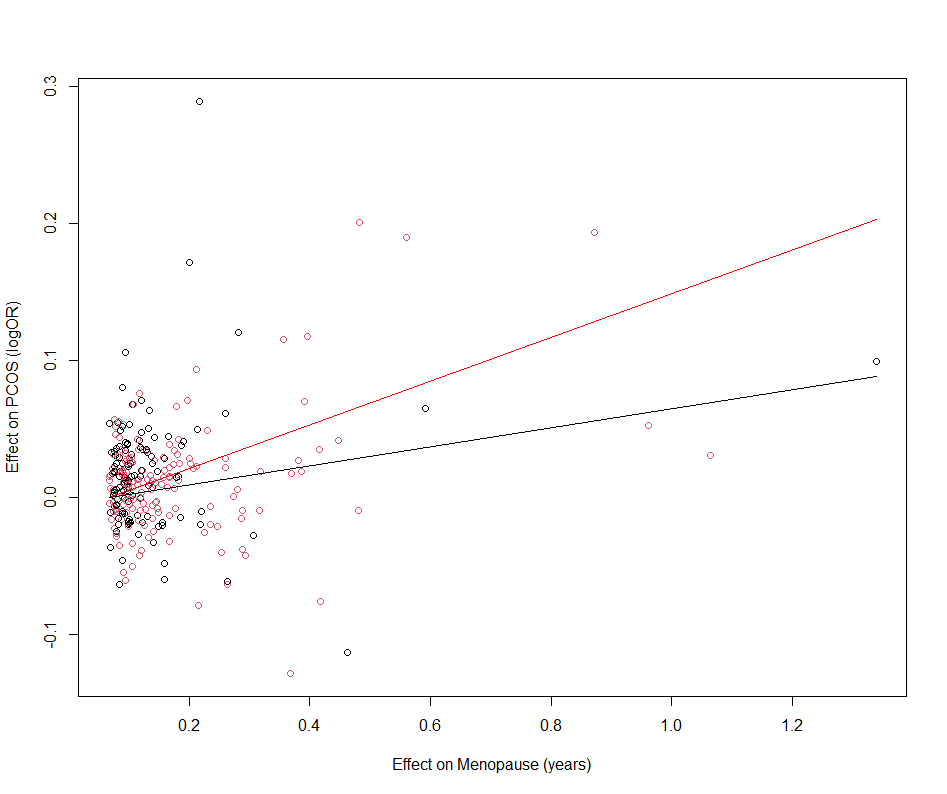

***Supplementary Figure 7*** *- Comparison of the MR split by DDR related menopause genes and (black) non-DDR related menopause variants (red).*

**Supplementary References**

1. Chang S, Dunaif A. Diagnosis of Polycystic Ovary Syndrome: Which Criteria to Use and When? Endocrinol Metab Clin North Am 2021;50:11-23.

2. Day FR, Hinds DA, Tung JY, Stolk L, Styrkarsdottir U, Saxena R, Bjonnes A, Broer L, Dunger DB, Halldorsson BV, Lawlor DA, Laval G, Mathieson I, McCardle WL, Louwers Y, Meun C, Ring S, Scott RA, Sulem P, Uitterlinden AG, Wareham NJ, Thorsteinsdottir U, Welt C, Stefansson K, Laven JS, Ong KK, Perry JR. Causal mechanisms and balancing selection inferred from genetic associations with polycystic ovary syndrome. Nat Commun 2015;6:8464.

3. Ferriman D, Gallwey JD. Clinical assessment of body hair growth in women. J Clin Endocrinol Metab 1961;21:1440-7.

4. Rotterdam EA-SPcwg. Revised 2003 consensus on diagnostic criteria and long-term health risks related to polycystic ovary syndrome (PCOS). Hum Reprod 2004;19:41-7.

5. Zhu Z, Zhang F, Hu H, Bakshi A, Robinson MR, Powell JE, Montgomery GW, Goddard ME, Wray NR, Visscher PM, Yang J. Integration of summary data from GWAS and eQTL studies predicts complex trait gene targets. Nat Genet 2016;48:481-7.

6. Perry JR, Hsu YH, Chasman DI, Johnson AD, Elks C, Albrecht E, Andrulis IL, Beesley J, Berenson GS, Bergmann S, Bojesen SE, Bolla MK, Brown J, Buring JE, Campbell H, Chang-Claude J, Chenevix-Trench G, Corre T, Couch FJ, Cox A, Czene K, D'Adamo A P, Davies G, Deary IJ, Dennis J, Easton DF, Engelhardt EG, Eriksson JG, Esko T, Fasching PA, Figueroa JD, Flyger H, Fraser A, Garcia-Closas M, Gasparini P, Gieger C, Giles G, Guenel P, Hagg S, Hall P, Hayward C, Hopper J, Ingelsson E, kConFab i, Kardia SL, Kasiman K, Knight JA, Lahti J, Lawlor DA, Magnusson PK, Margolin S, Marsh JA, Metspalu A, Olson JE, Pennell CE, Polasek O, Rahman I, Ridker PM, Robino A, Rudan I, Rudolph A, Salumets A, Schmidt MK, Schoemaker MJ, Smith EN, Smith JA, Southey M, Stockl D, Swerdlow AJ, Thompson DJ, Truong T, Ulivi S, Waldenberger M, Wang Q, Wild S, Wilson JF, Wright AF, Zgaga L, ReproGen C, Ong KK, Murabito JM, Karasik D, Murray A. DNA mismatch repair gene MSH6 implicated in determining age at natural menopause. Hum Molec Genet 2014;23:2490-7.

7. GTex Consortium, Battle A, Brown CD, Engelhardt BE, Montgomery SB. Genetic effects on gene expression across human tissues. Nature 2017;550:204-13.

8. Khera AV, Chaffin M, Aragam KG, Haas ME, Roselli C, Choi SH, Natarajan P, Lander ES, Lubitz SA, Ellinor PT, Kathiresan S. Genome-wide polygenic scores for common diseases identify individuals with risk equivalent to monogenic mutations. Nat Genet 2018;50:1219-24. 9. Udler MS, Kim J, von Grotthuss M, Bonas-Guarch S, Cole JB, Chiou J, Christopher DAoboM, the I, Boehnke M, Laakso M, Atzmon G, Glaser B, Mercader JM, Gaulton K, Flannick J, Getz G, Florez JC. Type 2 diabetes genetic loci informed by multi-trait associations point to disease mechanisms and subtypes: A soft clustering analysis. PLoS Med 2018;15:e1002654.

10. Eastwood SV, Mathur R, Atkinson M, Brophy S, Sudlow C, Flaig R, de Lusignan S, Allen N, Chaturvedi N. Algorithms for the Capture and Adjudication of Prevalent and Incident Diabetes in UK Biobank. PLoS One 2016;11:e0162388.

11. Smith DJ, Nicholl BI, Cullen B, Martin D, Ul-Haq Z, Evans J, Gill JM, Roberts B, Gallacher J, Mackay D, Hotopf M, Deary I, Craddock N, Pell JP. Prevalence and characteristics of probable major depression and bipolar disorder within UK biobank: cross-sectional study of 172,751 participants. PLoS One 2013;8:e75362
